# Health Signatures During COVID-19: A Precision Fitness Case Study

**DOI:** 10.1101/2020.12.07.20245001

**Authors:** Erin P. Pollet, Aishwarya Sathish, Zacharie Maloney, Byron L. Long, Jennifer Brethen, Amina Ann Qutub

## Abstract

**BACKGROUND:** Stay-at-home orders have proven a controversial, while effective, method of SARS-CoV-2 containment. However objective measures of how the pandemic and stay-at-home orders are affecting the daily health of uninfected individuals have been lacking.

**METHODS:** We investigated the effect of pandemic-related events on 61 individuals in San Antonio, Texas whose daily activity and sleep data were recorded via wearable activity trackers from April 2019 to August 2020. We assessed changes in six fitness metrics (steps walked, resting heart rate, sedentary minutes, wake duration after sleep onset, rapid eye movement (REM) duration, total sleep duration). Cluster analysis and time-course analysis identified trends in activity before, after and during stay-at-home orders. Quantitative measures of activities were compared to survey responses.

**RESULTS:** Four behavior patterns during stay-at-home orders were identified. Most individuals suffered declines in healthy habits compared to their daily activity in 2019 and early 2020 (e.g., up to −60% steps walked). Inflection points corresponded with key dates relevant to SARS-CoV-2 including the first reported case in the U.S. (Feb 29) and city-wide stay-at-home orders (Mar 23). Pre-existing conditions (diabetes, asthma) were associated with a steeper than average decline in sleep quality during stay-at-home orders. Unexpectedly, we also identified a group of predominately male individuals who improved their daily fitness during stay-at-home orders.

**CONCLUSIONS:** Objective measures of daily activity indicated most individuals’ fitness suffered at the onset of stay-at-home orders and slowly returned towards baseline. For a subset of individuals, fitness quantitatively improved – *better sleep, more exercise, lower resting heart rate* – during stay-at-home orders.

## Introduction

Cases of severe acute respiratory syndrome coronavirus 2 (SARS-CoV-2) reached pandemic levels within months of the virus emerging in late 2019^1^, and infections have yet to abate in the U.S. as of late 2020. Governments continue to struggle with ways to contain the virus while preserving their economies. One controversial, but effective, method of viral containment is strictly enforced stay-at-home orders^2^. While this strategy proved successful in East Asian^3^, Oceanic^4,5^ and European countries^6,7^ to varying degrees, the contentiousness of stay-at-home orders in the U.S. has caused policymakers to question the long-term feasibility of the approach. Beyond economic and political ramifications, the effects of stay-at-home rules on the physical and mental health in otherwise healthy individuals need to be weighed^8,9^. However, a much-needed, scientific, quantitative study of the effects of the orders has been lacking.

In this study, we fill this knowledge gap by addressing how the pandemic and stay-at-home orders are objectively changing daily habits. We investigated the effect of stay-at-home orders on 61 individuals in Texas whose daily activity and sleep data was recorded via wearable activity trackers for >12 months, from April 2019 to August 2020. We observed changes across six physical fitness metrics^10^ (steps walked, resting heart rate, sedentary minutes, wake duration after sleep onset, rapid eye movement (REM) duration, total sleep duration) during the course of pandemic-related events and stay-at-home rules affecting residents in San Antonio, Texas and the surrounding areas (Figure 1). To identify whether pandemic-related events and/or stay-at-home rules altered daily behavior in residents, we focused on three key questions: (1) Which date saw the greatest inflection point in daily behaviors? (2) What is the demographic and health profile for individuals whose behavior (a) changed for the worse or (b) improved, during the stay-at-home orders? (3) Is self-evaluation an effective barometer of health during stay-at-home orders?

**Figure 1.**
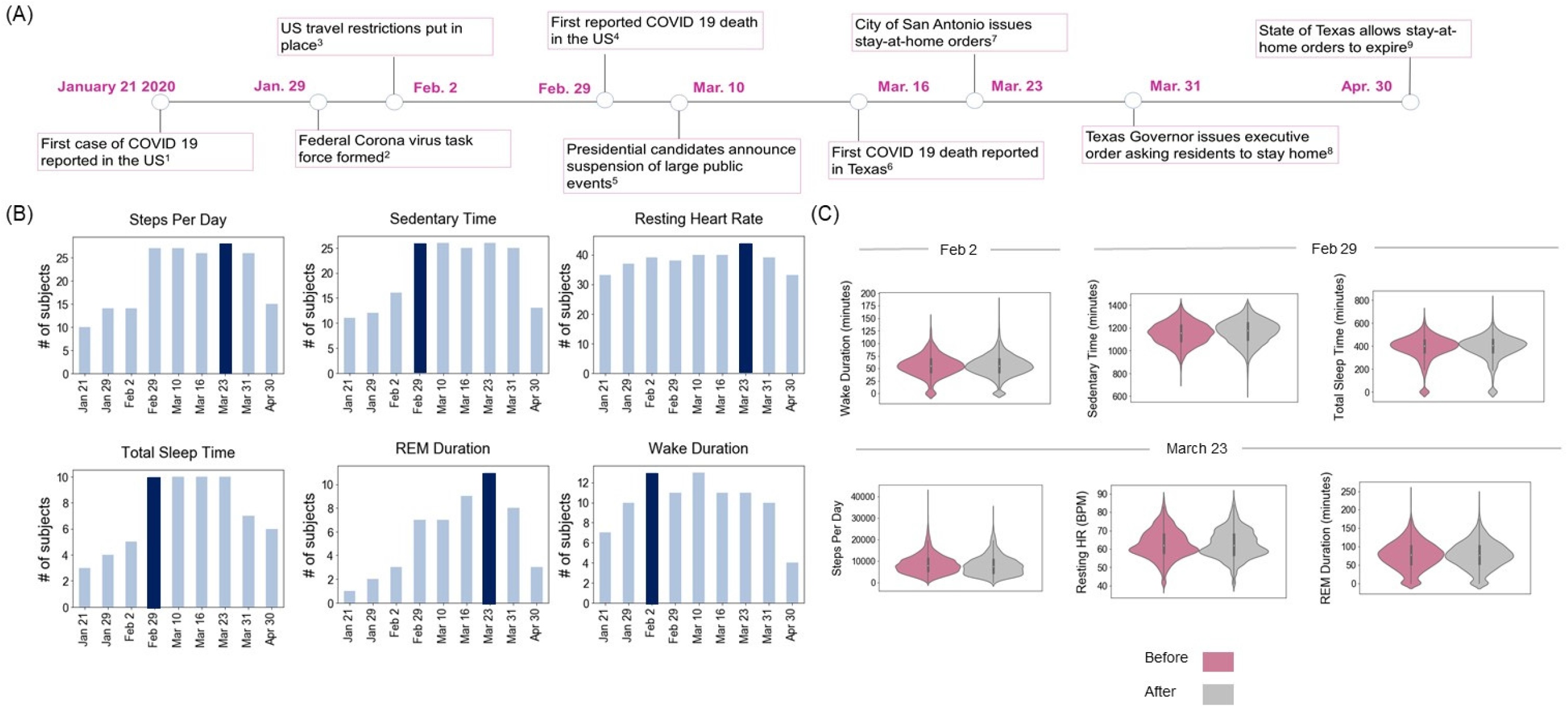
Events affecting daily activity during the COVID19 pandemic. **(A)** Timeline of prominent COVID 19 events relevant to the subjects in Texas, January 2020 to April 2020. **(B)** The number of subjects (of 61 total subjects) with statistically significant changes for six daily activity metrics (steps, sedentary time, resting heartrate, total sleep time, REM duration and wake duration) at those dates. (C) Violin plots showing the probability density of the six activity metrics for subjects before (magenta) and after (gray) the date for which most people had a significant change in the variable.

## Background

Government-mandated and individually-motivated lifestyle changes due to the COVID-19 pandemic have the potential to adversely affect the health of non-infected individuals. States of emergency induce population-wide mental distress^11^. The COVID-19 pandemic, along with epidemics such as SARS and MERS, have shown strong adverse effects on mental health in the uninfected, isolating population^12-14^. Bidirectional relationships between stress and sleep compound the effects. Anxiety decreases sleep quality in healthy individuals^15-18^, while lack of good-quality sleep exacerbates the cognitive stress caused by the pandemic^19^. Moreover, poor sleep quality decreases immunity to viral infection^20,21^. Like sleep, exercise also regulates immune response and mitigates stress^22^. Moderate exercise can decrease susceptibility to infections^23,24^, and has proven effective in ameliorating stress surrounding the COVID-19 pandemic^19^. Conversely, ceasing moderate exercise increases depressive symptoms^25^. As such, mandatory closure of gyms, fitness centers, or parks can impact exercise, stress relief, and overall health for the ∼40% of people relying on these facilities^26^.

However, knowledge about the effect of COVID-19 mitigation measures on sleep, exercise and other daily activity remains incomplete. First, the majority of activity studies have focused on the population in China, where the COVID-19 virus was first reported and early, stringent social measures were taken to combat its spread. Second, studies have primarily relied on surveys to determine self-reported sleep quality and activity levels^27^. Yet self-reported sleep quality often differs from sensor-tracked measurements and observation of sleep via polysomnography^28^. Moreover, most studies to date lack longitudinal measures of sleep quality and fitness. However, behavior would be expected to change over weeks and months, as individuals adapt to stresses of a pandemic.

With this in mind, we used wearable devices to gather data over the course of the COVID-19 pandemic in the United States. We focused on a cohort of 61 subjects in Texas, whose activity had been studied longitudinally for months prior to the pandemic. This allowed us test whether daily activity changes could be directly related to specific events in the progression of the pandemic. We focused on six metrics that reflect general health and that are indicative of long-term cognitive health^10,29,30^: sedentary time^31,32^, resting heart rate^33^, steps per day^34^, rapid-eye-movement (REM) duration^35^, wake duration during a sleep session, and total sleep duration^36,37^. To address our third aim (*identify whether self-reported behaviors are reliable barometers of health*), we analyzed subjects’ responses to a questionnaire and compared self-reported values to those recorded by activity tracking.

## Methods

125 adults (79 women, 46 men, age 56 ± 18.1 years) were recruited for a longitudinal study on daily behaviors and brain health between October 2017 and November 2019 (IRB Protocol 19-077R, Appendix C). Subjects were given a Fitbit Charge 3 activity tracker, which they were instructed to wear day and night. A questionnaire focusing on changes in activity levels, social interactions, sleep, and diet due to the stay-at-home orders caused by the COVID-19 pandemic was emailed to subjects and shared with the public between March 13 and June 10, 2020 (Appendix G).

We compiled activity tracker data for the months of August 2019 through August 2020 (Appendices A and D). Our analysis was restricted to subjects whose Fitbit device recorded ≥ 70% usage (by total possible minutes), for each 3-month period (Aug to Oct, Nov to Jan, Feb to Apr). This group (Group A) consisted of 40 women and 21 men (age, 59.0 ± 15.9). Within Group A, those who answered the stay-at-home order questionnaire were additionally analyzed (Group B: 34 women, 14 men, age 61.7 ± 14.6). Data analysis was performed using the Python Stats package and Shrinkage Clustering in R^38^.

### Determining Significant Dates

Figure 1 shows the timeline of nine target dates where pandemic-related events or announcements occurred that we hypothesized would affect subjects’ activities. For each target date, we performed a two-sided t-test for two independent samples for each individual in Group A comparing six selected activities (steps, sedentary active minutes, resting heart rate, REM duration, wake duration, and total sleep time) from (a) January 1, 2020 through the target date to (b) the target date through May 31.

### Assessing Activity Changes

For the dates where significant changes for each variable were observed, we calculated the mean values of each activity and determined their linear trend (*activity metric vs. time*) for every subject before and after that date (Figure 1, Appendix D). To identify activity patterns, we analyzed subjects in three ways: **(1)** by demographics (Figure 2A); **(2)** by grouping subjects whose mean increased/decreased for each activity before and after the target date (Figure 2B); and **(3)** by classification based on shared activity levels across all six metrics (Figure 3). For the last, Shrinkage Clustering^38^ identified the optimal number of groups and assigned subjects into clusters based on activity before and after each target date (Table D2, Appendices D-F). Clusters were ranked from best health changes to worst, and demographics for each cluster were analyzed. We also performed the aforementioned analysis for dates after the stay-at-home orders had lifted (June 1 to August 31), comparing activity to during stay-at-home orders (target date to May 31) (Figure 3). To account for normal variations in the same subjects, z-scores for each activity change were assessed in comparison to monthly changes for August to December 2019 (Table 1).

**Table 1.** Trends in daily activity during and after stay-at-home orders. Activity changes based on those subjects that increased or decreased each activity during or after stay-at-home orders. Target date = date where the most subjects had a significant change in activity. Mean change = average daily change in that activity for the entire cohort. Mean z-score = average number of standard deviations away from the month-to-month average change from the previous year. Maximum difference = values for the subject with the highest % difference between the average % change month-to-month in the previous year and their change after the target date. Activity changes generally considered positive for fitness and brain health are highlighted in green, while negative changes are in light red/pink. Values greater than 50% are highlighted in bold. A star (*) denotes significant change after the target date.

| Daily Activity | Target Date | Time Period Relative to SAMO | Mean Change (%) | Increase in Activity |  |  |  |  | Decrease in Activity |  |  |  |  |
| --- | --- | --- | --- | --- | --- | --- | --- | --- | --- | --- | --- | --- | --- |
|  |  |  |  | # of Subjects (%) | Overall Mean | Mean z-score | Maximum Difference |  | # of Subjects (%) | Overall Mean | Mean z-score | Maximum Difference |  |
|  |  |  |  |  |  |  | % | Average Daily Value |  |  |  | % | Average Daily Value |
| Wake Time | Feb 2 | Before-to-During | 2.0 ± 15.7 | **33 (54%)** | 9.8 ± 10.6 | 1.0 ± 1.3 | 34.6 | +17.4 min | 28 (46%) | -8.5 ± 5.6* | -1.2 ± 1.0 | -25.2 | -10.6 min |
|  |  | During-to-After | -1.4 ± 9.3 | 23 (45%) | 6.3 ± 3.7 | 0.8 ± 0.9 | 10.8 | +6.7 min | **28 (55%)** | -5.9 ± 4.7 | -1.0 ± 1.2 | -27.2 | -3.7 min |
| Sedentary Time | Feb 29 | Before-to-During | 1.2 ± 3.0 | **43 (70%)** | 2.7 ± 2.1* | 1.5 ± 1.5 | 10.9 | +116.7 min | 18 (30%) | -2.3 ± 1.6 | -0.9 ± 0.9 | -6.8 | -66.4 min |
|  |  | During-to-After | 0.8 ± 3.2 | **27 (53%)** | 2.9 ± 3.0 | 1.7 ± 1.6 | 12.1 | +135.0 min | 24 (47%) | -1.5 ± 1.4 | -0.6 ± 0.8 | -7.1 | -76.6 min |
| Total Sleep Time | Mar 23 | Before-to-During | -0.4 ± 6.1 | 25 (41%) | 4.4 ± 6.1 | 0.6 ± 1.0 | 29.1 | +103.6 min | **36 (59%)** | -3.8 ± 3.0 | -0.9 ± 1.0 | -18.4 | -33.9 min |
|  |  | During-to-After | -1.1 ± 4.8 | 23 (45%) | 3.1 ± 2.4 | 0.3 ± 0.7 | 9.7 | +32.7 min | **28 (55%)** | -4.5 ± 3.4 | -1.0 ± 1.3 | -14.5 | -59.0 min |
| Resting HR | Mar 23 | Before-to-During | -0.6 ± 3.5 | 22 (36%) | 3.1 ± 2.9 | 1.0 ± 1.2 | 11.7 | +7.0 bpm | **39 (64%)** | -2.7 ± 1.6 | -1.4 ± 1.8 | -7.8 | -3.7 bpm |
|  |  | During-to-After | 0.2 ± 3.6 | **28 (55%)** | 2.5 ± 2.7 | 1.0 ± 1.1 | 12.0 | +8.0 bpm | 23 (45%) | -2.6 ± 2.4 | -1.0 ± 0.9 | -13.7 | -6.9 bpm |
| REM | Mar 23 | Before-to-During | 2.2 ± 18.9 | 29 (48%) | 11.9 ± 13.1 | 0.9 ± 1.3 | **50.8** | +29.0 min | **32 (52%)** | -8.0 ± 7.0 | -1.0 ± 1.0 | -31.5 | -16.7 min |
|  |  | During-to-After | -5.9 ± 15.7 | 20 (39%) | 4.6 ± 3.5 | 0.3 ± 0.4 | 10.1 | +8.5 min | **31 (61%)** | -11.2 ± 14.8 | -0.8 ± 0.7 | **-100.6** | -4.1 min |
| Steps | Mar 23 | Before-to-During | -5.3 ± 20.0 | 22 (36%) | 14.3 ± 13.5 | 1.3 ± 1.4 | **52.6** | +1438.5 steps | **39 (64%)** | -16.5 ± 13.5* | -1.7 ± 1.4 | **-66.8** | -4618.1 steps |
|  |  | During-to-After | -1.7 ± 23.1 | 19 (37%) | 19.4 ± 20.2 | 1.2 ± 1.4 | **84.9** | +4173.8 steps | **32 (63%)** | -14.2 ± 13.9 | -1.8 ± 2.3 | **-57.3** | -6109.1 steps |

**Figure 2.**
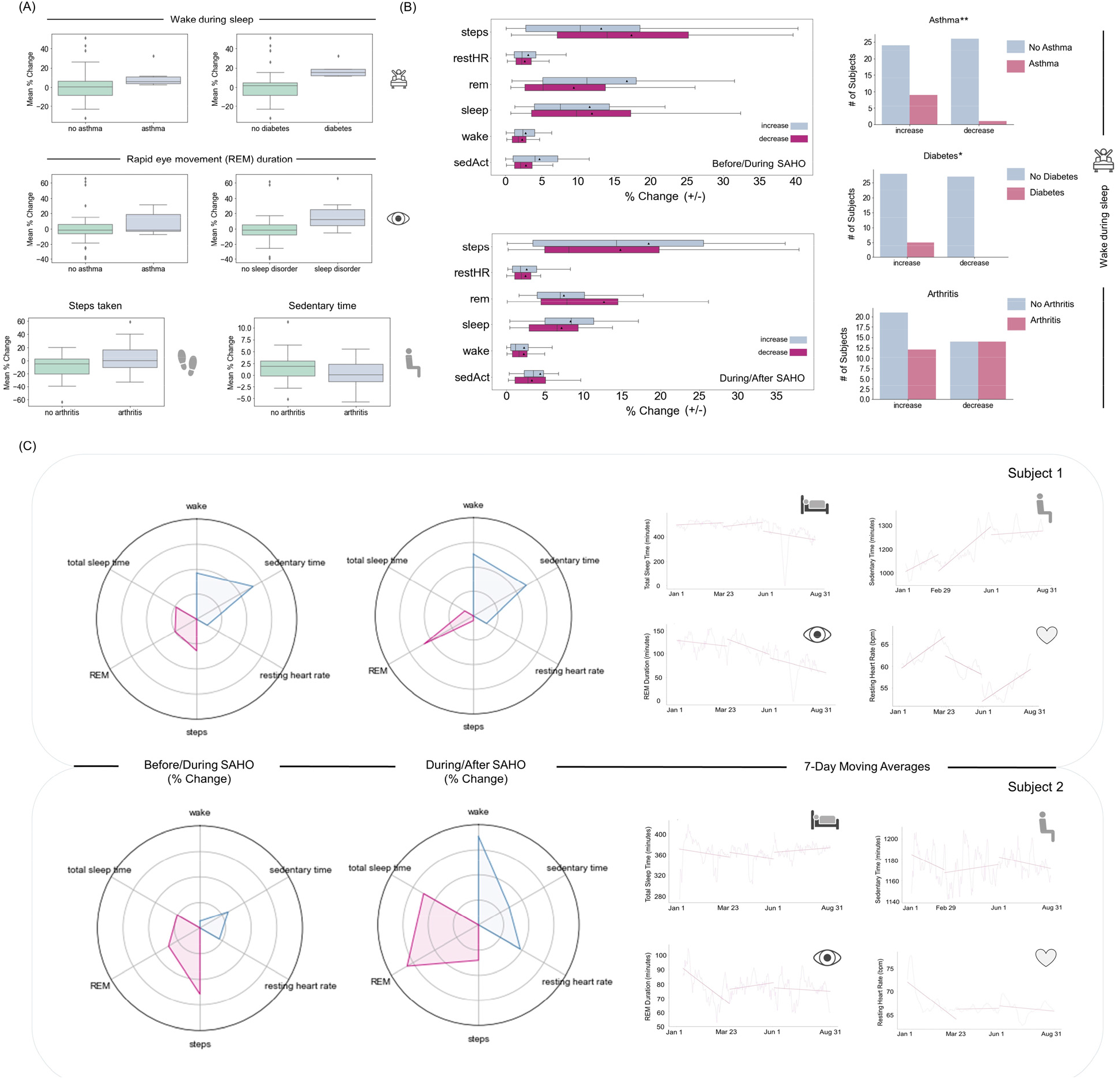
Activity changes during and after stay-at-home orders. **(A)** Population level trends in activity metrics during stay-at-home orders, grouped by demographics. Welch’s t-test, all displayed graphs p ≤ 0.05, except REM and asthma. **(B)** Population level trends grouped by subjects whose activity increased or decreased. Demographics are shown for the metric wake-during-sleep duration. Fischer test, **p ≤ 0.05, *p ≤ 0.01 **(C)** Representative trends before-to-during and during-to-after the stay-at-home orders shown for two subjects. Radar plots show the relative mean change (%) in daily activities, normalized by min-max normalization across the population. Activity trends that are positively correlated with fitness (i.e., an increase in steps walked) are shown in red. Activity trends that are negatively correlated with fitness (i.e., an increase in sedentary time) are in blue. 7-day moving averages of wake time, sedentary time, REM duration and resting heartrate for the same two subjects are shown on the right. Before/during = activity changes during stay-at-home orders (*target date to May 31*) were compared to before (*Jan 1 to the target date*). During/after: activity changes after stay-at-home orders lifted (*from June 1 to August 31, compared to the target date to May 31*). See Figures E1-E26, and Appendices D and E for full analysis.

**Figure 3.**
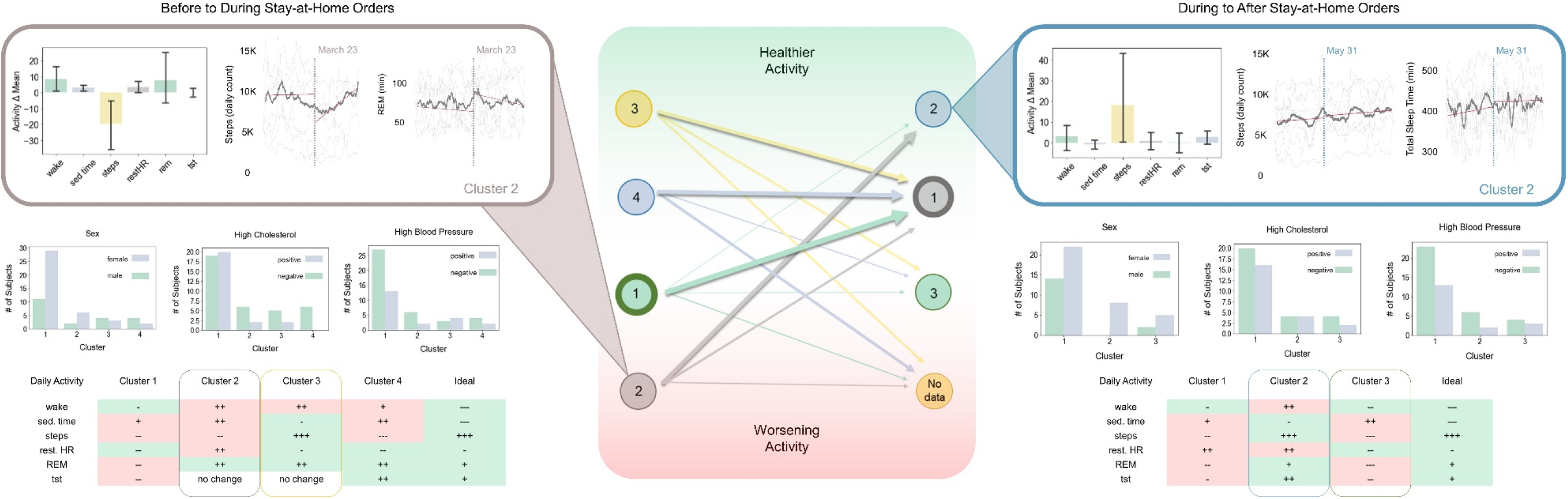
Activity Patterns Identified by Cluster Analysis. Four clusters before/during (left) and during/after (right) stay-at-home orders were identified. Shifts in cluster membership during these two time periods are shown in the center. The width of outlines and arrows represent the relative number of subjects in that group. Clusters are rank ordered by the healthiness of the observed activity trend. Representative activity patterns are displayed in the inset images for Cluster 2, which contained subjects with the worst health trends during stay-at-home orders (left inset), and Cluster 2B, which included subjects with the healthiest activity after stay-at-home orders were lifted (right inset). Select demographic and health data for shown for all clusters. See Appendices B, D, E and F for additional details.

### Determining Subjects’ Ability to Self-Evaluate Activity

Subjects’ answers to a COVID-19 questionnaire were compared to their digitally recorded activity (Figure 4, Tables D3-D4, Appendix G).

**Figure 4.**
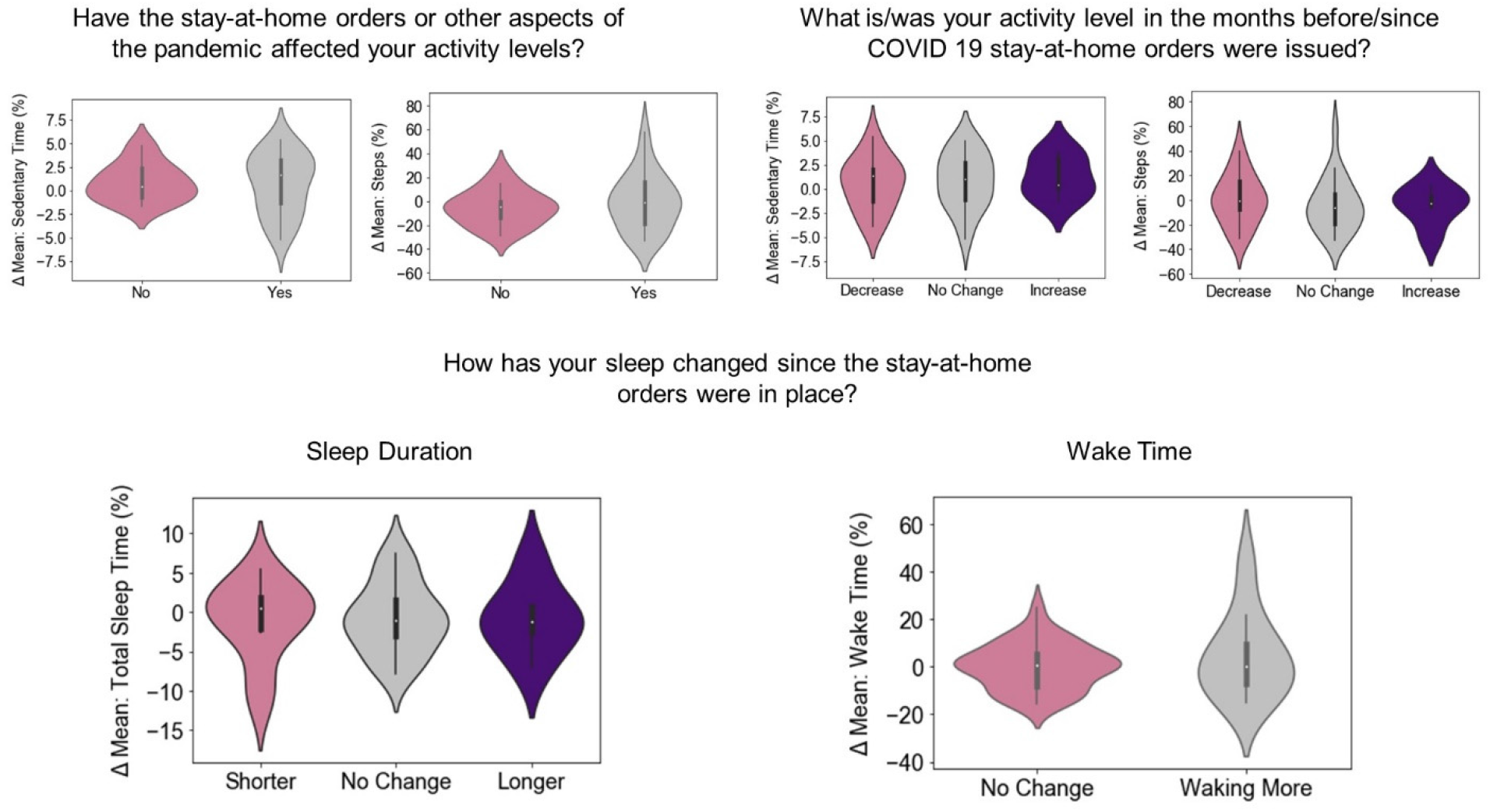
Quantitative vs. qualitative measures of daily activity. Subjects’ recorded activity levels (y-axis) compared to the same subjects’ responses (x-axis) to three survey questions (titles).

## Results

### Significant Dates

For each health metric, there was a significant difference before and after nine dates which marked milestones in the COVID-19 pandemic (Figure 1). Three metrics had the most subjects with significant changes on March 23, the day the city of San Antonio announced stay-at-orders: steps walked, resting heart rate, and REM duration. Wake duration had the highest number of subjects with significant change on February 2, the day the US announced international travel restrictions. The greatest change across the subjects in the duration of sedentary time and total sleep duration occurred on February 29, which coincided with the first reported death due to COVID 19 in the U.S.

### Activity Changes

**During stay-at-home orders**, the activity metric that changed the most across the population was steps walked daily, decreasing by an average of −5.3 ± 20.0% (Table 1A). When subjects were separated between those that increased or decreased activity, 70% increased the time they spent sedentary (2.7 ± 2.1%), 64% of subjects decreased the number of steps they walked daily (−16.5 ± 13.5%), and 36% saw an increase in their resting heart rate (3.1 ± 2.9%) during stay-at-home orders. 59% decreased total sleep time (−3.8 ± 3.0%) and 54% of subjects increased the time they spent awake each night (9.8 ± 10.6%).

**After stay-at-home orders** were lifted in San Antonio while the pandemic still raged, nightly REM duration (−5.9 ± 15.7%) changed the most across the population (Table 1B). 45% of subjects had an increase in wake time (6.3 ± 3.7% change); 53% increased their sedentary time (2.9 ± 3.0% change); and 55% saw an increase in resting heart rate (2.5 ± 2.7% change). 55% decreased total sleep time by −4.5 ± 3.4% and 63% of subjects decreased steps (−14.2 ± 13.9%).

Shrinkage Clustering identified groups of subjects by trends in their activity patterns. From daily activity metrics collected before and after the target date, four clusters were identified (Figure 3). 66% of subjects fell into Cluster 1 (61.2 ± 15.0 years old, 29 female, 11 male) with decreases in steps, REM, total sleep time and resting heart rate, and a slight increase in sedentary time during stay-at-home orders. The 13% of subjects in Cluster 2 (52.8 ± 17.9 years old, 6 female, 2 male) showed the worst health changes with increases in wake time, sedentary time, and REM and a substantial decrease in steps walked. Individuals in Cluster 3 (11% of subjects, 57.6 ± 20.8, 3 females, 4 males) showed the best health changes during stay-at-home orders, with a decrease in sedentary time and resting heart rate, and an increase in steps walked (Table D2).

After stay-at-home orders were lifted, three clusters of activity patterns emerged. 71% of subjects fell into a common cluster (Cluster 1B, 63.6 ± 12.5 years old, 22 females, 14 males) where steps, REM, and total sleep decreased, while sedentary time and resting heart rate increased. 16% of the subjects fell into Cluster 2B (50.6 ± 18.7 years old, 8 females, 0 males) which showed improvement in activities, e.g., increase in steps (18.2 ± 31.8%) (Table D2). 10 subjects stopped regularly wearing the fitness trackers, a potential indicator of waning commitment to fitness.

When we tracked shifts in cluster membership, 50% of subjects (all female) originally in Cluster 2 (the worst ranked cluster for that dataset) shifted to the cluster with the healthiest activity changes (Cluster 2B) after the stay-at-home orders lifted. Conversely, all subjects in Cluster 3, originally the best ranked cluster, dropped into worse performing clusters once stay-at-home orders lifted; 43% dropped into Cluster 1B, while the remaining 57% dropped into Cluster 3B or stopped recording their activity (Figure 3).

## Discussion

In San Antonio, Texas, home to subjects of this study, thousands of new SAR-CoV-2 infections are being reported daily. Six months after the expiration of statewide, 30-day stay-at-home order, San Antonio had a cumulative SARS2-CoV-2 infection rate of ∼1 in 24 people (*85,895 infected in a city of 2,005,409 individuals, 12*.*05*.*20, Source: City of San Antonio)*. Extending stay-at-home orders can be an effective, if contentious, infectious disease mitigation strategy. This work identifies whether unintended health consequences result^39^. We studied six metrics indicative of daily health and correlated to long-term cognitive health: steps walked per day, resting heart rate, sedentary minutes, time spent awake at night, total sleep time and REM duration^10,40^. Our goals were to understand in which ways stay-at-home orders affected activity, how individuals from different demographics were affected, and how accurately subjects were able to gauge the effect on their behaviors.

Nine key dates in the timeline of the SAR-CoV-2 pandemic in the U.S. were identified as possible instigators of behavioral changes (Figure 1). 93% of the study’s subjects live in the San Antonio metropolitan area, and hence we included the city’s dates for stay-at-home-orders. Significant changes in four of the six health metrics (steps per day, resting heart rate, total sleep time, REM duration) occurred in the most subjects before and after March 23, the day the city of San Antonio issued stay-at-home orders.

When each health metric was studied in more detail, a broader picture emerged of how the target dates affected behavior (Figure 2). 70% of subjects increased the amount of time they spent being sedentary from Feb 29 to May 31. The majority (64%) of subjects had a decrease in steps per day, while 36% saw an increase in resting heart rate. In total, most people in the study saw a decrease in activity and overall physical fitness, however the average changes appeared modest (Table 1). We observed a similar pattern in sleep, where more than half of the subjects (54%) increased the time spent awake during the night and decreased total sleep time (59%). Individuals with pre-existing conditions of asthma or diabetes were more likely to experience declines in sleep quality during stay-at-home orders (Figure 2, Appendix E). Consistently poorer sleep has been shown to negatively affect physical and mental fitness^21^, even when daily changes are small. Notably, there were extremes. One individual took 66.8% less steps on average each day during the pandemic compared to their month-to-month average in 2019 and early 2020. Another spent on average 31.5% less time in REM (min) per night. Given the length of the pandemic, these changes are alarming. Just two weeks of increased sedentary activity can reduce muscle mass and insulin sensitivity^9^. The elderly, pre-diabetic and diabetic populations may be particularly vulnerable to long-term consequences of these activity changes. Surprisingly, some subjects appeared to significantly benefit from stay-at-home orders. One individual increased their daily steps by 52.6%, and another increased their average nightly REM duration by 50.8%. 28 subjects saw a decrease in arousals (wake disruptions) during sleep, indicating that following the issuance of stay-at-home orders, they slept more soundly than normal (Table 1).

With an almost even divide between those that showed an improvement in health metrics and those that did not, we sought to understand how stay-at-home orders were affecting individuals differently. Cluster analysis revealed four patterns of activity (Figure 3). Individuals in Cluster 3 showed a decrease in sedentary time with an increase in steps (∼8000 steps/day to >10,000 steps/day) indicating an improvement in physical fitness. For most subjects in Cluster 3, this positive trend reversed after stay-at-home orders lifted (Appendices F and G). Notable demographics of Cluster 3 included that it was 57% male (4 of 7), and >50% of its members had arthritis (whereas the overall study population demographics are 34% male and 42% with arthritis). In contrast to the improvements seen in Cluster 3, subjects in Clusters 1, 2 and 4 consistently increased how much time they spent sedentary during stay-at-home orders, and all three groups took fewer daily steps on average. Subjects in Cluster 2 initially fared the worst. Before the pandemic, subjects in Cluster 2 were active (averaging ∼10,000 steps/day); at the onset of stay-at-home orders, their step count fell quickly to <6000-7000 steps/day and then slowly increased towards baseline (Figures 3 and F2).

Benefits of this study include a data-rich, 16-month quantitative analysis of daily activity before and during the pandemic. Passively recorded digital health metrics provided a critical addition to user-based surveys (Figure 4). Limitations of extrapolating this study across populations should also be noted. The cohort size was small and of a narrow demographic (61 people, age 56 ± 18.1 years, 54.8% retired, 100% in Texas). Subjects were specifically asked about stay-at-home orders; however, we cannot eliminate the possibility that unaccounted or combinatorial factors contributed to the observed activity changes.

In summary, this work has the potential to guide policy decision-making by illuminating how stay-at-home orders during a pandemic quantitatively affect the daily health of a population. An exciting, unexpected conclusion from this work is that there exists a subset of individuals whose health quantitatively improves – *better sleep, more exercise, lower resting heart rate* – in the midst of a devastating pandemic and the accompanying restrictions imposed by stay-at-home orders. Future work remains to understand in detail what mitigation methods are most effective in combating a pandemic while encouraging optimal health.

## Supporting information

Appendices A through G

## Data Availability

Data described and analyzed in the paper is provided and archived.

https://qutublab.org/

