## Appendices A through G for "Health Signatures During COVID-19: A Precision Fitness Case Study"

**Supplementary Material:**

**Appendices A-H**

Supplement to:

*Pollet EP, Sathish A, Maloney Z, Long BL, Brethen J, Qutub AA. Health Signature Changes During the COVID-19 Pandemic: A Precision Fitness Case Study.*

**Appendices Table of Contents**


**Appendix A. Overview of collected activity data**

**Figure A1.** Timeline of data collected by Fitbit over a 24-hour period.

**Table A1.** Table of the types of data collected for this project. Subjects in the study were provided with a Fitbit Charge 3 (device shown at right).

**Appendix B**. **Supplemental Methods**

**B1.** Linear regression models for trends in activity

**B2.** Metrics used for Shrinkage Clustering

**B3.** Parameters used in Shrinkage Clustering

**B4.** Citations

**Appendix C**. **IRB approval letter**

**Appendix D**. **Supplementary Data Tables**

- **Table D1. Analysis of all subjects.** Target date found to have the most subjects with significant differences for each activity. Average z-scores for month-to-month % change (Aug-Dec 2019). Before % changes compare activity changes from Jan 1 – target date and target date – May 31. After % changes compare activity changes from target date-May 31 and June 1 – Aug 31.

- **Table D2. Cluster Membership Data.** Comparing % change in activity before the target date and after the target date to May 31, 2020 for each cluster. (a) Comparing % change in activity after the target date to May 31 and June 1 – August 31, 2020 for each cluster. (b) Z-scores are comparing month-to-month changes from Aug – December of 2019.

- **Table D3. Pandemic survey answers and activity.** Answers to the survey question about exercise and activity levels compared to sedentary time and steps provided by Fitbit.

- **Table D4. Pandemic survey answers and sleep.** Total sleep time and wake time after sleep onset compared to answers from the Pandemic Survey.

**Appendix E. Demographic Figures -** * indicate $p<0.1$, ** $p<0.05$

- **Before/During Stay-at-Home Orders:**
  - **Figure E1.** Overall demographics and health data on all subjects included in before/during dataset. Subjects were given a questionnaire upon entering the study.
  - **Figure E2**. REM trend before/during SAHO by demographic and health data.
  - **Figure E3.** Step trend before/during SAHO by demographic and health data.
  - **Figure E4.** Resting heart rate trend before/during SAHO by demographic and health data.

- **Figure E5.** Wake trend before/during SAHO by demographic and health data.
- **Figure E6.** Sedentary active trend before/during SAHO by demographic and health data.
- **Figure E7.** Total sleep time trend before/during SAHO by demographic and health data.
- **Figure E8.** REM trend by demographics and health data before/during SAHO divided by those that increased or decreased activity.
- **Figure E9.** Step trend by demographics and health data before/during SAHO divided by those that increased or decreased activity.
- **Figure E10.** Resting heart rate trend by demographics and health data before/during SAHO divided by those that increased or decreased activity.
- **Figure E11.** Wake trend by demographics and health data before/during SAHO divided by those that increased or decreased activity.
- **Figure E12.** Sedentary active trend by demographics and health data before/during SAHO divided by those that increased or decreased activity.
- **Figure E13.** Total sleep time trend by demographics and health data before/during SAHO divided by those that increased or decreased activity.

❖ **During/After Stay-at-Home Orders:**

- **Figure E14.** Overall demographics and health data on all subjects included in during/after dataset. Subjects were given a questionnaire upon entering the study.
- **Figure E15.** REM trend during/after SAHO by demographic and health data.
- **Figure E16.** Step trend during/after SAHO by demographic and health data.
- **Figure E17.** Resting heart rate trend during/after SAHO by demographic and health data.
- **Figure E18.** Wake trend during/after SAHO by demographic and health data.
- **Figure E19.** Sedentary active trend during/after SAHO by demographic and health data.
- **Figure E20.** Total sleep time trend during/after SAHO by demographic and health data.
- **Figure E21.** REM trend by demographics and health data during/after SAHO divided by those that increased or decreased activity.
- **Figure E22.** Step trend by demographics and health data during/after SAHO divided by those that increased or decreased activity.
- **Figure E23.** Resting heart rate trend by demographics and health data during/after SAHO divided by those that increased or decreased activity.
- **Figure E24.** Wake trend by demographics and health data during/after SAHO divided by those that increased or decreased activity.
- **Figure E25.** Sedentary active trend by demographics and health data during/after SAHO divided by those that increased or decreased activity.
- **Figure E26.** Total sleep time trend by demographics and health data during/after SAHO divided by those that increased or decreased activity.

❖ **Cluster Membership Demographics**

- **Figure E27.** Demographic and health graphs for clusters found by Shrinkage Clustering method from the dataset containing the % change in means and slope of the linear trendlines comparing before/during the SAHO.
- **Figure E28.** Mean % change (left) and % change of slope of the linear trendline (right) for before/during the SAHO by activity.
- **Figure E29.** Mean % change (left) and % change of slope of the linear trendline (right) for before/during the SAHO by cluster.

- **Figure E30.** Demographic and health graphs for clusters found by Shrinkage Clustering method from the dataset containing the % change in means and slope of the linear trendlines comparing during/after the SAHO.
- **Figure E31.** Mean % change (left) and % change of slope of the linear trendline (right) for during/after the SAHO by activity.
- **Figure E32.** Mean % change (left) and % change of slope of the linear trendline (right) for during/after the SAHO by cluster.

**Appendix F. Highlighted Activity Trends by Cluster**

- **Figure F1.** Sedentary active minutes (a) and steps (b) for cluster 1 before/during SAHO.
- **Figure F2.** Steps for cluster 2 before/during SAHO.
- **Figure F3.** Sedentary active minutes (a) and steps (b) for cluster 3 before/during SAHO.
- **Figure F4.** Sedentary active minutes (a) and REM duration (b) for cluster 4 before/during SAHO.
- **Figure F5.** Steps for cluster 2 during/after SAHO.
- **Figure F6.** REM duration (a), steps (b), and wake duration (c) for cluster 3 during/after SAHO.

**Appendix G**. **Self-evaluation of activity.** Survey provided to participants and the general public via email (B1) and results from the survey (B2)

**Appendix H. Fitbit Data File.**

**Appendix A.** Overview of collected activity data

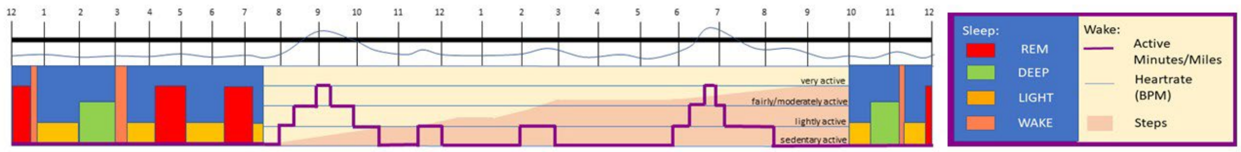


**Figure A1.** Timeline of data collected by Fitbit over a 24-hour period.

| Heart Rate | Activity | Sleep | Surveys |
|---|---|---|---|
| Resting HR | Steps | Total Sleep Duration | Demographic |
| Average HR | Activity Level Minutes | Nap Duration and Nap Count | Health |
| Max HR | Activity Level Miles | Individual Sleep Stage Duration and % | Activity During the Pandemic |
| Time of Max HR | | Bedtime | |
| | | Number of Sleep Stage Transitions | |

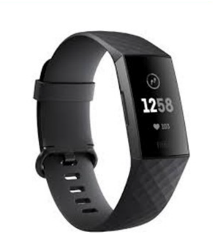

**Table A1.** Table of the types of data collected for this project. Subjects in the study were provided with a Fitbit Charge 3 (device shown at right).

**Appendix B**. Supplemental Methods

#### B1. Statistical Tests

| Figure | Statistical Test | Implementation | Parameters |
|---|---|---|---|
| Figure 1B | Two-sided t-test for independent samples | SciPy.stats | Equal_var: True<br>Nan_policy: 'omit' |
| Figure 2A/Appendix E | Two-sided t-test for independent samples | SciPy.stats | Equal_var: False (Welch's t-test) |
| Figure 2B/Appendix E | Fisher Exact test | SciPy.stats | Alternative: 'two-sided' |

#### B2. Linear regression models for trends in activity

The slope of the linear trendline was defined as:

$$\frac{\Delta\, A}{\Delta\, time} = \frac{A_0 - A_i}{t_0 - t_i},$$

$$where\ A = activity\ metric, 0 = original\ date, i = \ subsequent\ date$$

For example, more subjects had a significant change in average REM duration per night starting on March 23 than other dates. We compared the mean REM duration per night for all individuals from Jan 1 to March 23 and March 23 to May 31. To assess trends in REM, we also calculated the slopes of the linear regression model for daily REM duration from Jan 1 to March 23, and March 23 to May 1. Analogous calculations were performed for each of the six activity metrics.

- Δ Slope Wake Time (%)[1,2]
- Δ Slope Sedentary Time (%)[3,4]
- Δ Slope Steps (%)[5]
- Δ Slope Resting Heart Rate (%)[6,7]
- Δ Slope REM Time (%)[8]
- Δ Slope Total Sleep Time (%)[1,2]

#### B3. Metrics used for Shrinkage Clustering

- Δ Slope Wake Time (%)[1,2]
- Δ Slope Sedentary Time (%)[3,4]
- Δ Slope Steps (%)[5]
- Δ Slope Resting Heart Rate (%)[6,7]
- Δ Slope REM Time (%)[8]
- Δ Slope Total Sleep Time (%)[1,2]
- Δ Mean Wake Time (%)
- Δ Mean Sedentary Time (%)
- Δ Mean Steps (%)
- Δ Mean Resting Heart Rate (%)
- Δ Mean REM Time (%)
- Δ Mean Total Sleep Time (%)

**B4. Parameters used in Shrinkage Clustering**

| Analysis | Method | Normalization | Implementation | Parameters |
|---|---|---|---|---|
| Cluster Analysis | Shrinkage Clustering[9] | Min-Max Normalization:<br><br>*(v-min)/(max-min)* | R Studio | s = simiMatrix<br>w = 5<br>k = 20<br>iter = 500<br>random = 1 |

| **Approval** | | |
|---|---|---|
| **Document No.:** | **Date:** | **Page:** |
| **HRP-522** | **26 June 2019** | **Page 1 of 1** |

Amina Ann Qutub, Ph.D.
Engineering(COE) - Biomedical Engineering
210-458-7092


Dear Principal Investigator:

On June 26, 2019 the IRB approved the following:

| | |
|---|---|
| Type of review: | **Modification – Repository & study** |
| Title: | **Profiling Cognitive Changes: Cells to Systems (Quantu Project)** |
| Principal investigator: | **Amina Ann Qutub, Ph.D.** |
| IRB number: | **19-077R** |
| Grant title and ID, if any: | **UTSA start-up (DGE045); NSF (pending)** |
| Documents reviewed: | **Modification Application; Research Personnel; Protocol (2) – Main & Repository; Consents (2) – Main & Repository;** |

*No later than one month prior to your approval anniversary date, you are to respond to the IRB annual check-in e-mail to request continuing oversight, or closure.* This e-mail will be sent to you approximately six weeks prior to the approval anniversary date of 0 4/11/2020.

Copies of any approved consent documents, consent scripts, or assent documents are attached.

When you have completed all research activities for the above listed study, please notify our office to close the study.

In conducting this study, you are required to follow the requirements in "INVESTIGATOR GUIDANCE: Investigator Obligations (HRP-800)."

Sincerely,

Tammy Lopez, J.D., C.I.P

Tammy Lopez, J.D., C.I.P.
IRB Member
Designee of the Chair
Senior Research Compliance Coordinator
UTSA Office of Research Integrity - IRB Office
GSR Rm.2.104N/P

210-458-6473

**Appendix D:** Supplemental Tables

**Table D1. Analysis of all subjects' activity**

**Table D2. Cluster membership data**

**Table D3. Pandemic survey answers and activity**

**Table D4. Pandemic survey answers and sleep**

| | | | | | Before | | | After | | |
|---|---|---|---|---|---|---|---|---|---|---|
| Activity | Target Date | Average Z-Score (%) | Min Z-Score (%) | Max Z-Score (%) | Average % Change | Min % Change | Max % Change | Average % Change | Min % Change | Max % Change |
| Wake Time | Feb 2 | 0.7 ± 3.3 | -14.4 | 6.9 | 2.0 ± 15.7 | -32.4 | 50.9 | -1.4 ± 9.3 | -31.1 | 16.8 |
| Sedentary Time | Feb 29 | -0.04 ± 0.9 | -1.7 | 2.2 | 1.2 ± 3.0 | -5.8 | 11.2 | 0.8 ± 3.2 | -6.3 | 12.8 |
| TST | Feb 29 | 0.7 ± 1.6 | -3.1 | 4.5 | -0.4 ± 6.1 | -11.5 | 30.5 | -1.1 ± 4.8 | -14.0 | 10.9 |
| Resting Heart Rate | Mar 23 | 0.3 ± 1.0 | -3.3 | 2.9 | -0.6 ± 3.5 | -6.9 | 11.6 | 0.2 ± 3.6 | -10.8 | 12.2 |
| REM | Mar 23 | 0.8 ± 4.5 | -23.2 | 9.9 | 2.2 ± 18.9 | -39.2 | 65.7 | -5.9 ± 15.7 | -83.6 | 17.8 |
| Steps | Mar 23 | 1.7 ± 4.9 | -9.0 | 17.9 | -5.3 ± 20.0 | -63.7 | 58.7 | -1.7 ± 23.1 | -54.8 | 88.7 |

**Table D1. Analysis of all subjects' activity.** Target date found to have the most subjects with significant differences for each activity. Average z-scores for month-to-month % change (Aug-Dec 2019). Before % changes compare activity changes from Jan 1 – target date and target date – May 31. After % changes compare activity changes from target date-May 31 and June 1 – Aug 31.

| | | Cluster 1 | | | | | Cluster 2 | | | | | Cluster 3 | | | | | Cluster 4 | | | | |
|---|---|---|---|---|---|---|---|---|---|---|---|---|---|---|---|---|---|---|---|---|---|
| Daily Activity | Target Date | | Overall Mean | | Mean z-score | | | Overall Mean | | Mean z-score | | | Overall Mean | | Mean z-score | | | Overall Mean | | Mean z-score | |
| | | # of Subjects | % | std dev | mean | std dev | # of Subjects | % | std dev | mean | std dev | # of Subjects | % | std dev | mean | std dev | #of Subjects | % | std dev | mean | std dev |
| Wake | 2-Feb | 40 | -1.4 | 8.9 | -0.1 | 1.6 | 8 | 8.3 | 11.1 | 0.8 | 1.5 | 7 | 8.9 | 16.0 | -0.1 | 1.9 | 6 | 1.7 | 20.9 | -0.1 | 1.4 |
| Sed Time | 29-Feb | 40 | 1.1 | 1.9 | 0.7 | 1.5 | 8 | 3.0 | 2.6 | 1.7 | 1.5 | 7 | -3.1 | 2.6 | -1.4 | 1.3 | 6 | 4.4 | 3.7 | 2.5 | 1.8 |
| Steps | 23-Mar | 40 | -5.6 | 15.1 | -0.6 | 1.5 | 8 | -19.5 | 21.4 | -1.6 | 1.4 | 7 | 23.1 | 19.1 | 2.0 | 1.8 | 6 | -18.1 | 12.4 | -2.6 | 2.1 |
| Resting HR | 23-Mar | 40 | -1.0 | 2.6 | -0.8 | 2.1 | 8 | 3.5 | 5.1 | 1.0 | 1.9 | 7 | -1.3 | 3.5 | -0.5 | 1.2 | 6 | -1.9 | 2.6 | -0.5 | 1.2 |
| REM | 23-Mar | 40 | -1.2 | 9.7 | -0.4 | 1.2 | 8 | 7.8 | 23.0 | 0.4 | 0.9 | 7 | 5.7 | 19.6 | 0.2 | 1.1 | 6 | 6.0 | 13.3 | 1.4 | 2.8 |
| TST | 29-Feb | 40 | -1.2 | 4.5 | -0.4 | 1.0 | 8 | 0.0 | 3.9 | -0.2 | 1.2 | 7 | 0.0 | 4.8 | -0.2 | 0.8 | 6 | 3.1 | 13.3 | 0.3 | 2.3 |

| | | Cluster 1 | | | | | Cluster 2 | | | | | Cluster 3 | | | | |
|---|---|---|---|---|---|---|---|---|---|---|---|---|---|---|---|---|
| Daily Activity | Target Date | | Overall Mean | | Mean z-score | | | Overall Mean | | Mean z-score | | | Overall Mean | | Mean z-score | |
| | | # of Subjects | % | std dev | mean | std dev | # of Subjects | % | std dev | mean | std dev | # of Subjects | % | std dev | mean | std dev |
| Wake | 2-Feb | 36 | -0.4 | 6.6 | -0.3 | 1.6 | 8 | 3.2 | 8.6 | 0.1 | 0.8 | 7 | -4.6 | 7.7 | -0.4 | 0.7 |
| Sed Time | 29-Feb | 36 | 0.3 | 2.0 | 0.3 | 1.2 | 8 | -0.7 | 3.2 | 0.8 | 3.1 | 7 | 5.1 | 4.7 | 1.8 | 1.5 |
| Steps | 23-Mar | 36 | -1.1 | 15.6 | -0.4 | 1.6 | 8 | 18.2 | 31.8 | 0.6 | 2.2 | 7 | -27.6 | 19.4 | -3.7 | 3.8 |
| Resting HR | 23-Mar | 36 | 0.2 | 2.7 | 0.0 | 1.1 | 8 | 0.9 | 5.9 | 0.6 | 2.4 | 7 | -0.6 | 4.0 | 0.1 | 1.1 |
| REM | 23-Mar | 36 | -3.4 | 8.5 | -0.3 | 0.8 | 8 | 0.2 | 7.2 | -0.1 | 0.6 | 7 | -19.5 | 27.5 | -1.0 | 1.0 |
| TST | 29-Feb | 36 | -1.3 | 3.6 | -0.3 | 0.9 | 8 | 2.8 | 4.7 | 0.0 | 0.8 | 7 | -4.8 | 6.8 | -1.4 | 2.3 |

**Table D2. Cluster membership data. (Top)** Comparing % change in activity before the target date and after the target date to May 31, 2020 for each cluster. **(Bottom)** Comparing % change in activity after the target date to May 31 and June 1 – August 31, 2020 for each cluster. Z-scores are relative to month-to-month changes from Aug – December of 2019.

| How has your sleep changed since the stay-at-home orders were in place? | | | | | | | |
|---|---|---|---|---|---|---|---|
| **Sleep Duration** | | | | | | | |
| | | Shorter | | No Change | | Longer | |
| Activity | Change in Average | # of subjects | Average % Change | # of subjects | Average % Change | # of subjects | Average % Change |
| Total Sleep Time | ↑ | 6 | 2.2 ± 1.8 | 8 | 4.0 ± 2.7 | 2 | 3.8 ± 4.0 |
| | ↓ | 5 | -5.1 ± 4.6 | 14 | -3.4 ± 2.6 | 4 | -3.0 ± 2.9 |
| **Wake Duration** | | | | | | | |
| | | Waking Less | | No Change | | Waking More | |
| Activity | Change in Average | # of subjects | Average % Change | # of subjects | Average % Change | # of subjects | Average % Change |
| Wake Time | ↑ | 0 | -- | 18 | 6.4 ± 6.6 | 5 | 17.0 ± 16.6 |
| | ↓ | 0 | -- | 12 | -8.9 ± 5.2 | 5 | -8.3 ± 5.4 |

**Table D3. Pandemic survey answers and activity.** Answers to the survey question about exercise and activity levels compared to sedentary time and steps provided by Fitbit.

| Activity | Change in Average | Have the stay-at-home orders or other aspects of the pandemic affected your activity levels? | | | | What is/was your activity level in the months before/since COVID 19 stay-at-home orders were issued? | | | | | |
|---|---|---|---|---|---|---|---|---|---|---|---|
| | | Yes | | No | | Decrease | | No Change | | Increase | |
| | | # of subjects | Average % Change | # of subjects | Average % Change | # of subjects | Average % Change | # of subjects | Average % Change | # of subjects | Average % Change |
| Sedentary Time | ↑ | 14 | 2.9 ± 1.5 | 12 | 1.8 ± 1.7 | 8 | 2.3 ± 1.6 | 14 | 2.6 ± 1.7 | 4 | 1.9 ± 1.9 |
| | ↓ | 9 | -2.7 ± 1.6 | 5 | -1.2 ± 0.5 | 5 | -2.3 ± 1.4 | 8 | -2.2 ± 1.6 | 1 | -1.3 |
| Steps | ↑ | 11 | 18.0 ± 18.0 | 4 | 14.9 ± 8.2 | 6 | 16.6 ± 14.7 | 7 | 20.4 ± 18.6 | 2 | 7.6 ± 6.5 |
| | ↓ | 12 | -17.8 ± 12.4 | 13 | -11.2 ± 9.9 | 7 | -13.5 ± 12.4 | 15 | -15.2 ± 11.1 | 3 | -12.6 ± 15.3 |

**Table D4. Pandemic survey answers and sleep.** Total sleep time and wake time after sleep onset compared to answers from the Pandemic Survey.

#### Appendix E. Demographic Figures

** denotes $p < 0.05$, * denotes $p < 0.1$

- **Before/During Stay-at-Home Orders:**
  - **Figure E1.** Overall demographics and health data on all subjects included in before/during dataset. Subjects were given a questionnaire upon entering the study.
  - **Figure E2**. REM trend before/during SAHO by demographic and health data.
  - **Figure E3.** Step trend before/during SAHO by demographic and health data.
  - **Figure E4.** Resting heart rate trend before/during SAHO by demographic and health data.
  - **Figure E5.** Wake trend before/during SAHO by demographic and health data.
  - **Figure E6.** Sedentary active trend before/during SAHO by demographic and health data.
  - **Figure E7.** Total sleep time trend before/during SAHO by demographic and health data.
  - **Figure E8.** REM trend by demographics and health data before/during SAHO divided by those that increased or decreased activity.
  - **Figure E9.** Step trend by demographics and health data before/during SAHO divided by those that increased or decreased activity.
  - **Figure E10.** Resting heart rate trend by demographics and health data before/during SAHO divided by those that increased or decreased activity.
  - **Figure E11.** Wake trend by demographics and health data before/during SAHO divided by those that increased or decreased activity.
  - **Figure E12.** Sedentary active trend by demographics and health data before/during SAHO divided by those that increased or decreased activity.
  - **Figure E13.** Total sleep time trend by demographics and health data before/during SAHO divided by those that increased or decreased activity.

- **During/After Stay-at-Home Orders:**
  - **Figure E14.** Overall demographics and health data on all subjects included in during/after dataset. Subjects were given a questionnaire upon entering the study.
  - **Figure E15.** REM trend during/after SAHO by demographic and health data.
  - **Figure E16.** Step trend during/after SAHO by demographic and health data.
  - **Figure E17.** Resting heart rate trend during/after SAHO by demographic and health data.
  - **Figure E18.** Wake trend during/after SAHO by demographic and health data.
  - **Figure E19.** Sedentary active trend during/after SAHO by demographic and health data.
  - **Figure E20.** Total sleep time trend during/after SAHO by demographic and health data.
  - **Figure E21.** REM trend by demographics and health data during/after SAHO divided by those that increased or decreased activity.
  - **Figure E22.** Step trend by demographics and health data during/after SAHO divided by those that increased or decreased activity.
  - **Figure E23.** Resting heart rate trend by demographics and health data during/after SAHO divided by those that increased or decreased activity.
  - **Figure E24.** Wake trend by demographics and health data during/after SAHO divided by those that increased or decreased activity.
  - **Figure E25.** Sedentary active trend by demographics and health data during/after SAHO divided by those that increased or decreased activity.
  - **Figure E26.** Total sleep time trend by demographics and health data during/after SAHO divided by those that increased or decreased activity.

- **Cluster Membership Demographics**
  - **Figure E27.** Demographic and health graphs for clusters found by Shrinkage Clustering method from the dataset containing the % change in means and slope of the linear trendlines comparing before/during the SAHO.
  - **Figure E28.** Mean % change (left) and % change of slope of the linear trendline (right) for before/during the SAHO by activity.
  - **Figure E29.** Mean % change (left) and % change of slope of the linear trendline (right) for before/during the SAHO by cluster.
  - **Figure E30.** Demographic and health graphs for clusters found by Shrinkage Clustering method from the dataset containing the % change in means and slope of the linear trendlines comparing during/after the SAHO.
  - **Figure E31.** Mean % change (left) and % change of slope of the linear trendline (right) for during/after the SAHO by activity.
  - **Figure E32.** Mean % change (left) and % change of slope of the linear trendline (right) for during/after the SAHO by cluster.

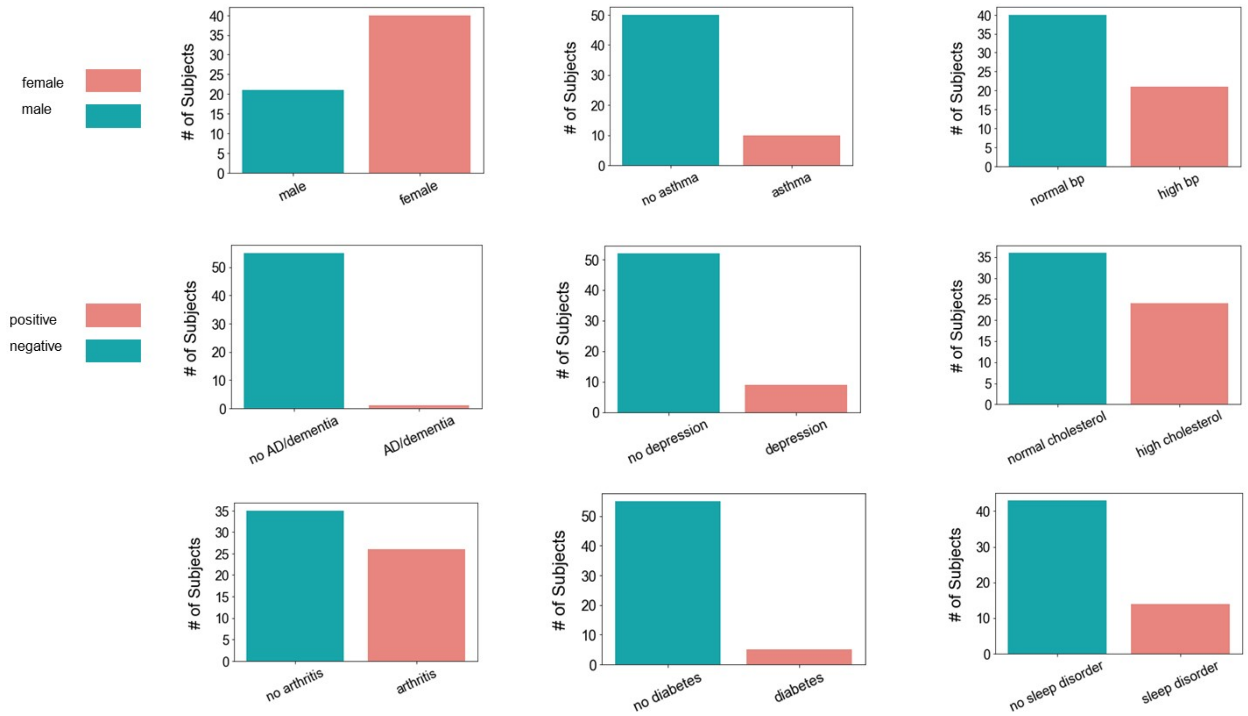


**Figure E1.** Overall demographics and health data on all subjects included in the before/during dataset. Subjects were given a questionnaire upon entering the study.

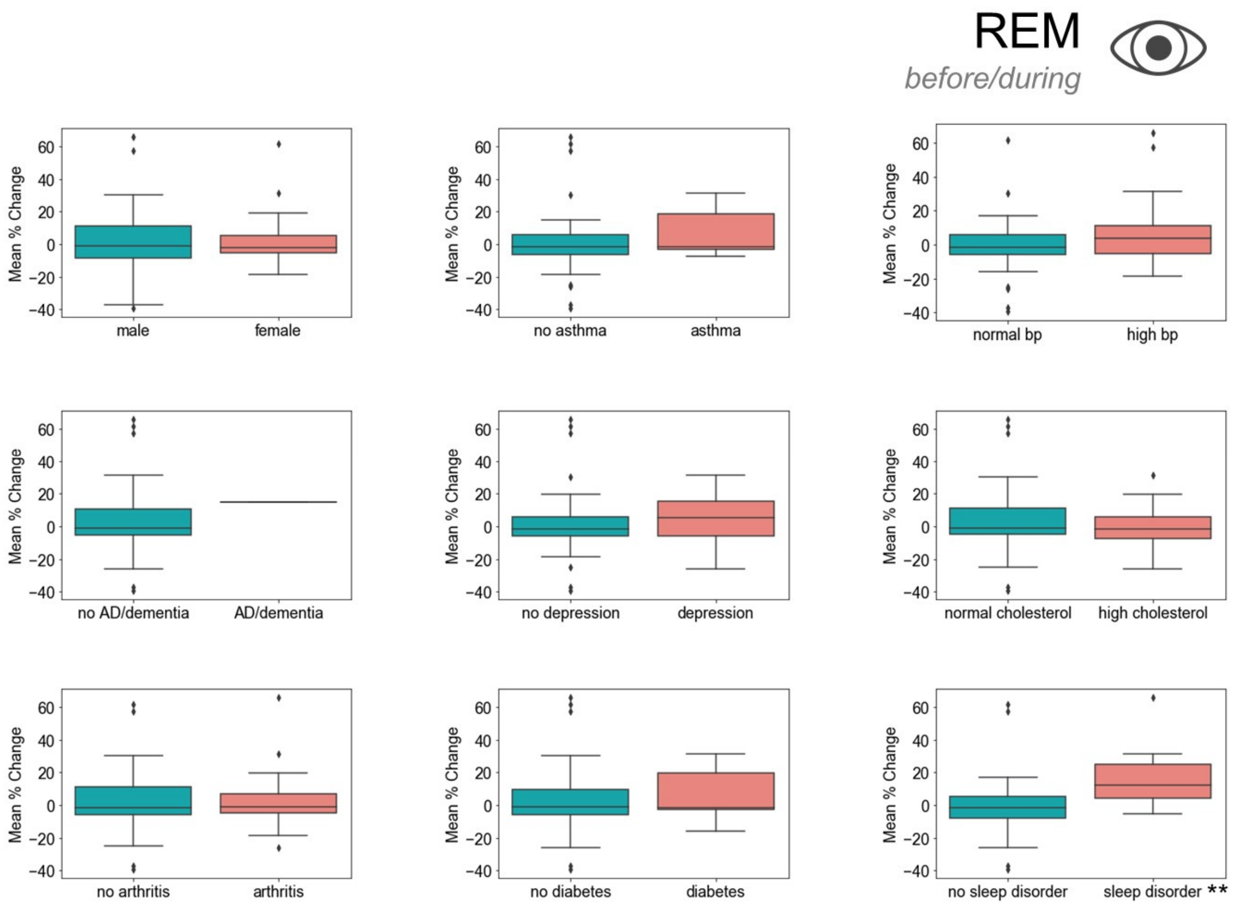


**Figure E2**. REM trend before/during SAHO by demographic and health data.

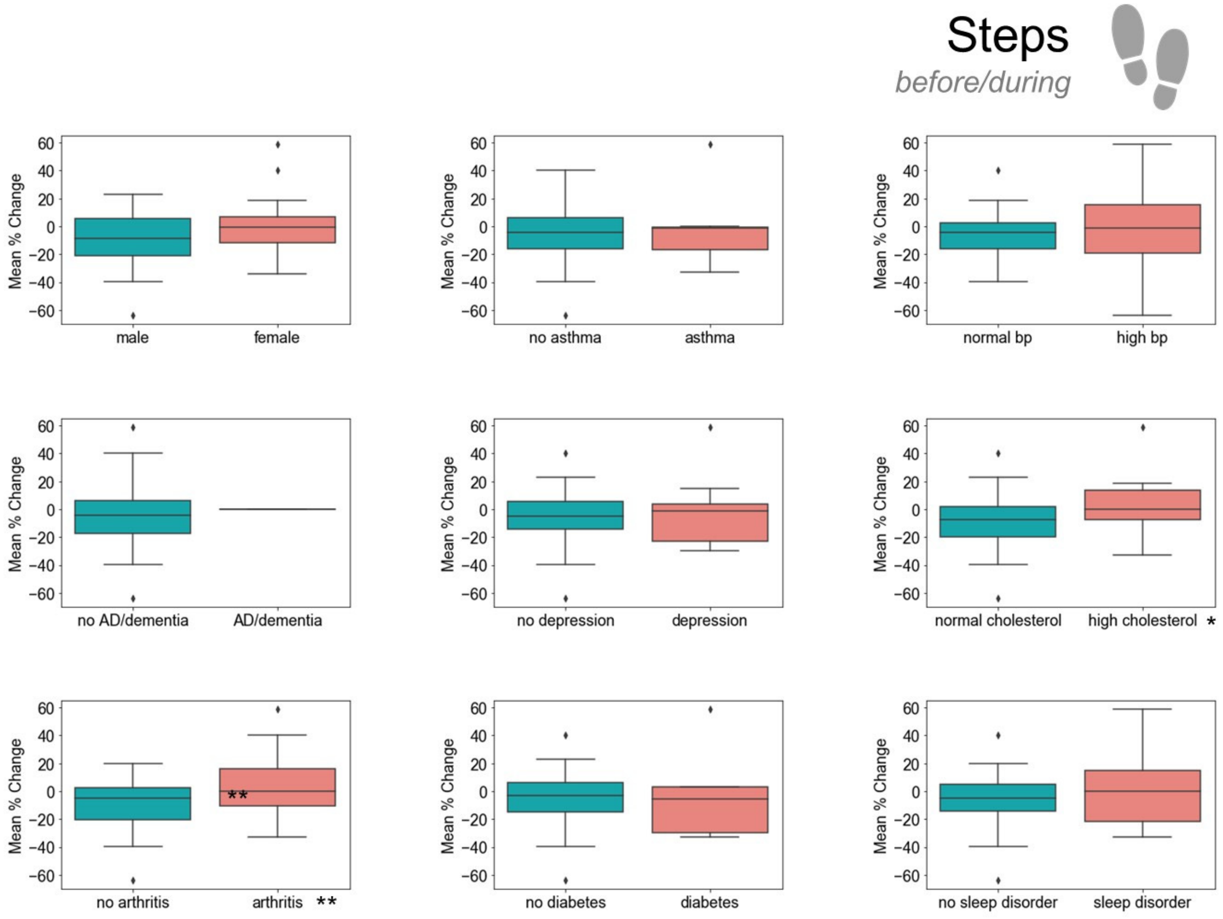


**Figure E3.** Step trend before/during SAHO by demographic and health data.

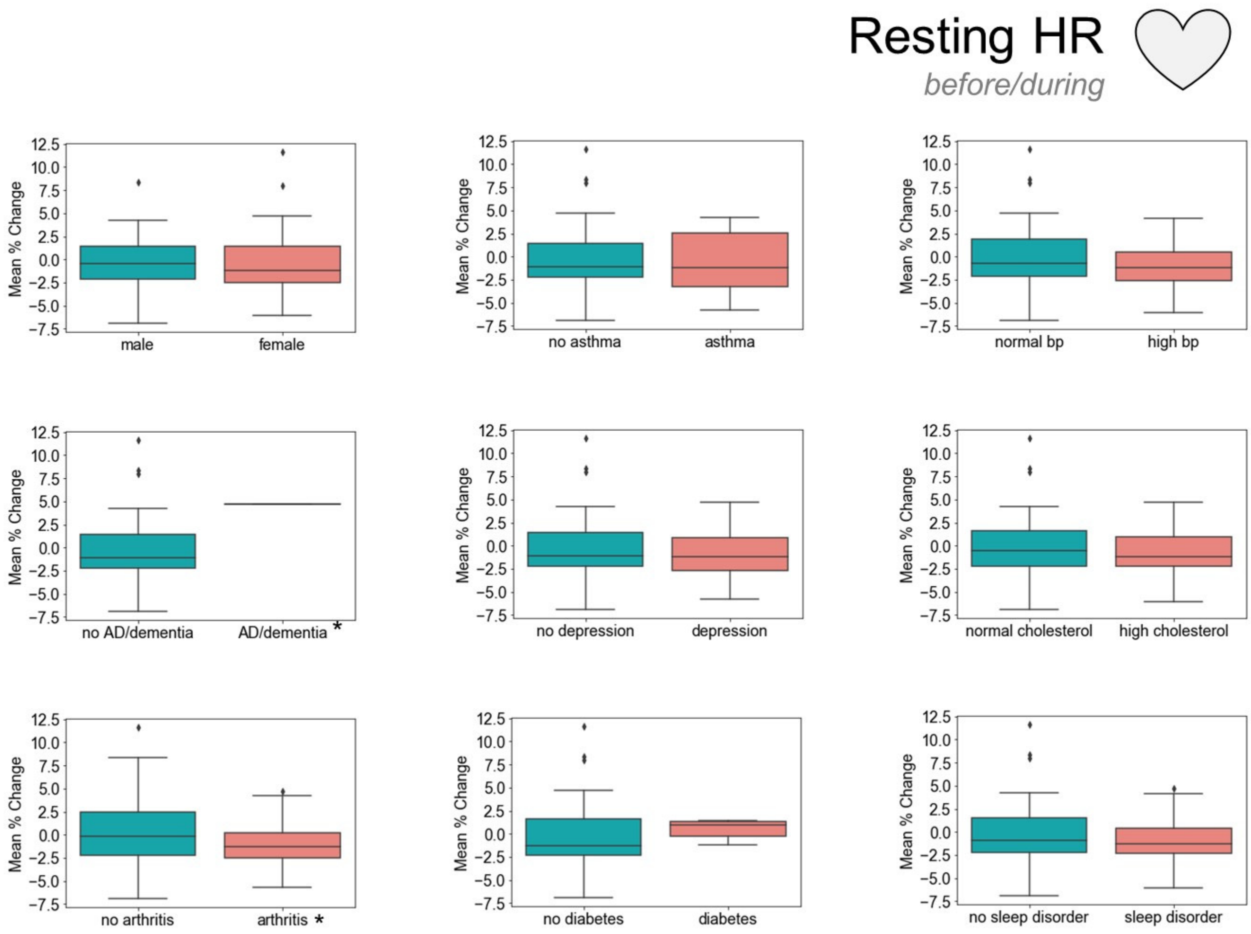


**Figure E4.** Resting heart rate trend before/during SAHO by demographic and health data.

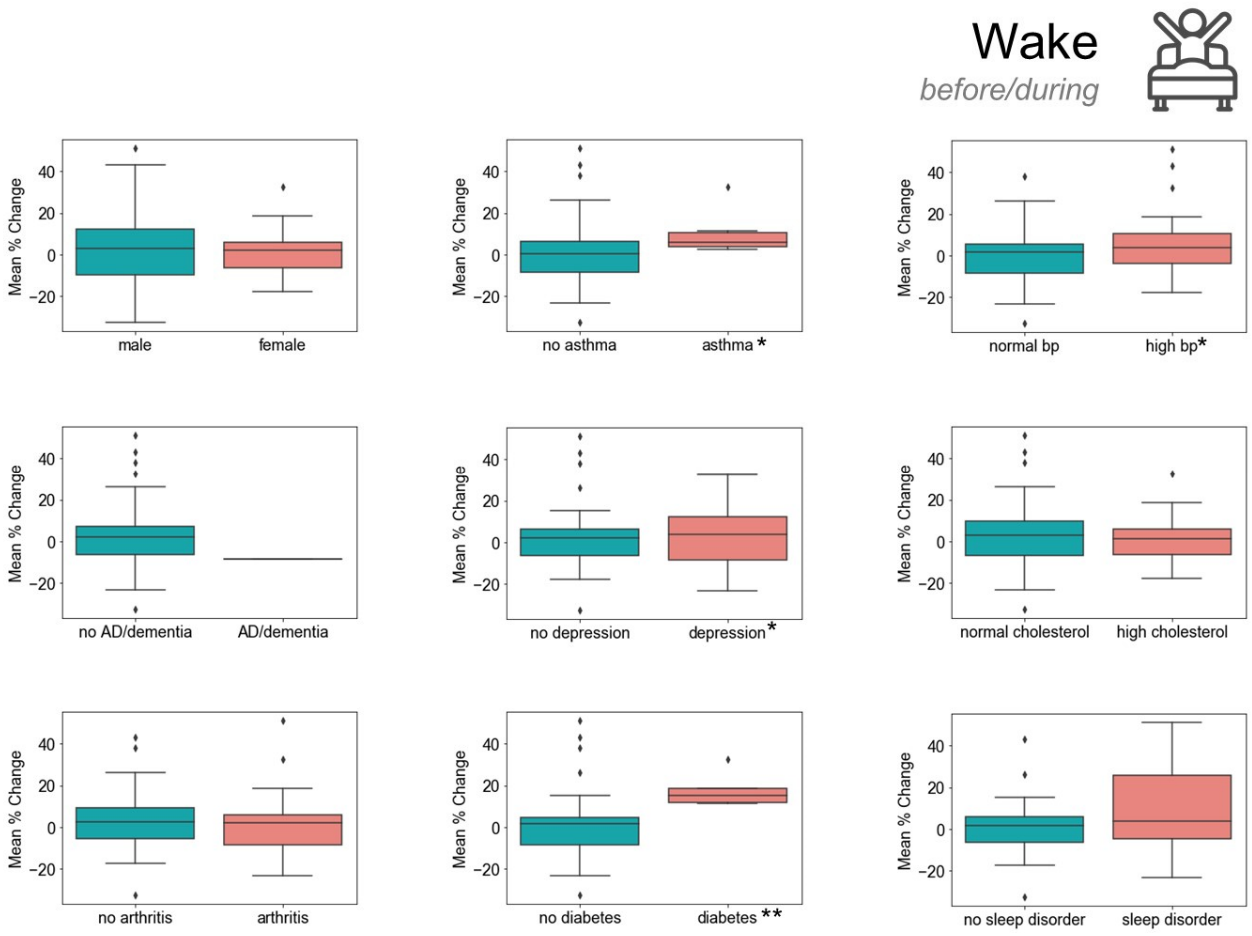


**Figure E5.** Wake trend before/during SAHO by demographic and health data.

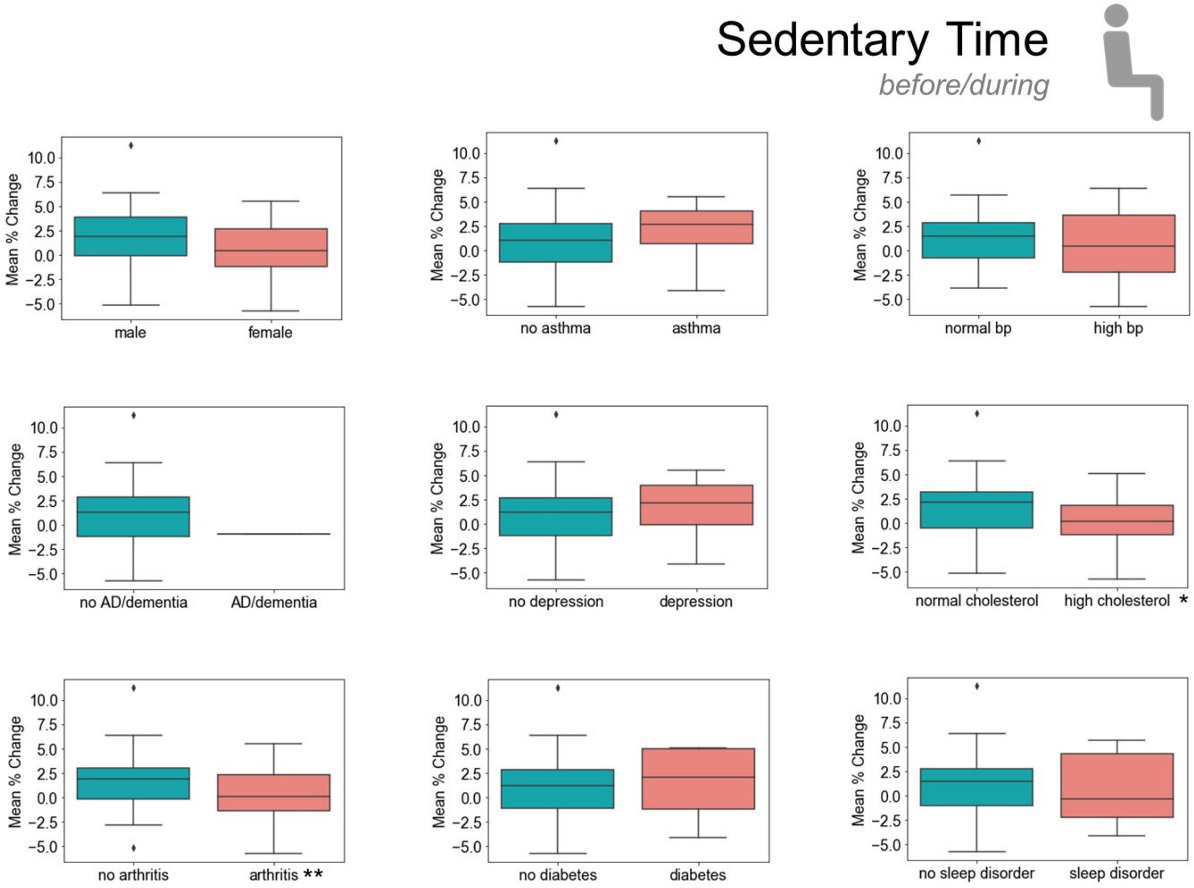


**Figure E6.** Sedentary active trend before/during SAHO by demographic and health data.

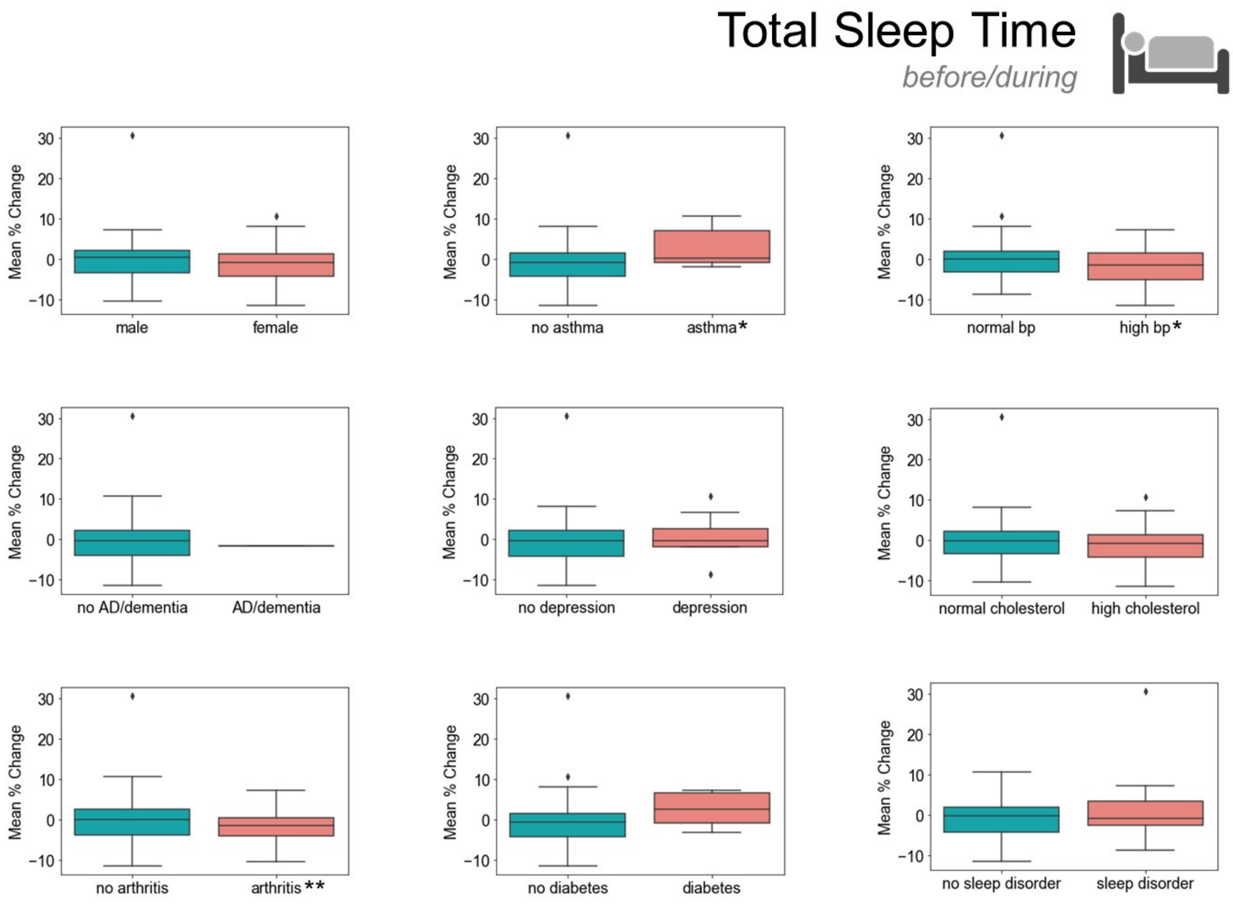


**Figure E7.** Total sleep time trend before/during SAHO by demographic and health data.

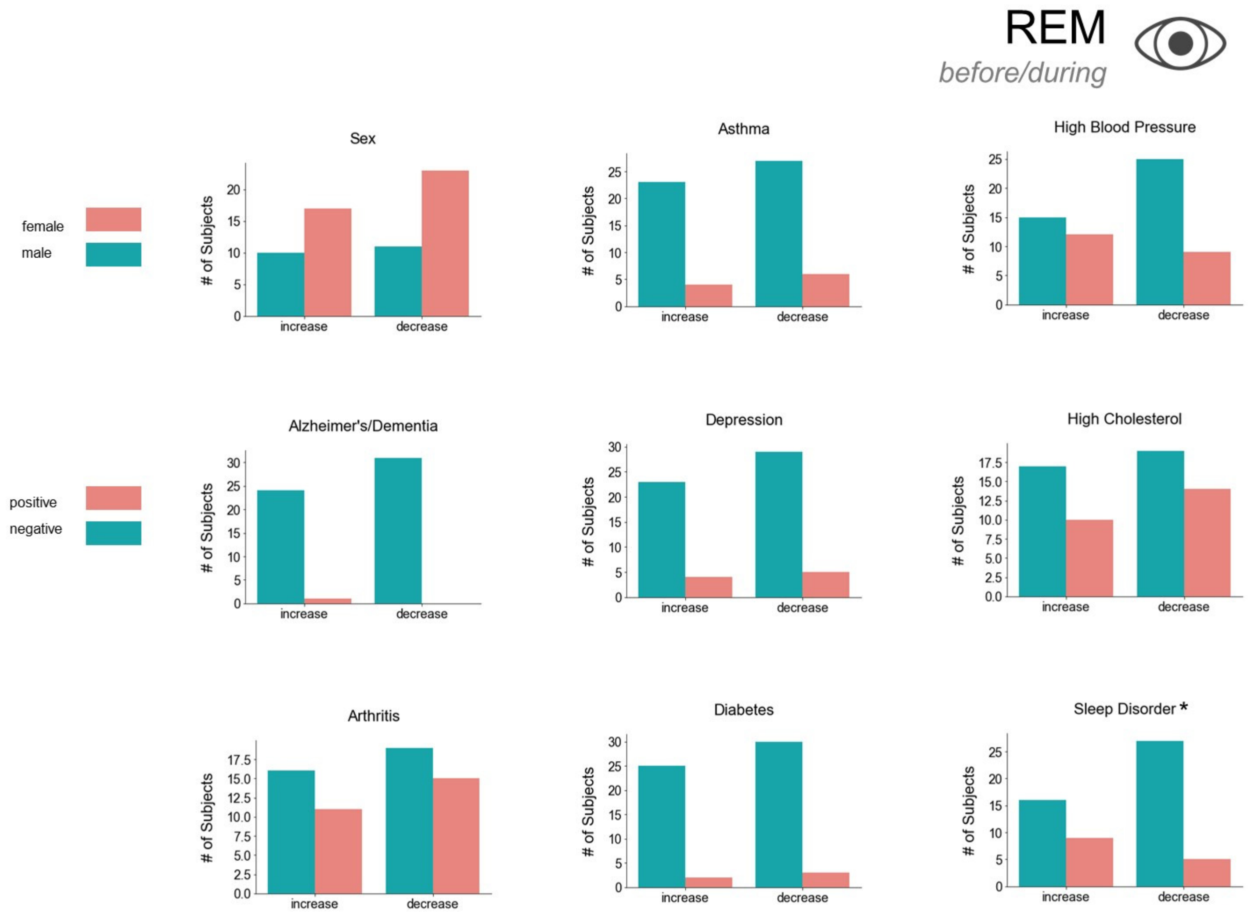


**Figure E8.** REM trend by demographics and health data before/during SAHO divided by those that increased or decreased activity.

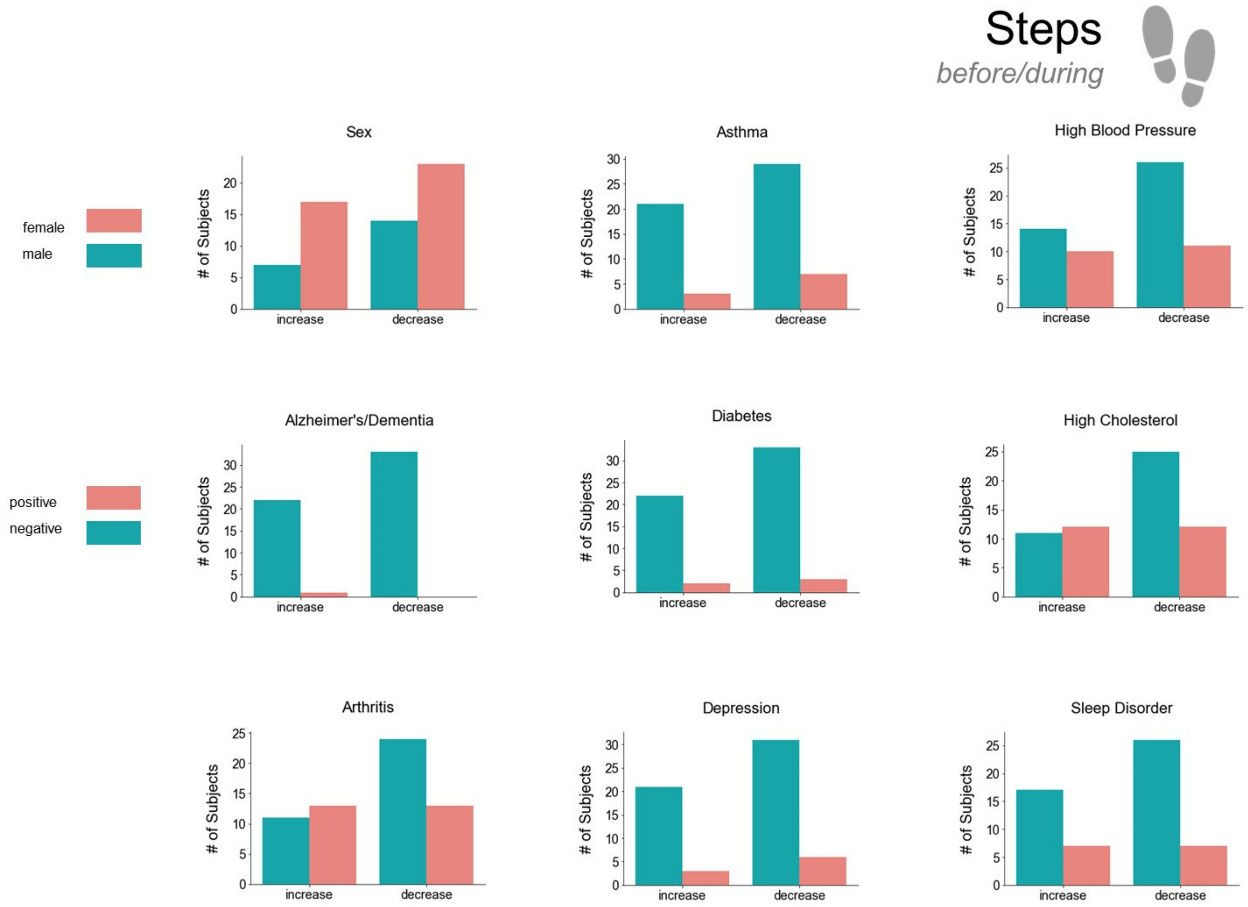


**Figure E9.** Step trend by demographics and health data before/during SAHO divided by those that increased or decreased activity.

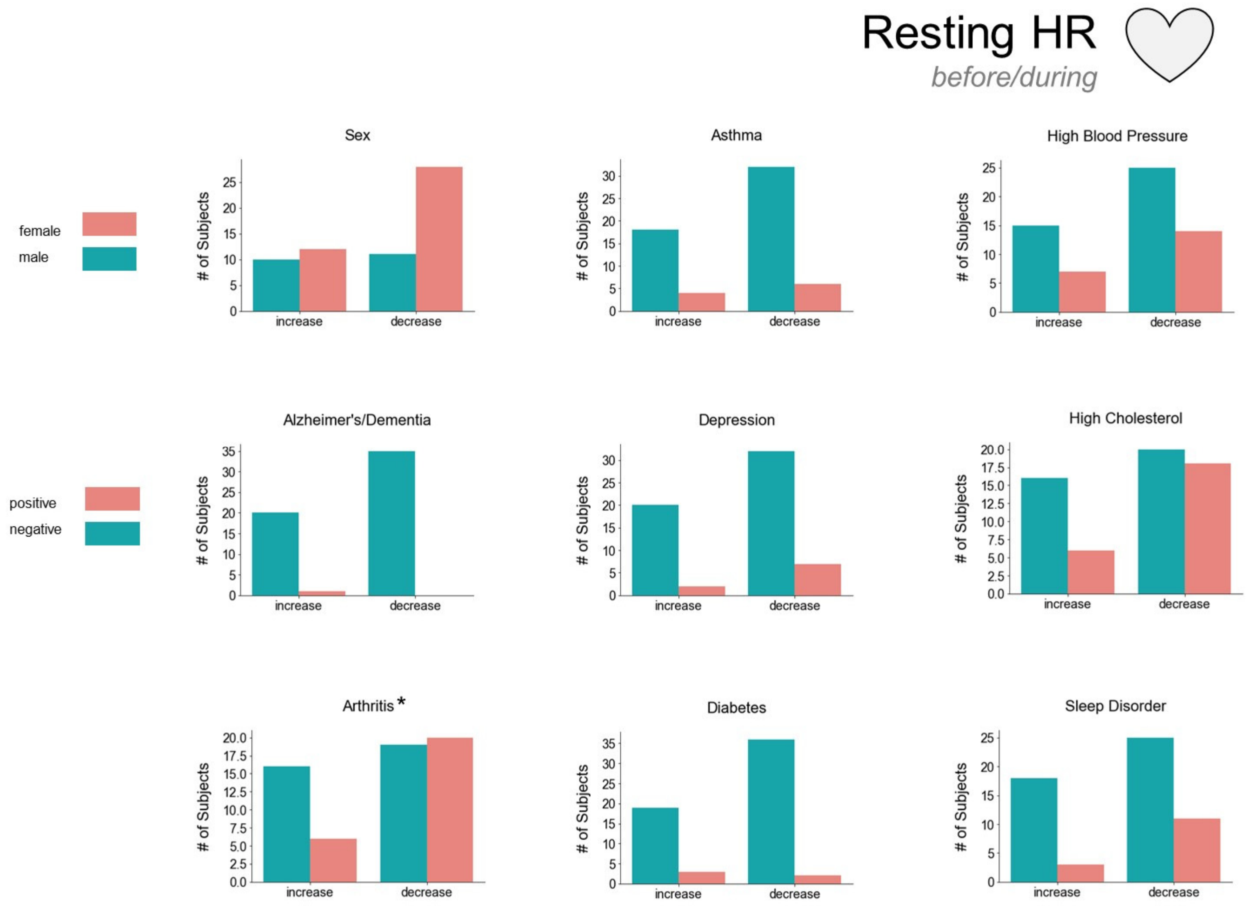


**Figure E10.** Resting heart rate trend by demographics and health data before/during SAHO divided by those that increased or decreased activity.

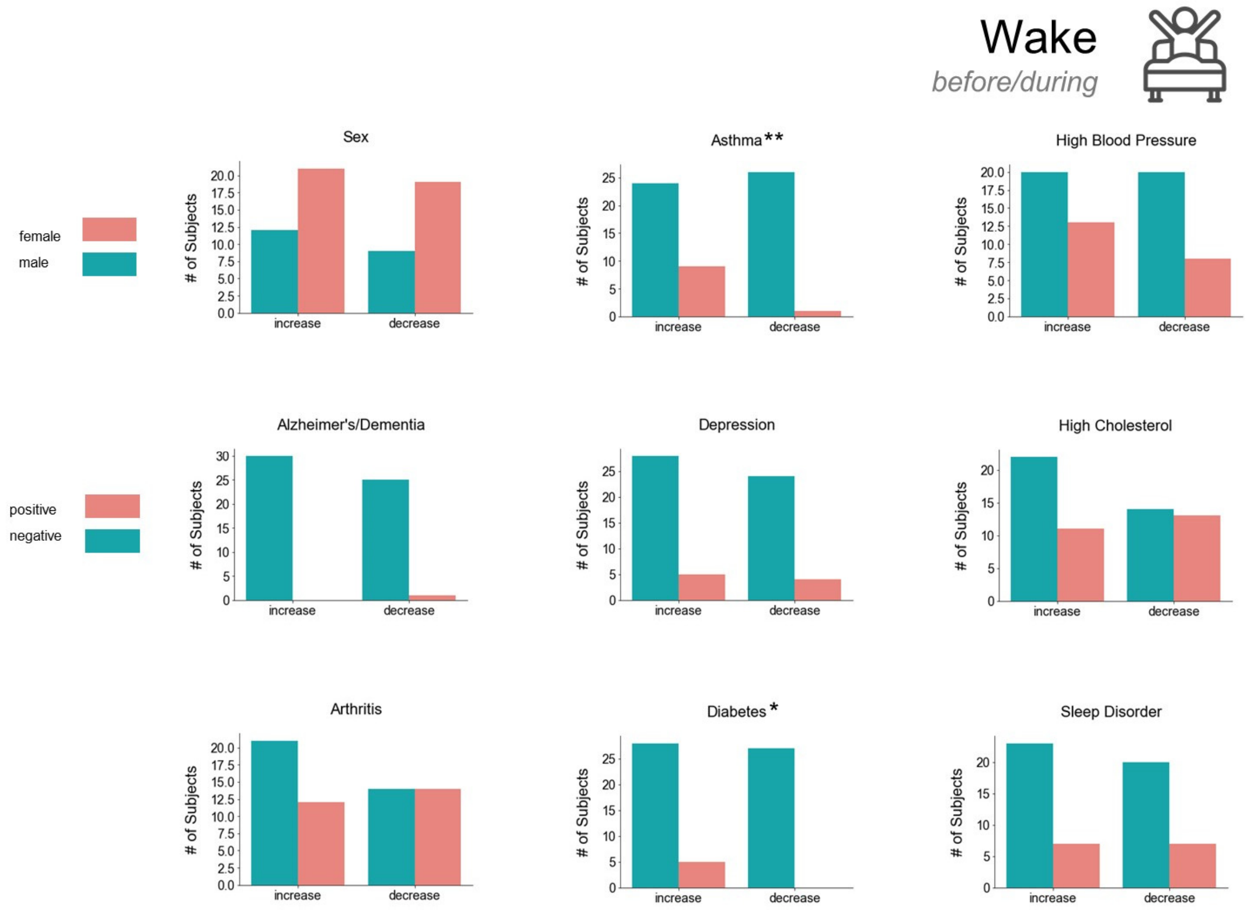


**Figure E11.** Wake trend by demographics and health data before/during SAHO divided by those that increased or decreased activity.

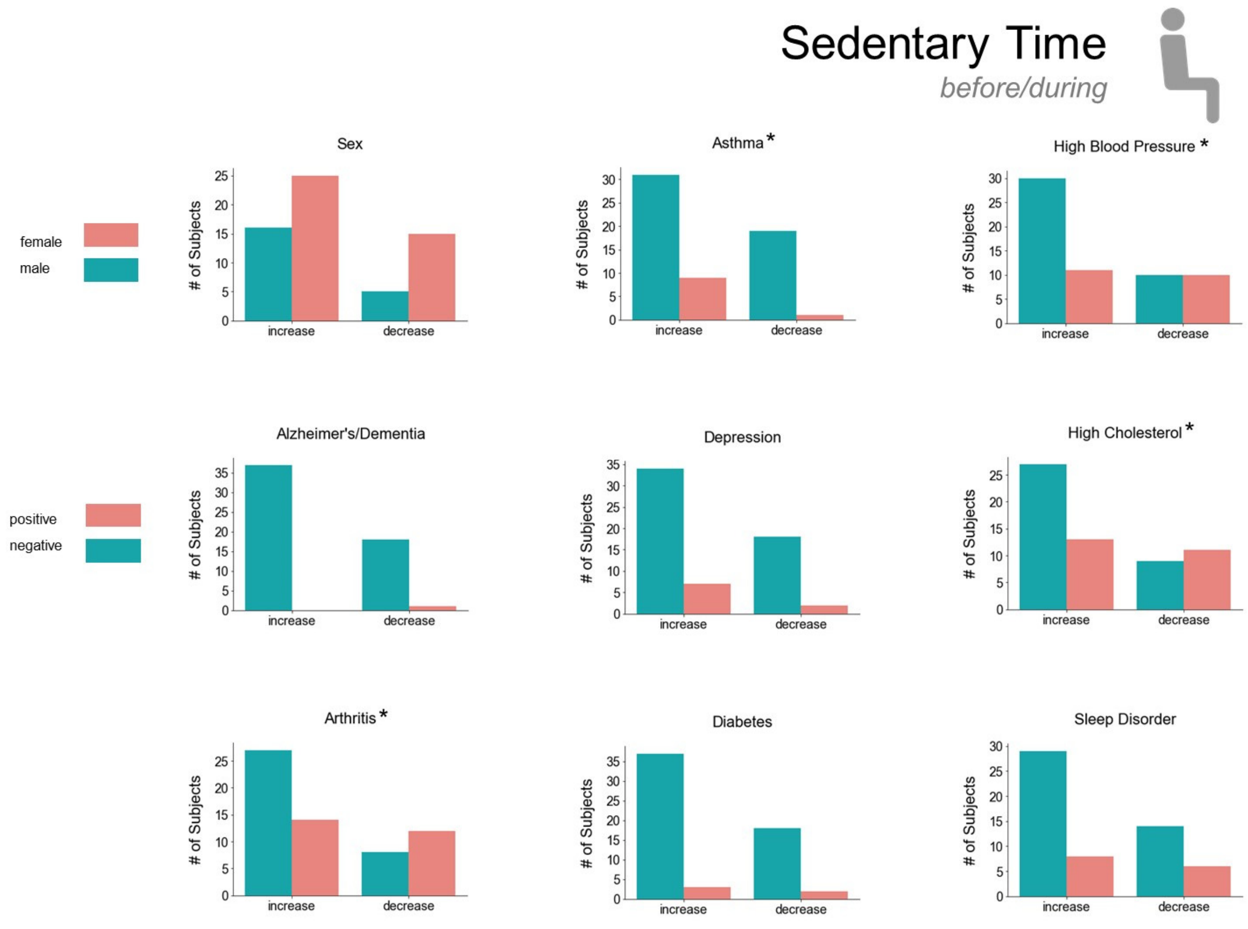


**Figure E12.** Sedentary active trend by demographics and health data before/during SAHO divided by those that increased or decreased activity.

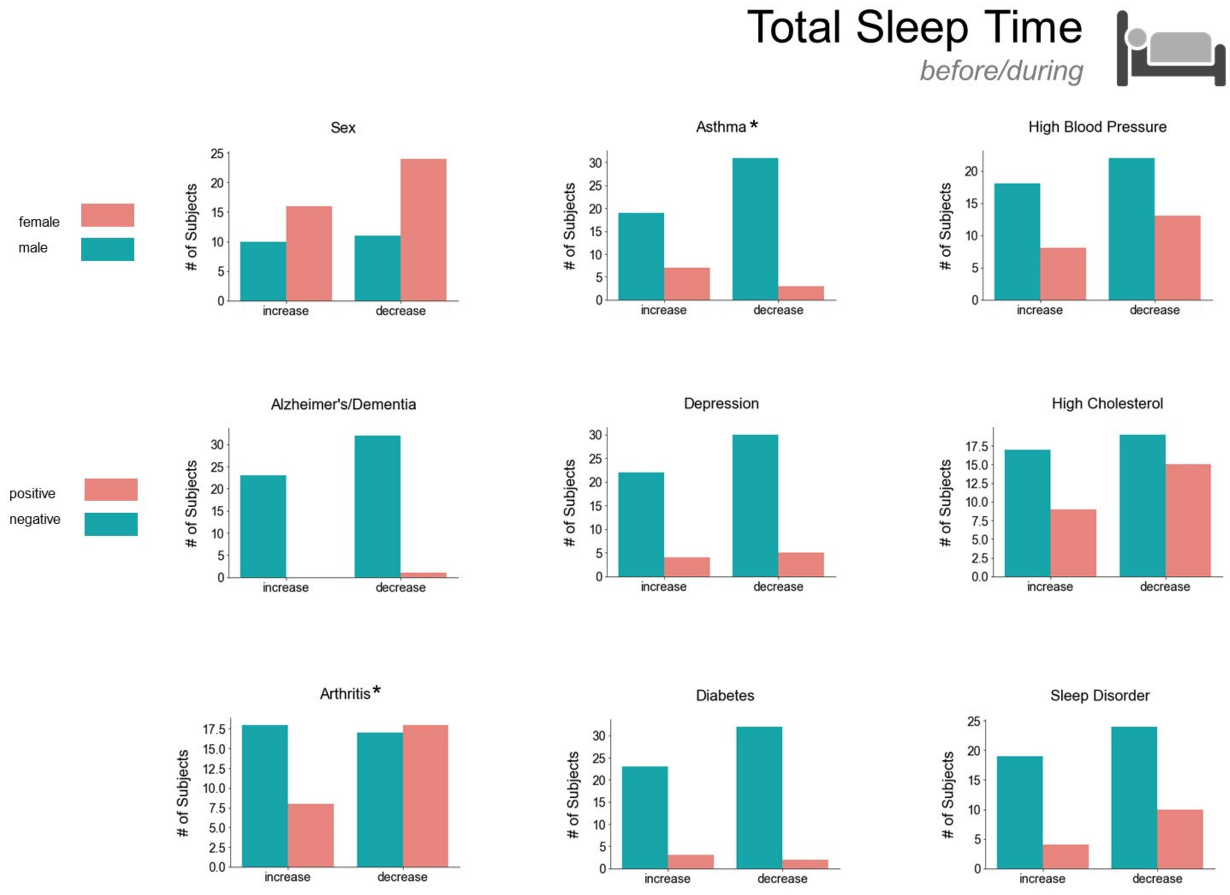


**Figure E13.** Total sleep time trend by demographics and health data before/during SAHO divided by those that increased or decreased activity.

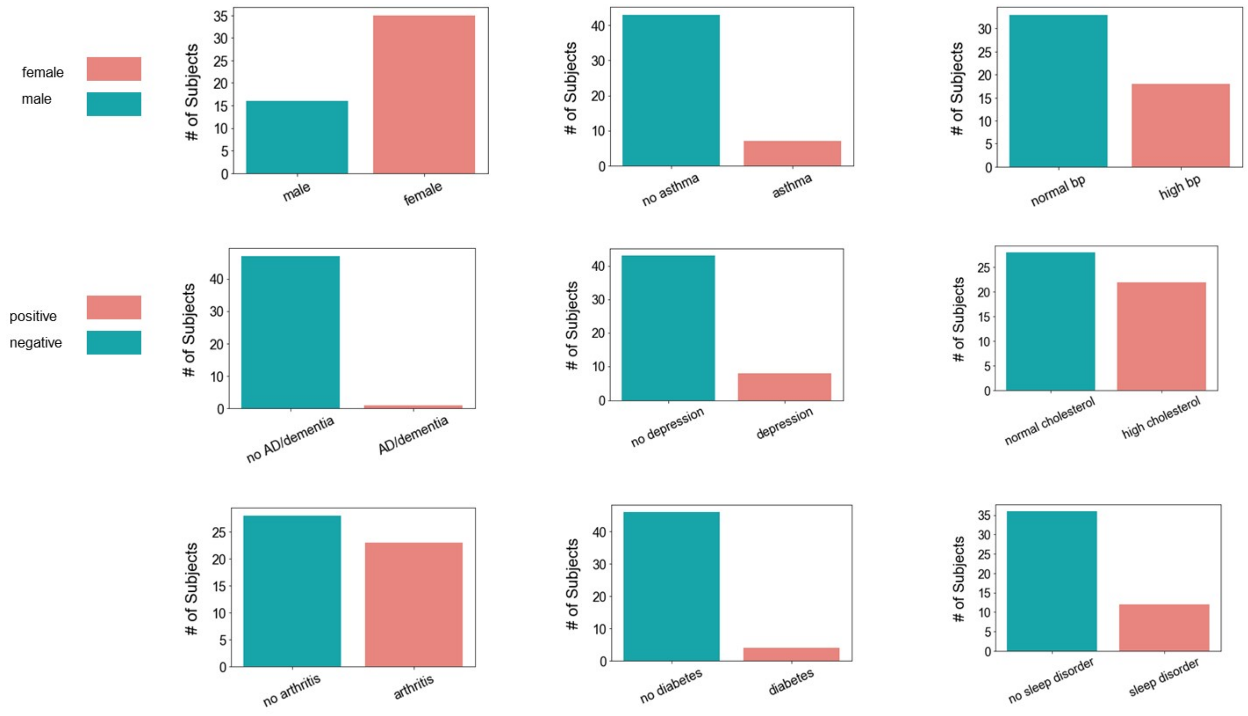


**Figure E14.** Overall demographics and health data on all subjects included in during/after dataset. Subjects were given a questionnaire upon entering the study.

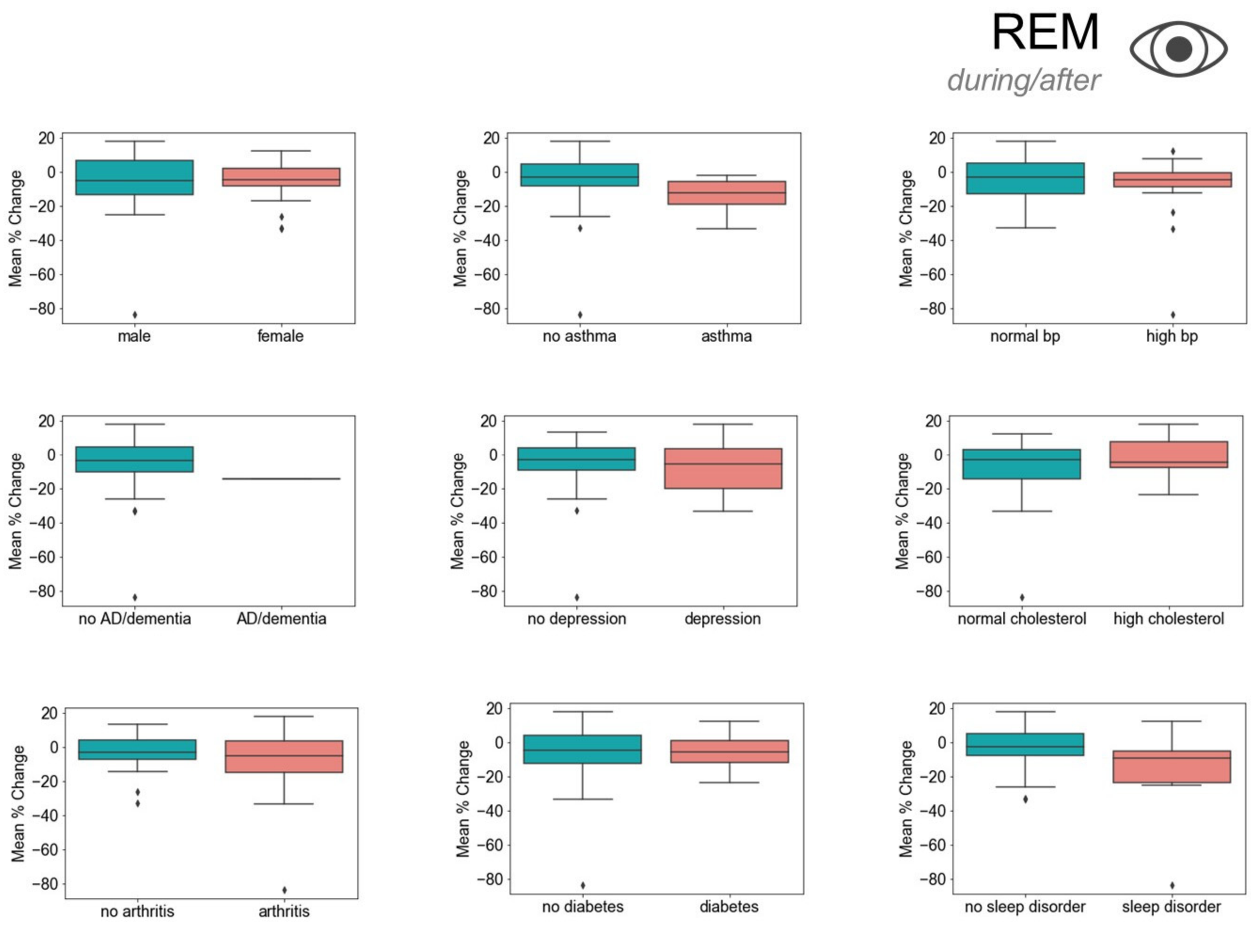


**Figure E15.** REM trend during/after SAHO by demographic and health data.

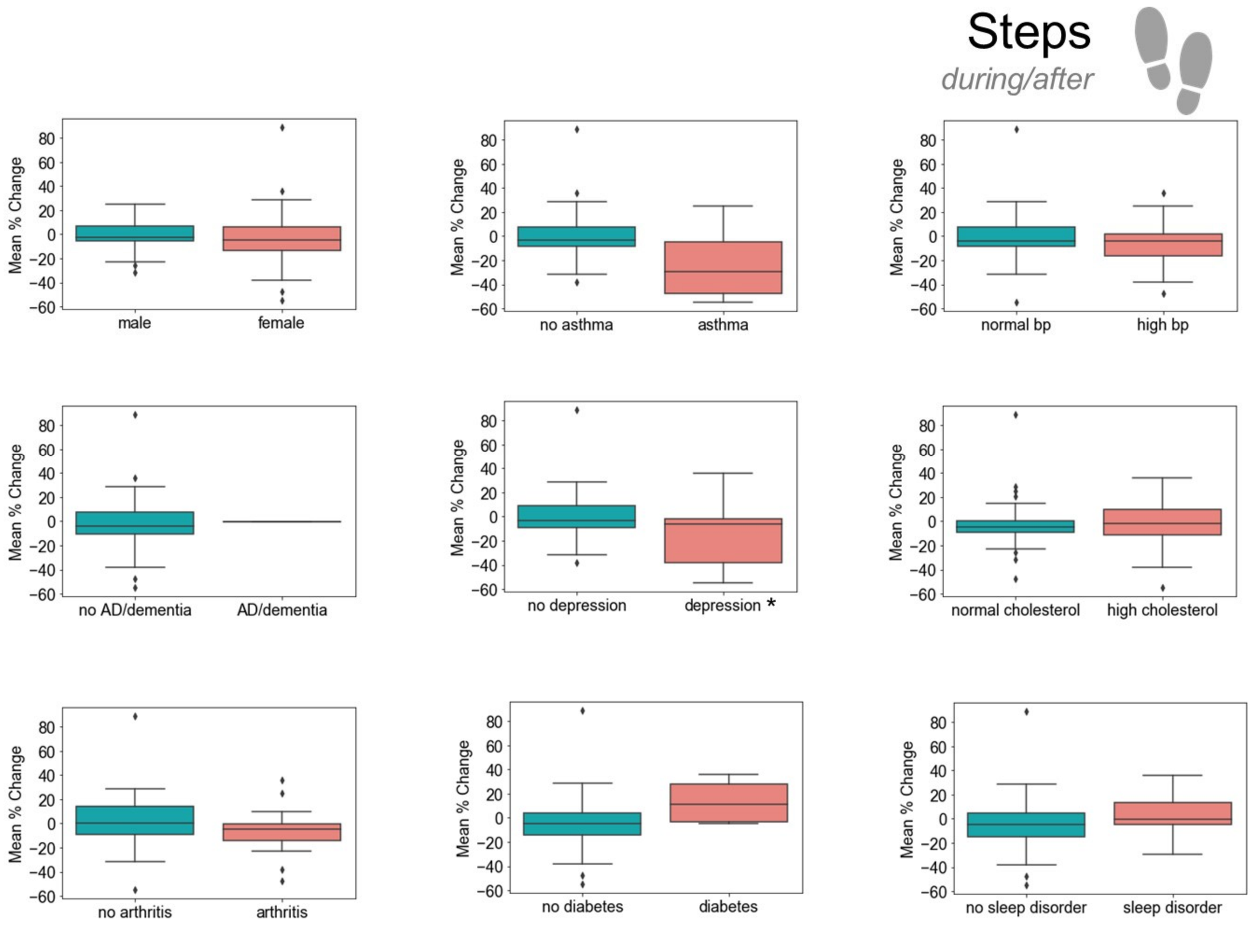


**Figure E16.** Step trend during/after SAHO by demographic and health data.

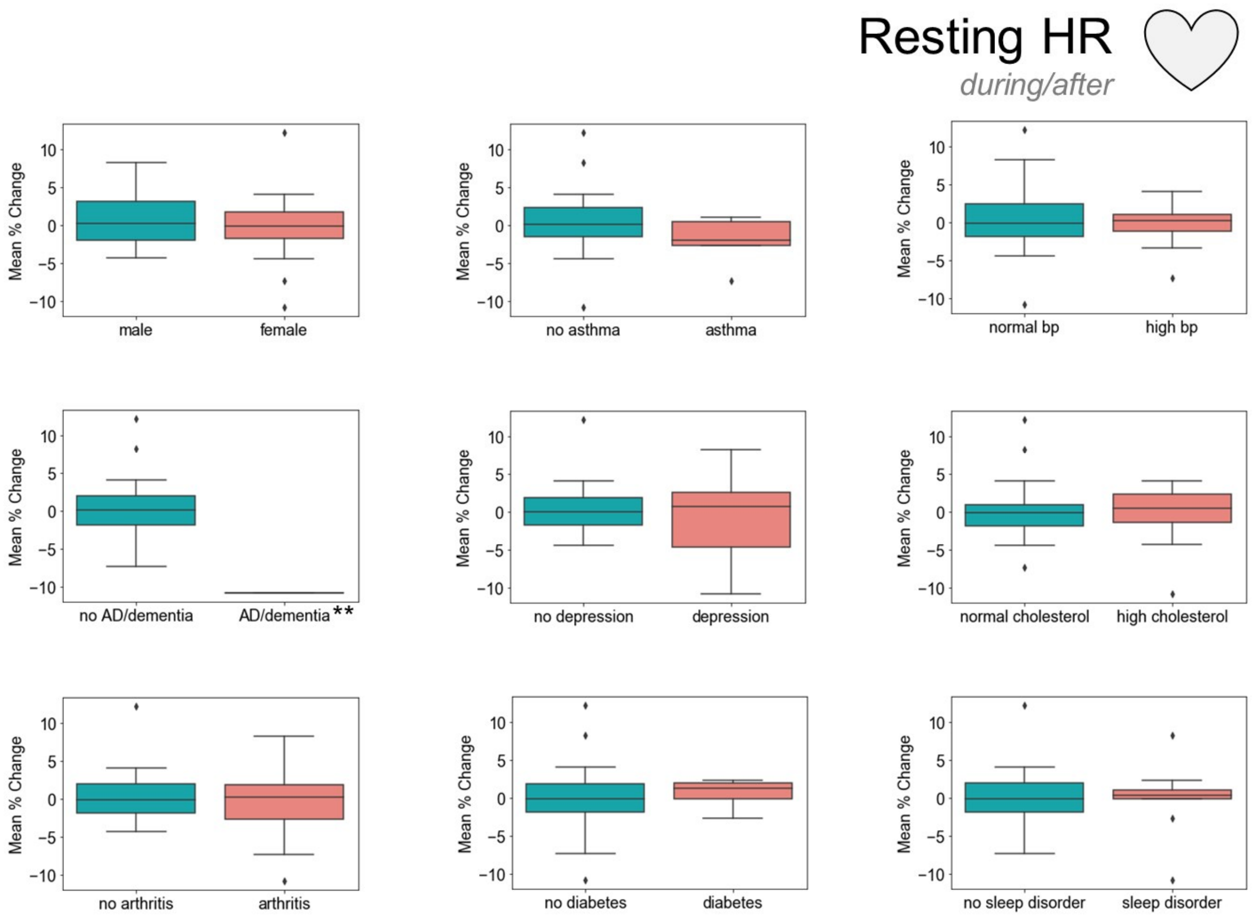


**Figure E17.** Resting heart rate trend during/after SAHO by demographic and health data.

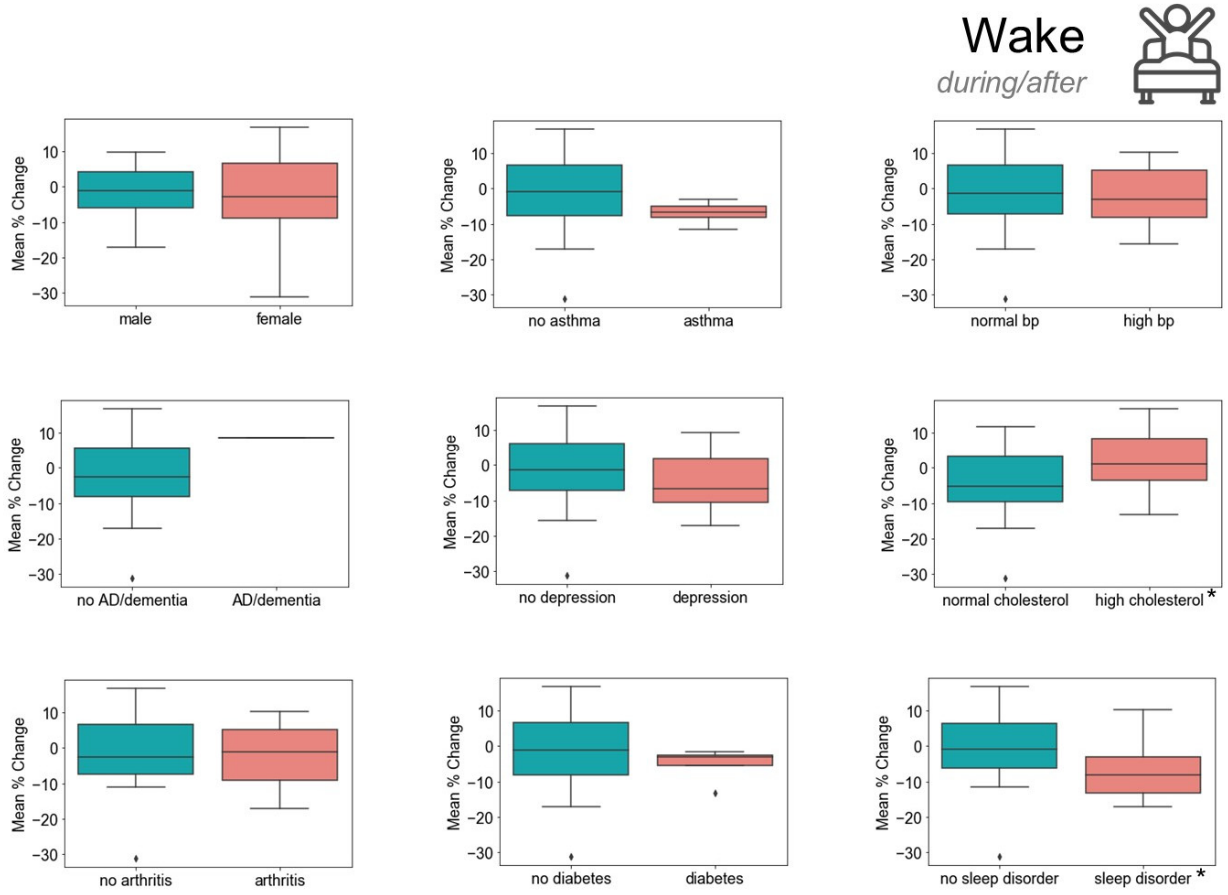


**Figure E18.** Wake trend during/after SAHO by demographic and health data.

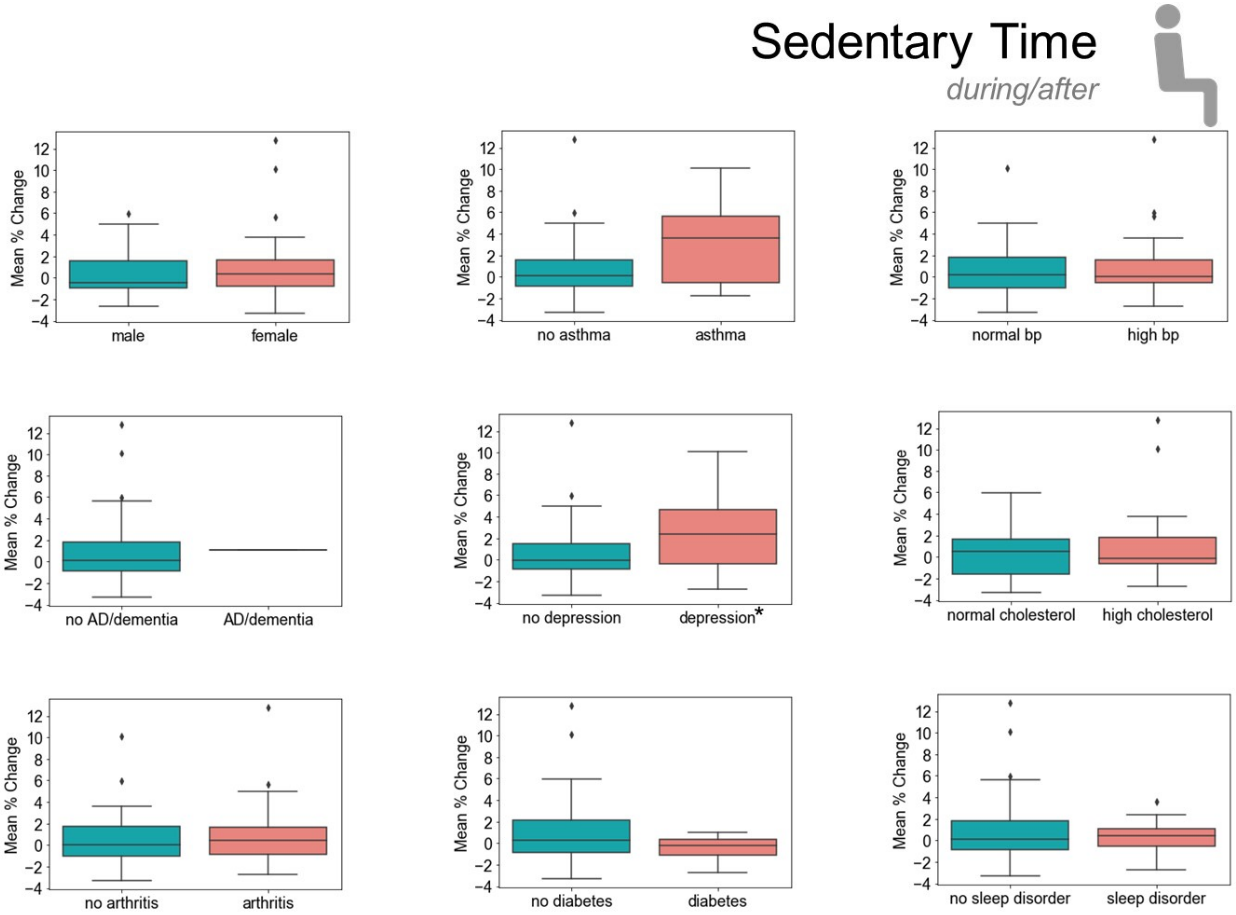


**Figure E19.** Sedentary active trend during/after SAHO by demographic and health data.

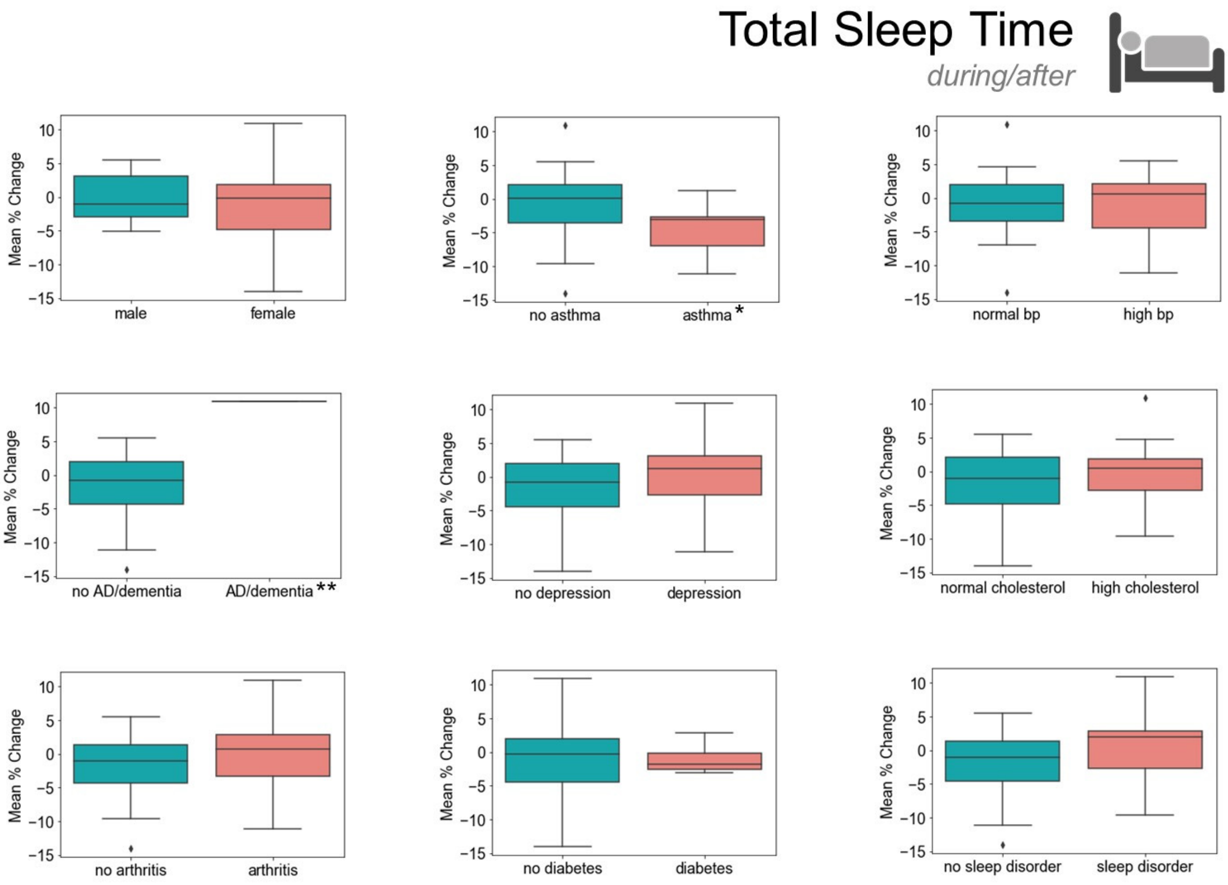


**Figure E20.** Total sleep time trend during/after SAHO by demographic and health data.

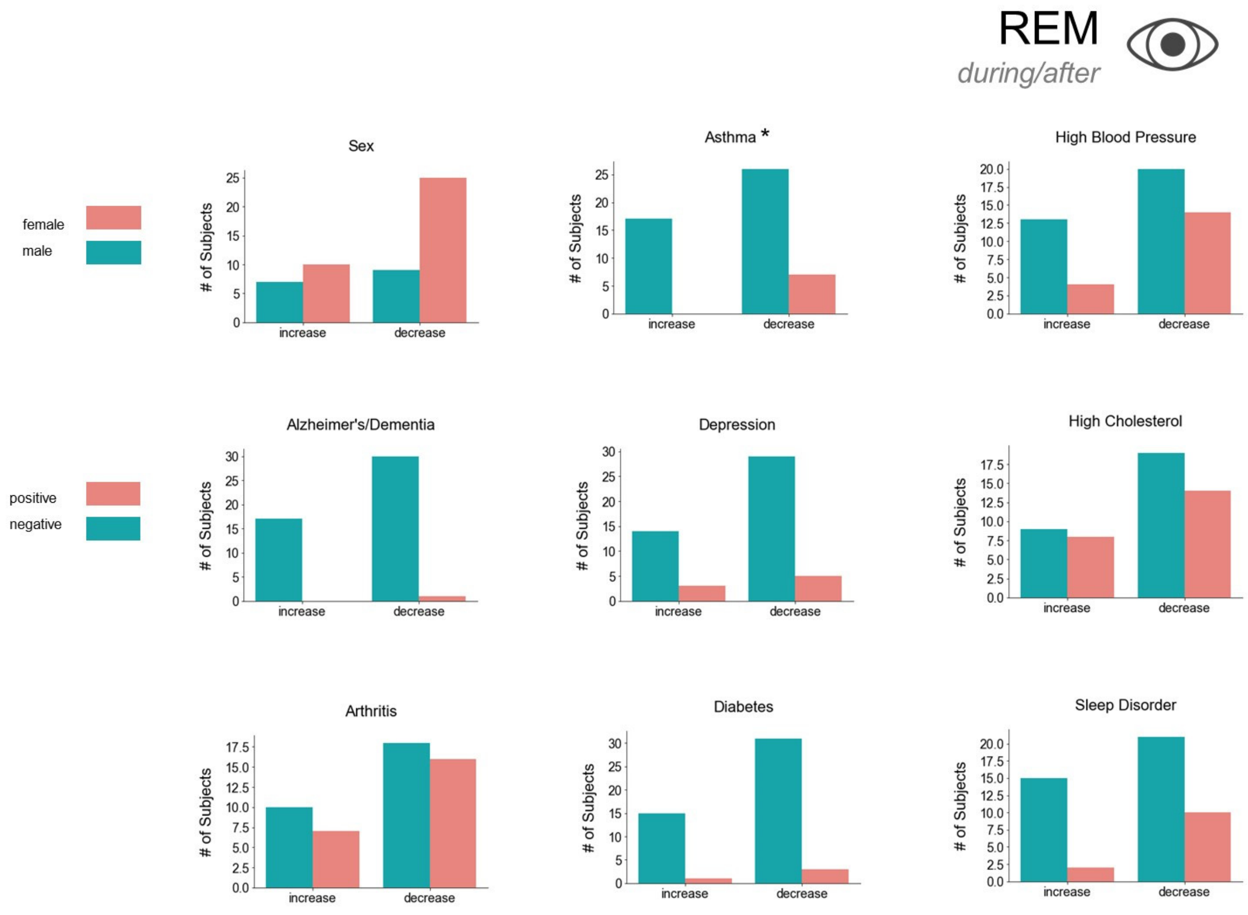


**Figure E21.** REM trend by demographics and health data during/after SAHO divided by those that increased or decreased activity.

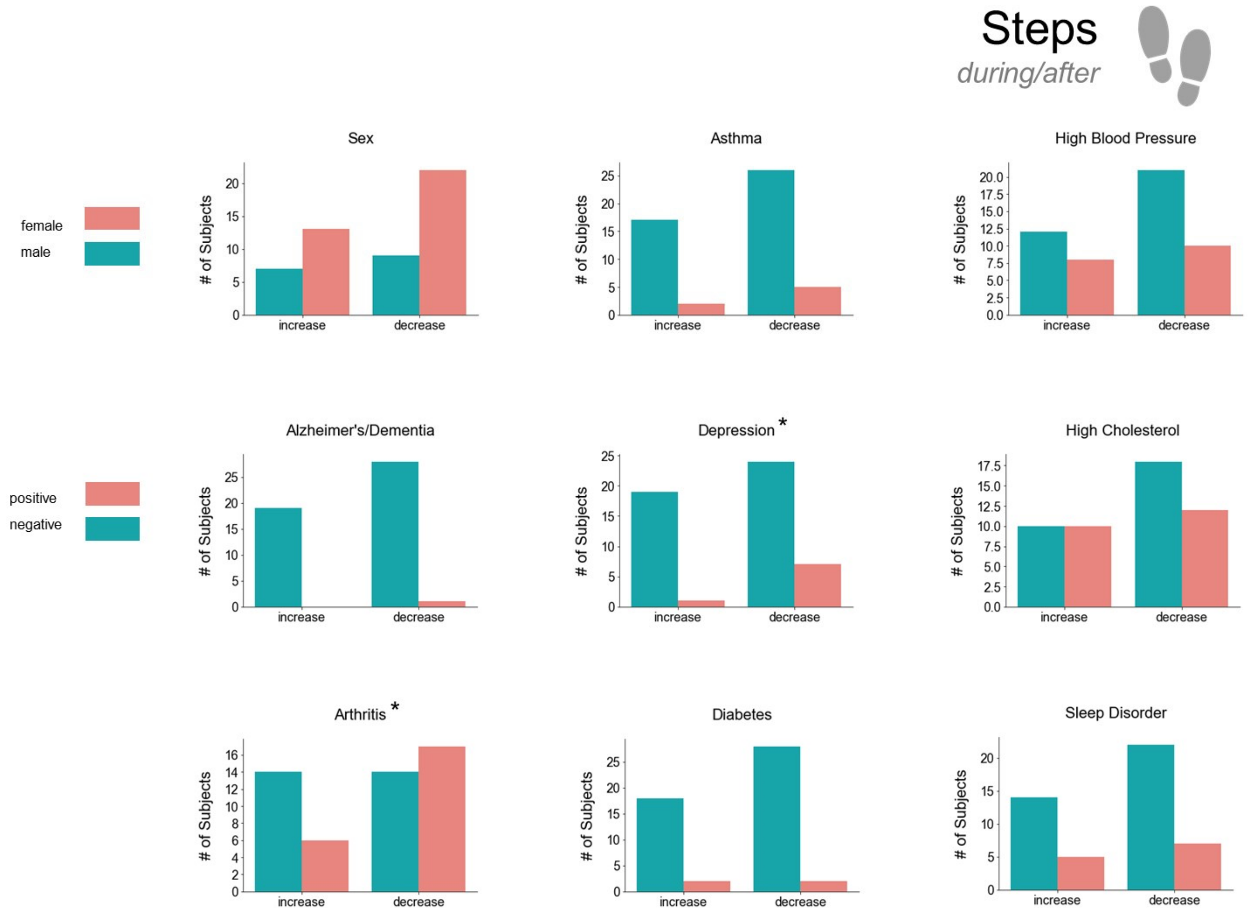


**Figure E22.** Step trend by demographics and health data during/after SAHO divided by those that increased or decreased activity.

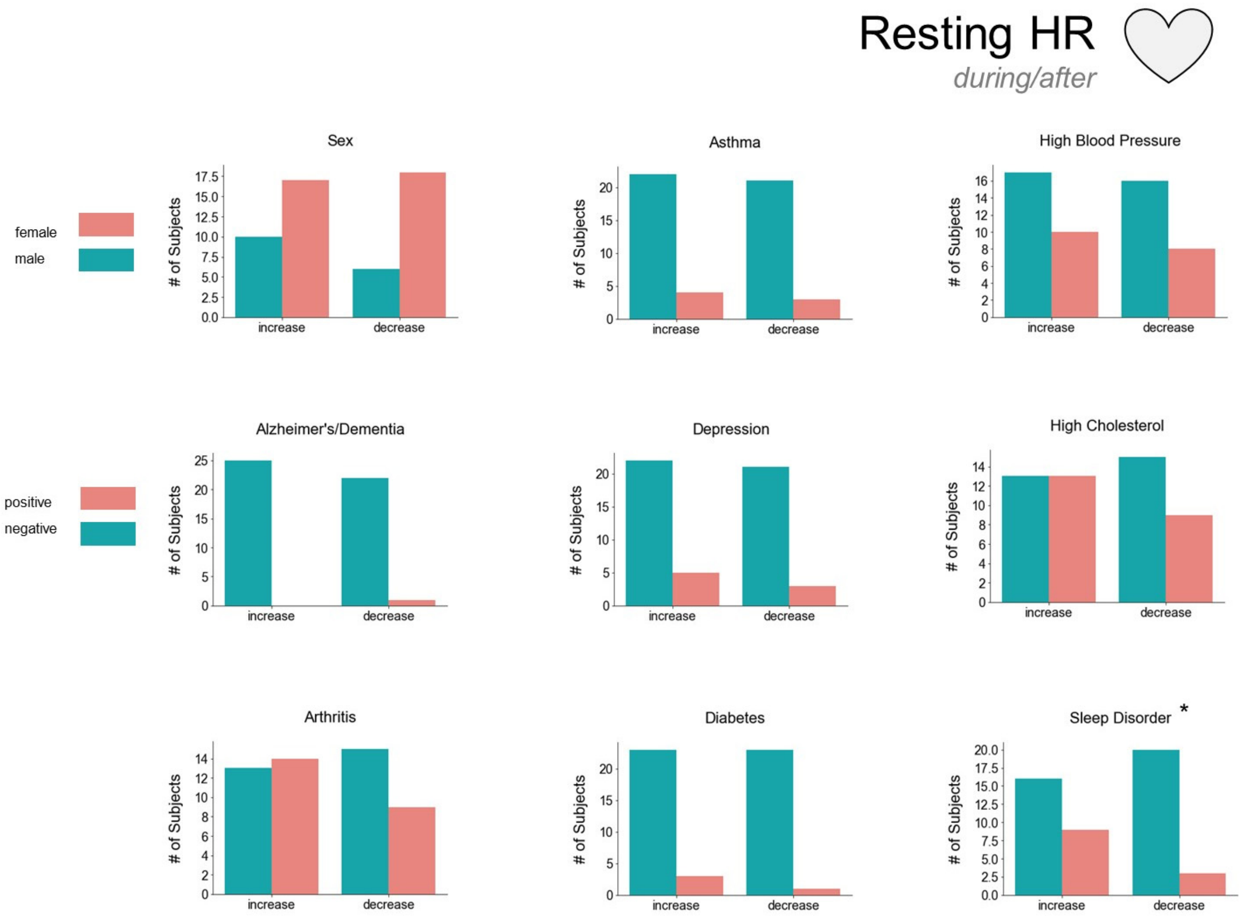


**Figure E23.** Resting heart rate trend by demographics and health data during/after SAHO divided by those that increased or decreased activity.

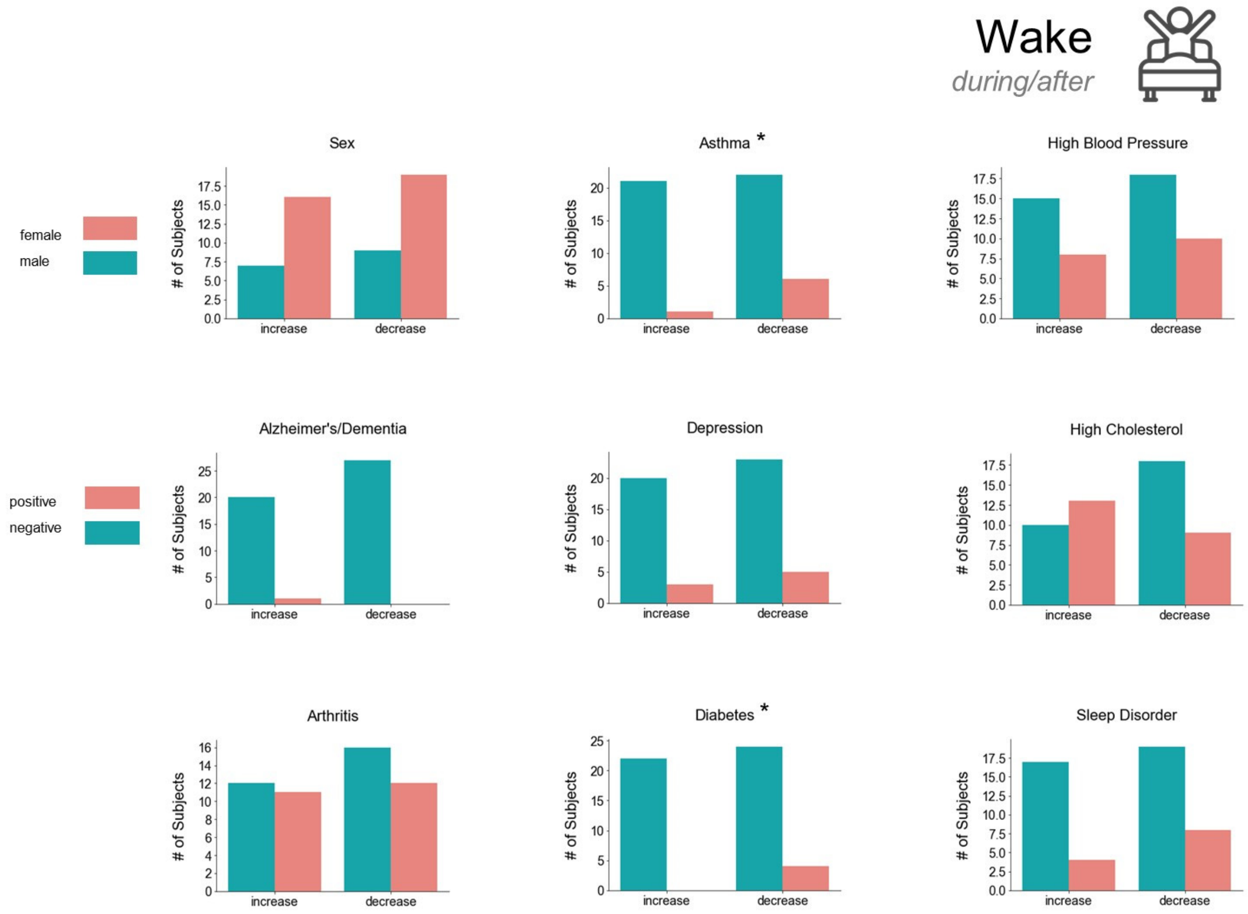


**Figure E24.** Wake trend by demographics and health data during/after SAHO divided by those that increased or decreased activity.

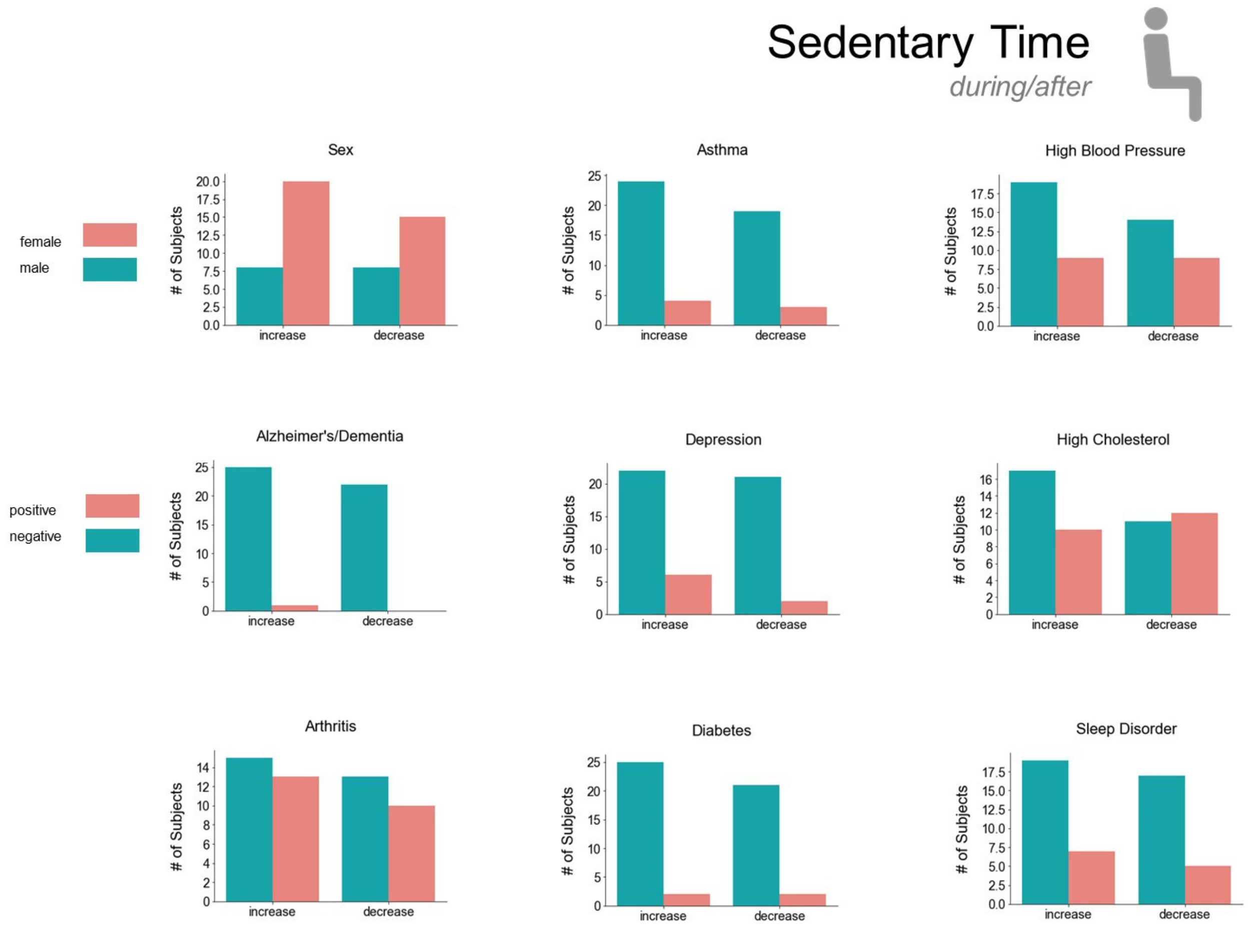


**Figure E25.** Sedentary active trend by demographics and health data during/after SAHO divided by those that increased or decreased activity.

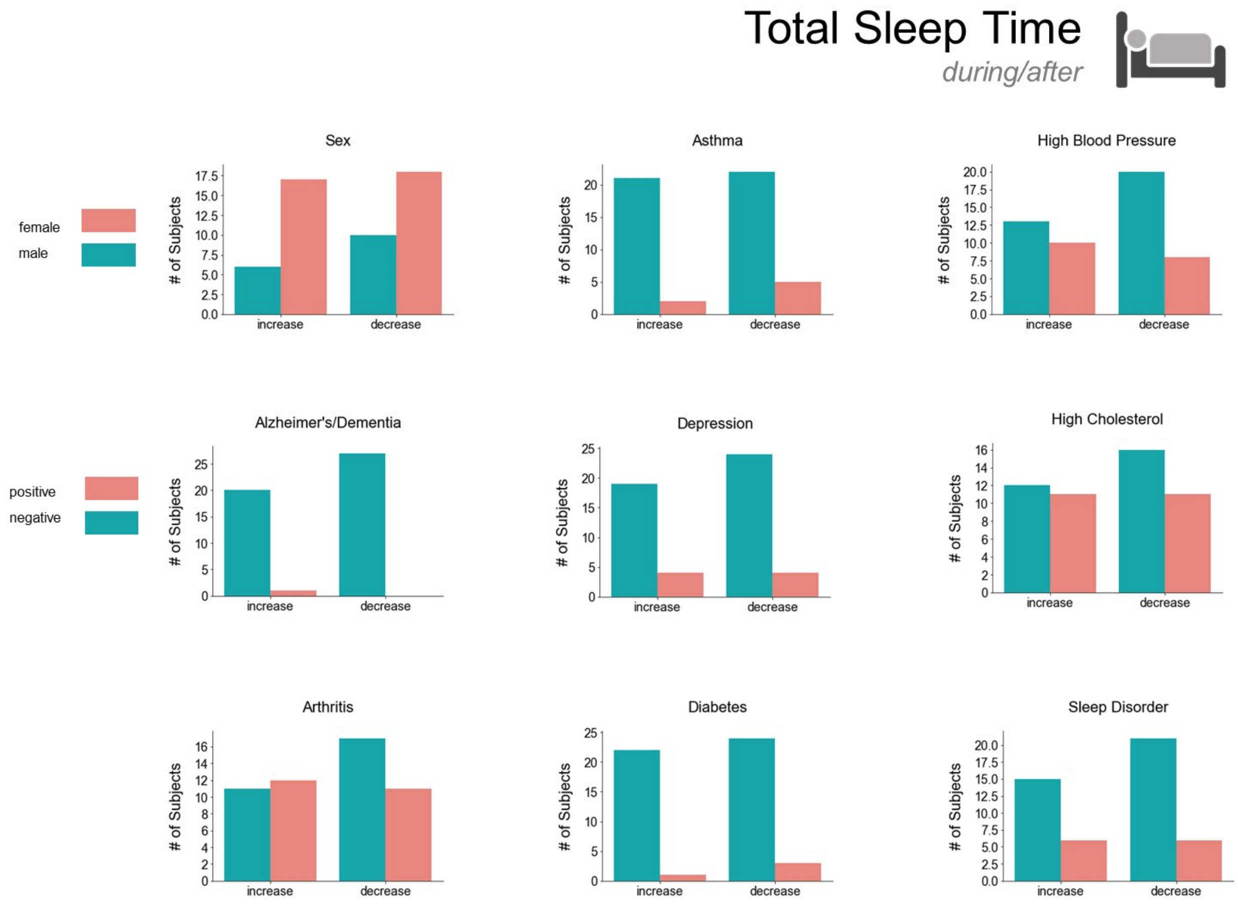


**Figure E26.** Total sleep time trend by demographics and health data during/after SAHO divided by those that increased or decreased activity.

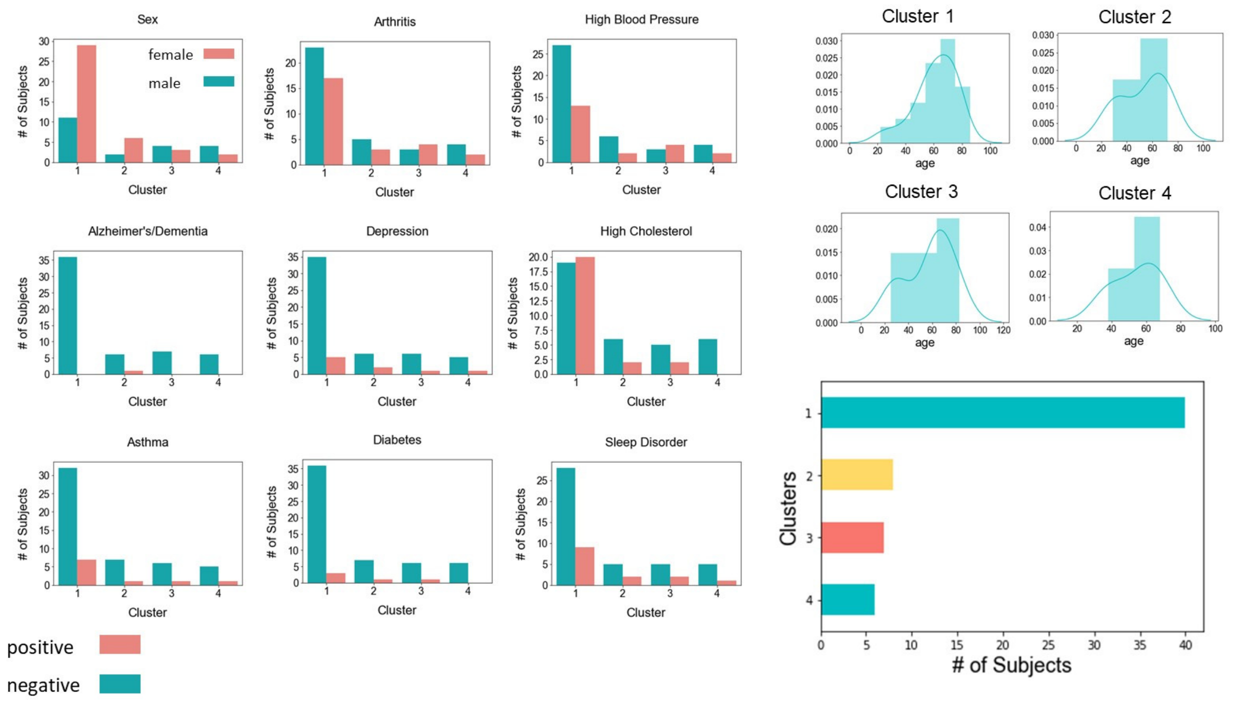


**Figure E27.** Demographic and health graphs for clusters found by Shrinkage Clustering method from the dataset containing the % change in means and slope of the linear trendlines comparing before/during the SAHO.

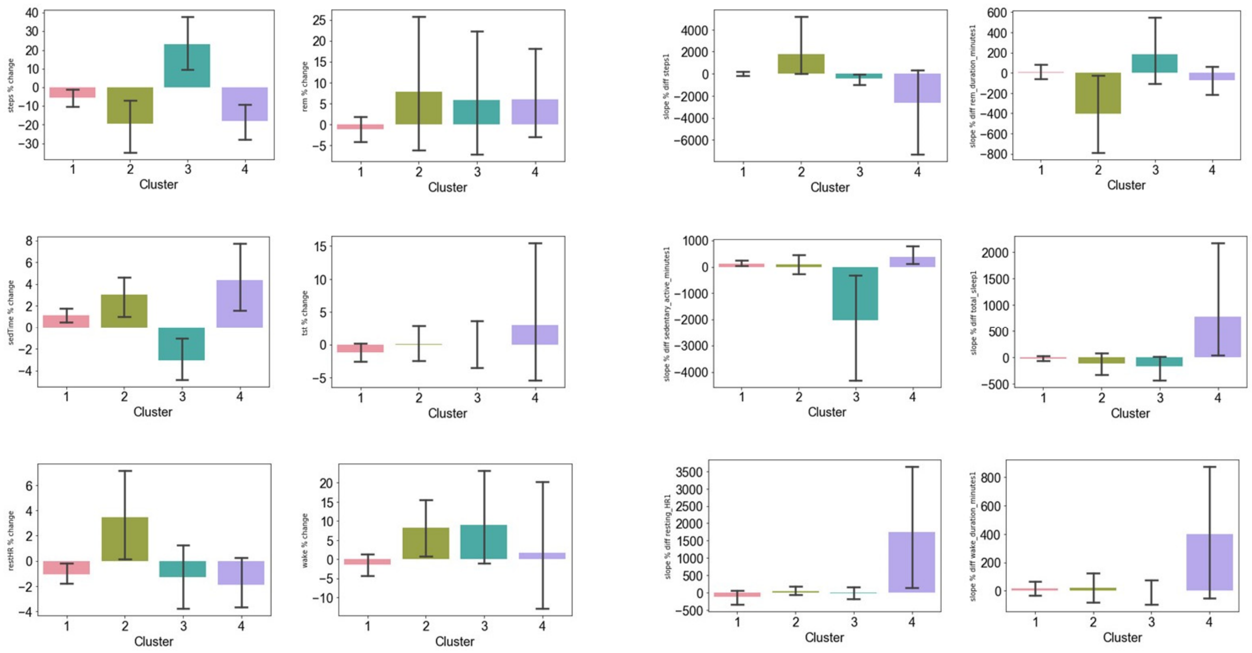

**Figure E28.** Mean % change (left) and % change of slope of the linear trendline (right) for before/during the SAHO by activity.

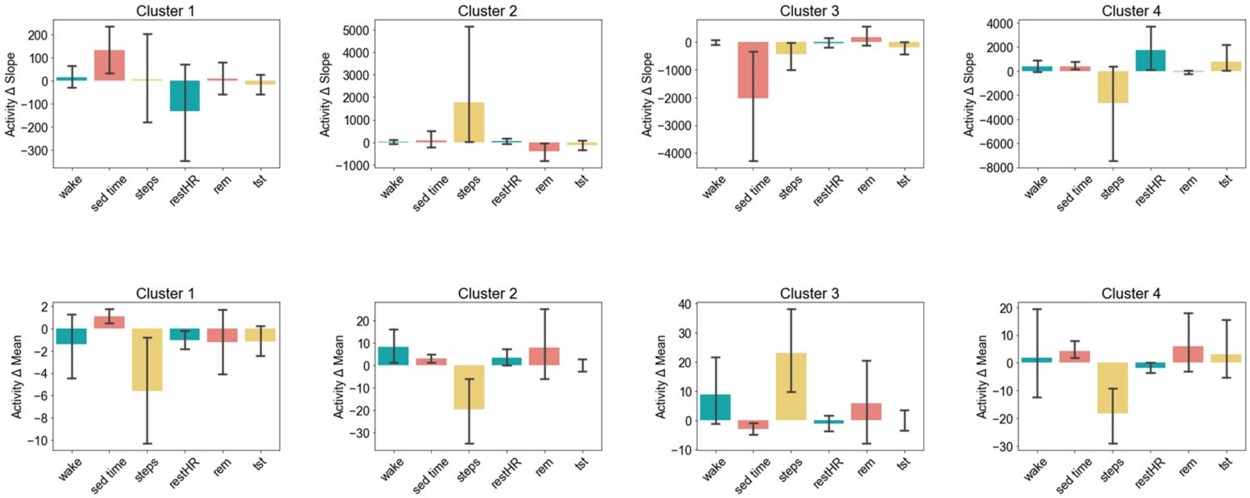


**Figure E29.** Mean % change (left) and % change of slope of the linear trendline (right) for before/during the SAHO by cluster.

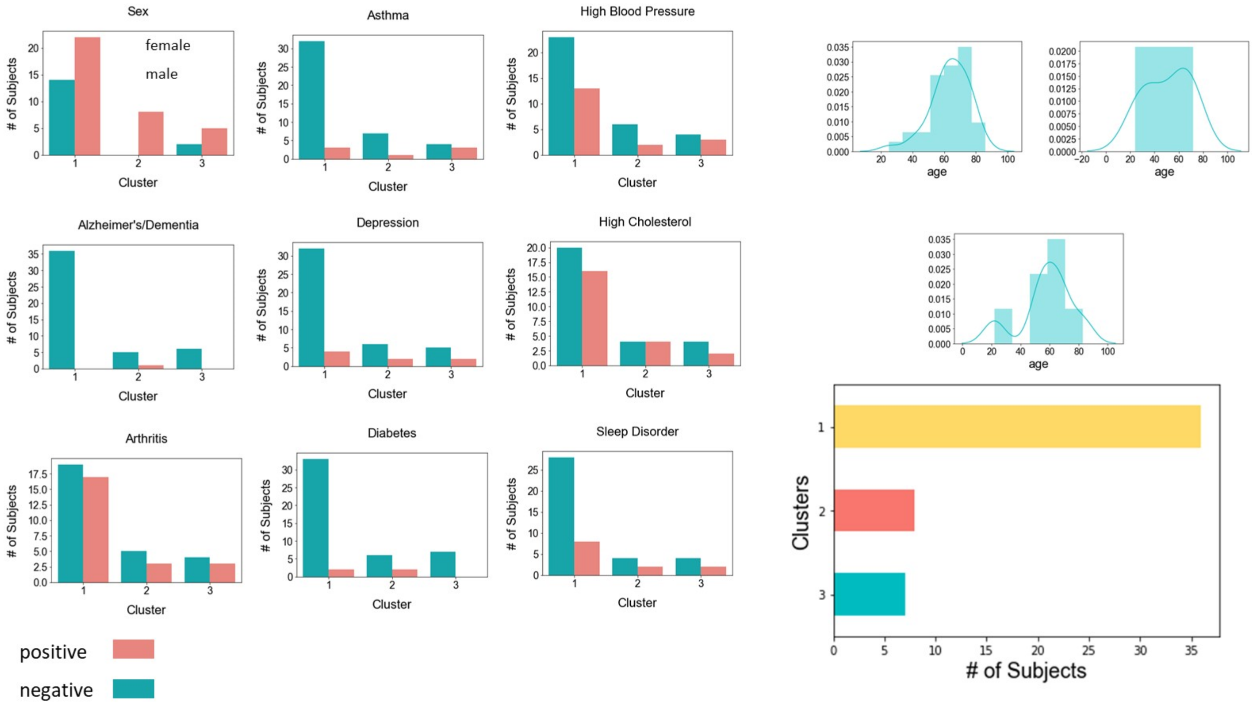


**Figure E30.** Demographic and health graphs for clusters found by Shrinkage Clustering method from the dataset containing the % change in means and slope of the linear trendlines comparing during/after the SAHO.

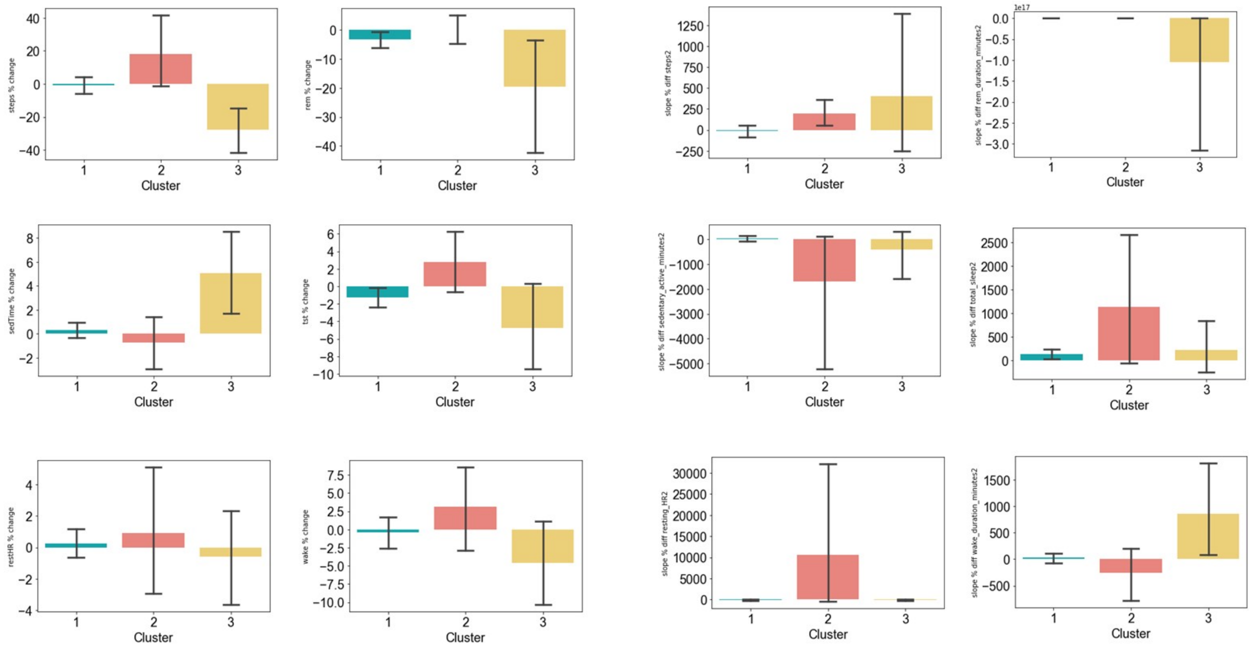


**Figure E31.** Mean % change (left) and % change of slope of the linear trendline (right) for during/after the SAHO by activity.

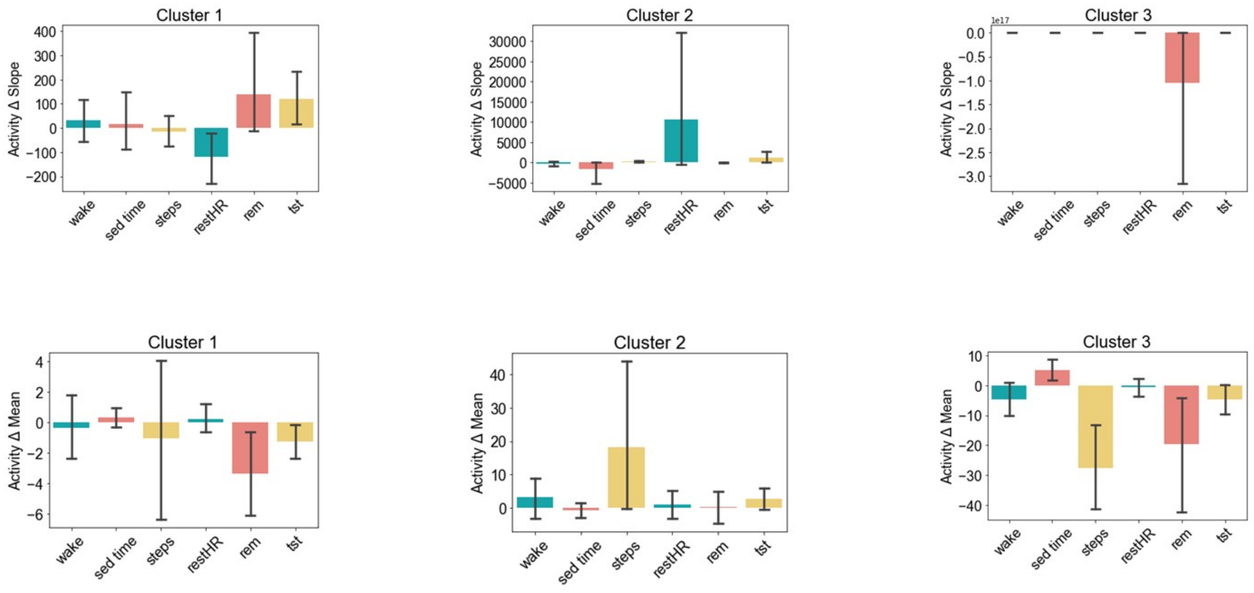


**Figure E32.** Mean % change (left) and % change of slope of the linear trendline (right) for during/after the SAHO by cluster.

#### Appendix F. Activity Trends by Cluster

- **Before/During Stay-at-Home Orders:**
  - **Figure F1.** 7-day moving average of REM duration in minutes with linear trendline. The bolded trace is the average for the cluster; the light grey tracings are for each individual in the cluster.
  - **Figure F2**. 7-day moving average of steps with linear trendline. The bolded trace is the average for the cluster; the light grey tracings are for each individual in the cluster.
  - **Figure F3.** 7-day moving average of resting heart rate in beats per minute with linear trendline. The bolded trace is the average for the cluster; the light grey tracings are for each individual in the cluster.
  - **Figure F4.** 7-day moving average of wake duration during sleep period in minutes with linear trendline. The bolded trace is the average for the cluster; the light grey tracings are for each individual in the cluster.
  - **Figure F5.** 7-day moving average of sedentary time in minutes with linear trendline. The bolded trace is the average for the cluster; the light grey tracings are for each individual in the cluster.
  - **Figure F6.** 7-day moving average of total sleep time in minutes with linear trendline. The bolded trace is the average for the cluster; the light grey tracings are for each individual in the cluster.

- **During/After Stay-at-Home Orders:**
  - **Figure F7.** 7-day moving average of REM duration in minutes with linear trendline. The bolded trace is the average for the cluster; the light grey tracings are for each individual in the cluster.
  - **Figure F8.** 7-day moving average of steps with linear trendline. The bolded trace is the average for the cluster; the light grey tracings are for each individual in the cluster.
  - **Figure F9.** 7-day moving average of resting heart rate in beats per minute with linear trendline. The bolded trace is the average for the cluster; the light grey tracings are for each individual in the cluster.
  - **Figure F10.** 7-day moving average of wake duration during sleep period in minutes with linear trendline. The bolded trace is the average for the cluster; the light grey tracings are for each individual in the cluster.
  - **Figure F11.** 7-day moving average of sedentary time in minutes with linear trendline. The bolded trace is the average for the cluster; the light grey tracings are for each individual in the cluster.
  - **Figure F12.** 7-day moving average of total sleep time in minutes with linear trendline. The bolded trace is the average for the cluster; the light grey tracings are for each individual in the cluster.

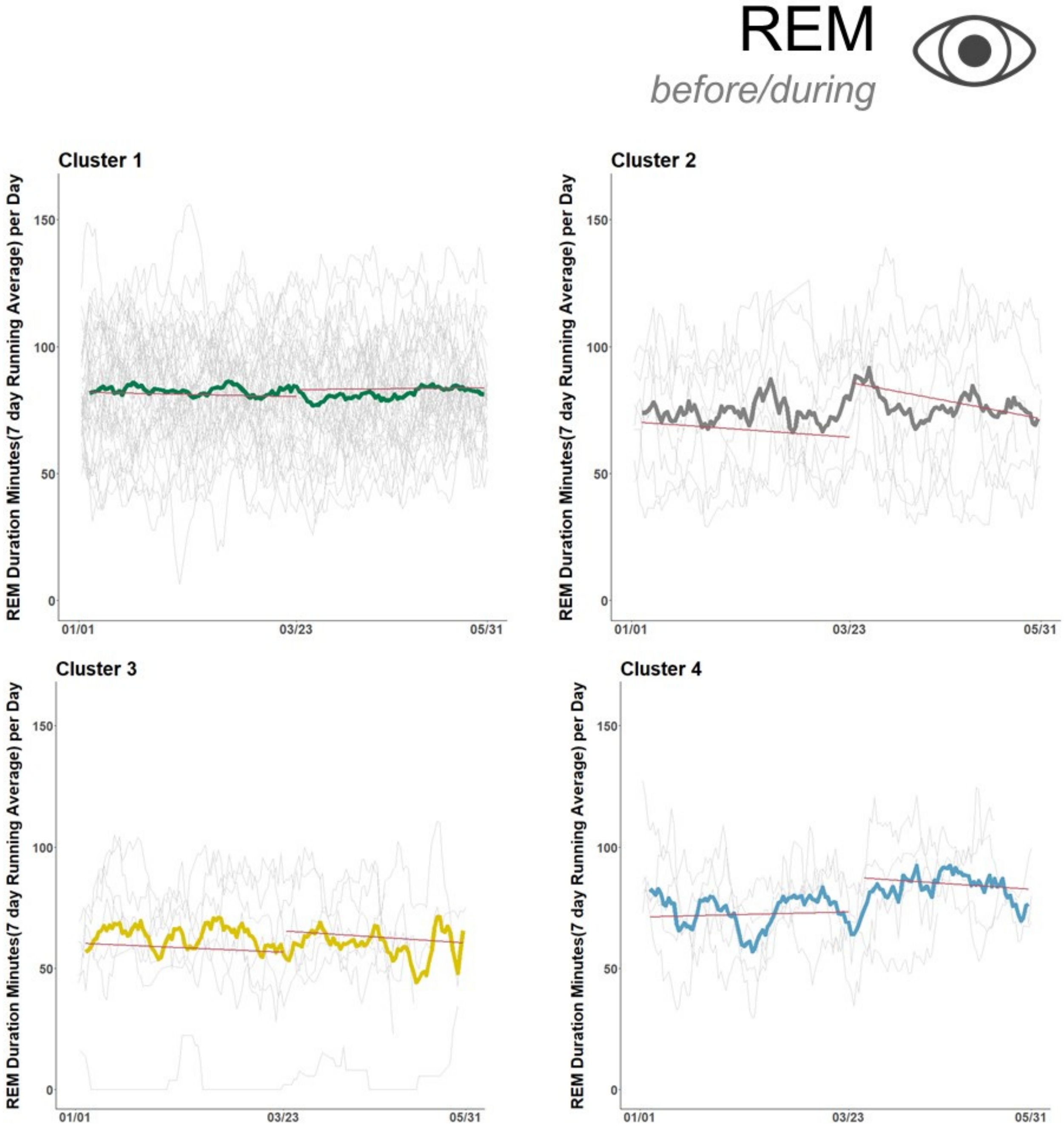


**Figure F1.** 7-day moving average of REM duration in minutes with linear trendline. The bolded trace is the average for the cluster; the light grey tracings are for each individual in the cluster.

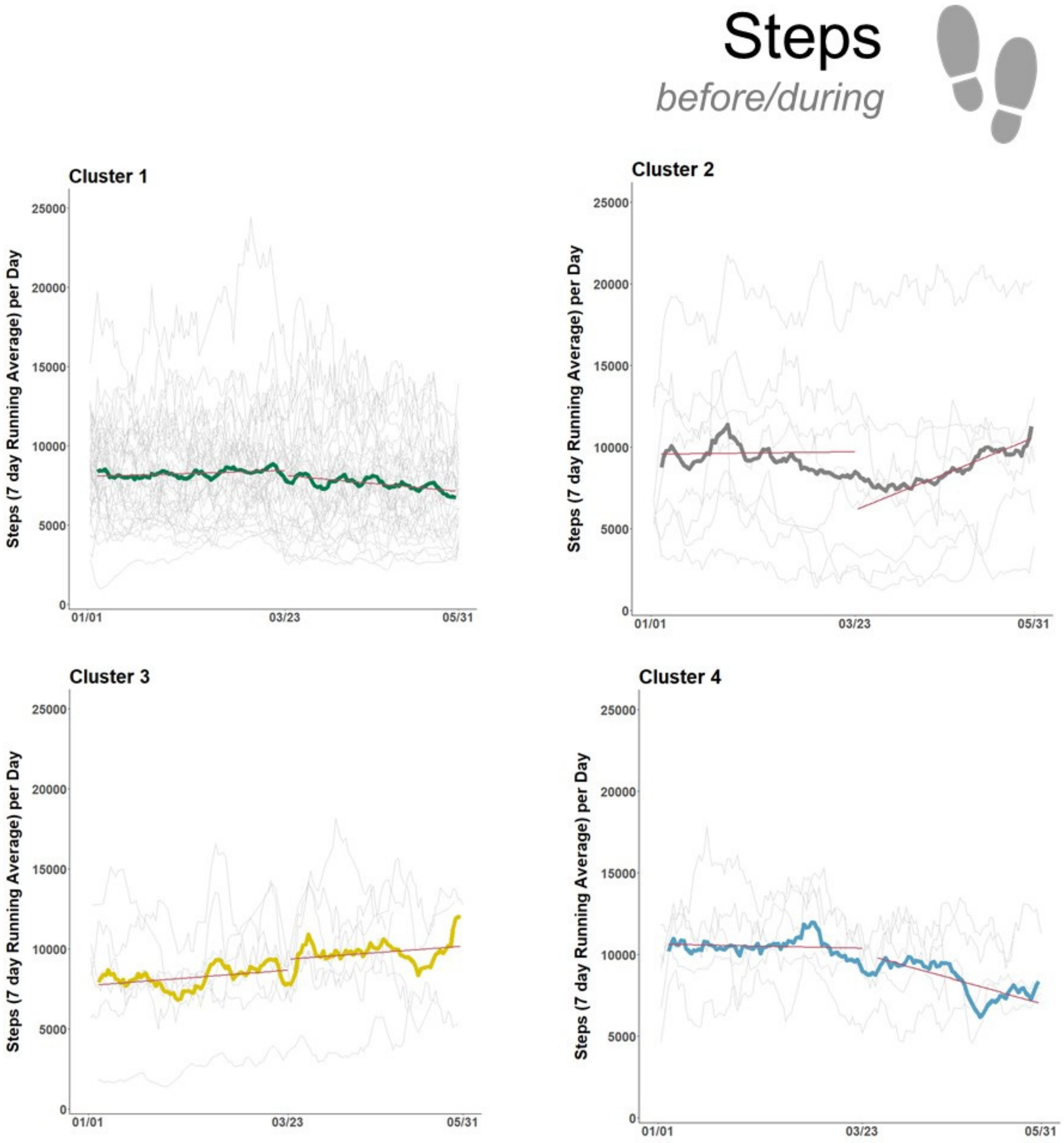


**Figure F2**. 7-day moving average of steps with linear trendline. The bolded trace is the average for the cluster; the light grey tracings are for each individual in the cluster.

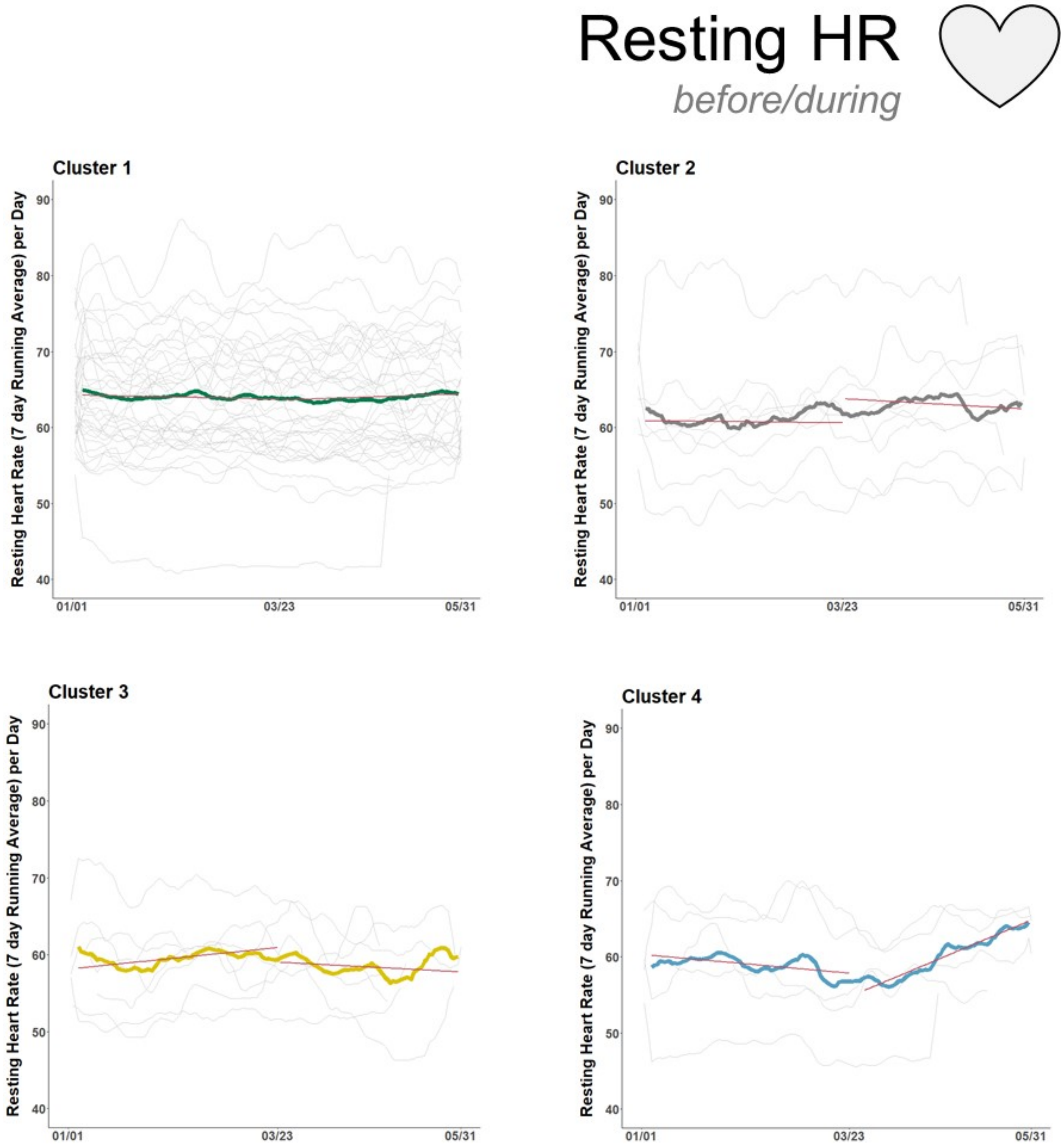


**Figure F3.** 7-day moving average of resting heart rate in beats per minute with linear trendline. The bolded trace is the average for the cluster; the light grey tracings are for each individual in the cluster.

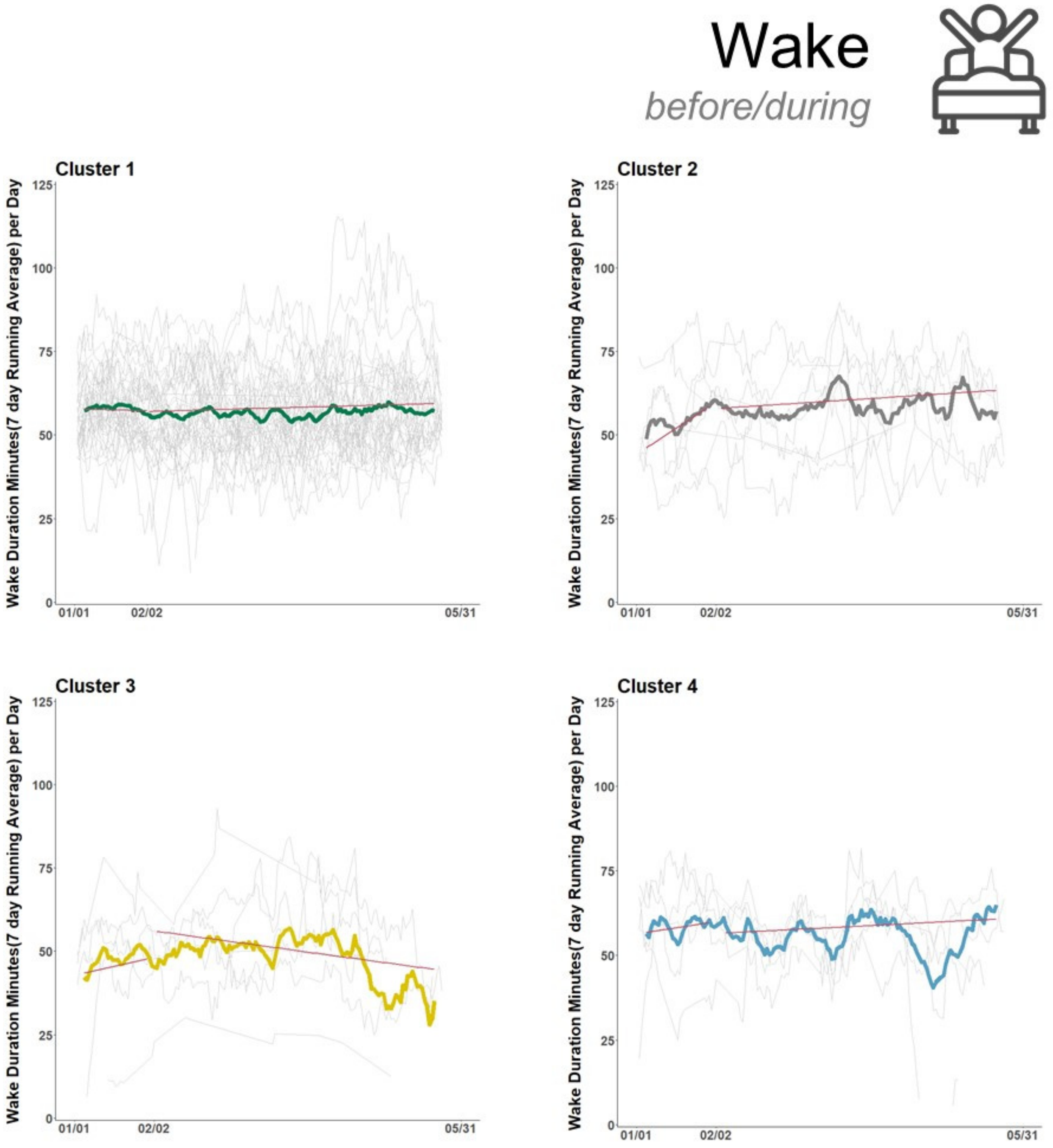


**Figure F4.** 7-day moving average of wake duration during sleep period in minutes with linear trendline. The bolded trace is the average for the cluster; the light grey tracings are for each individual in the cluster.

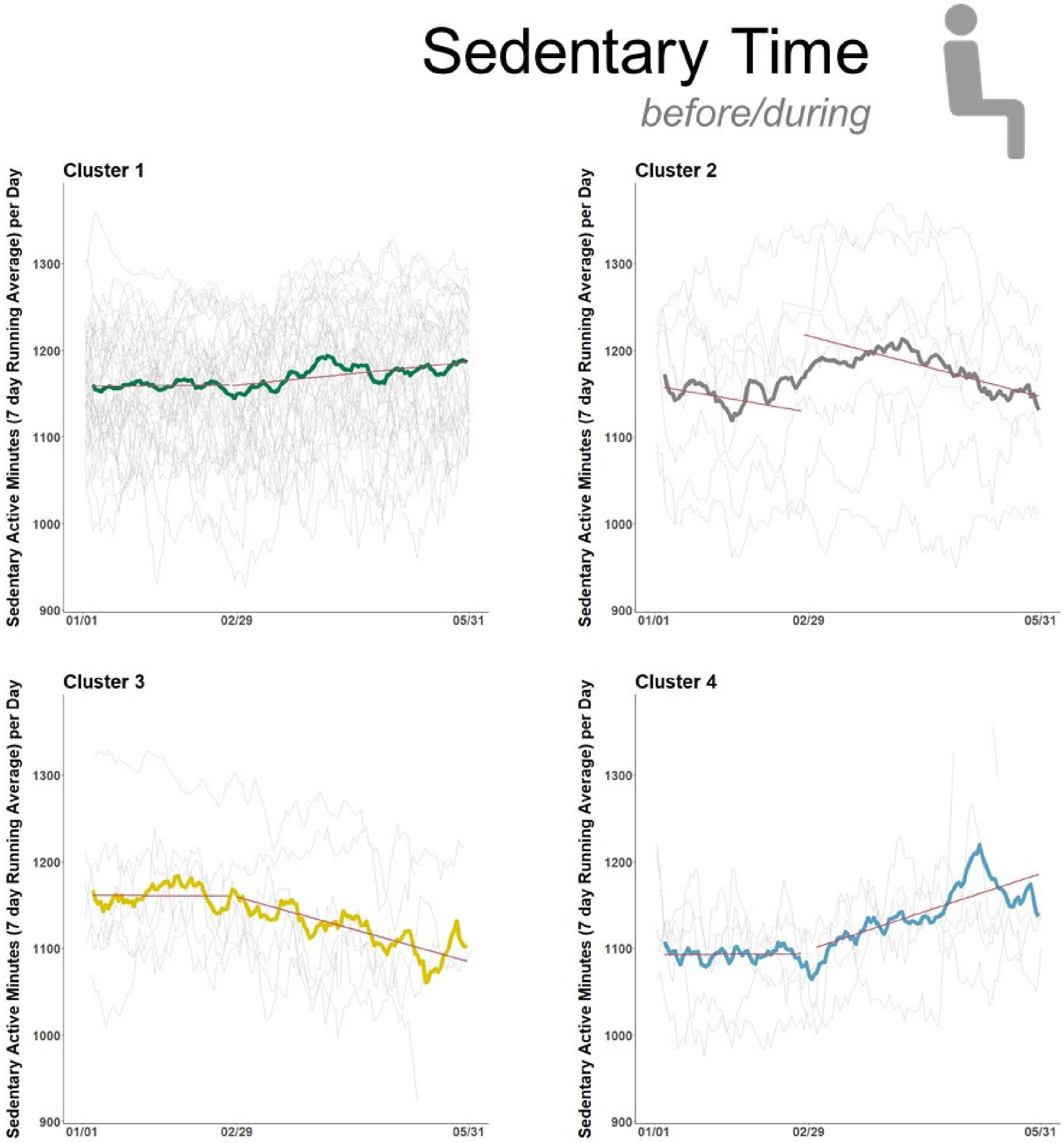


**Figure F5.** 7-day moving average of sedentary time in minutes with linear trendline. The bolded trace is the average for the cluster; the light grey tracings are for each individual in the cluster.

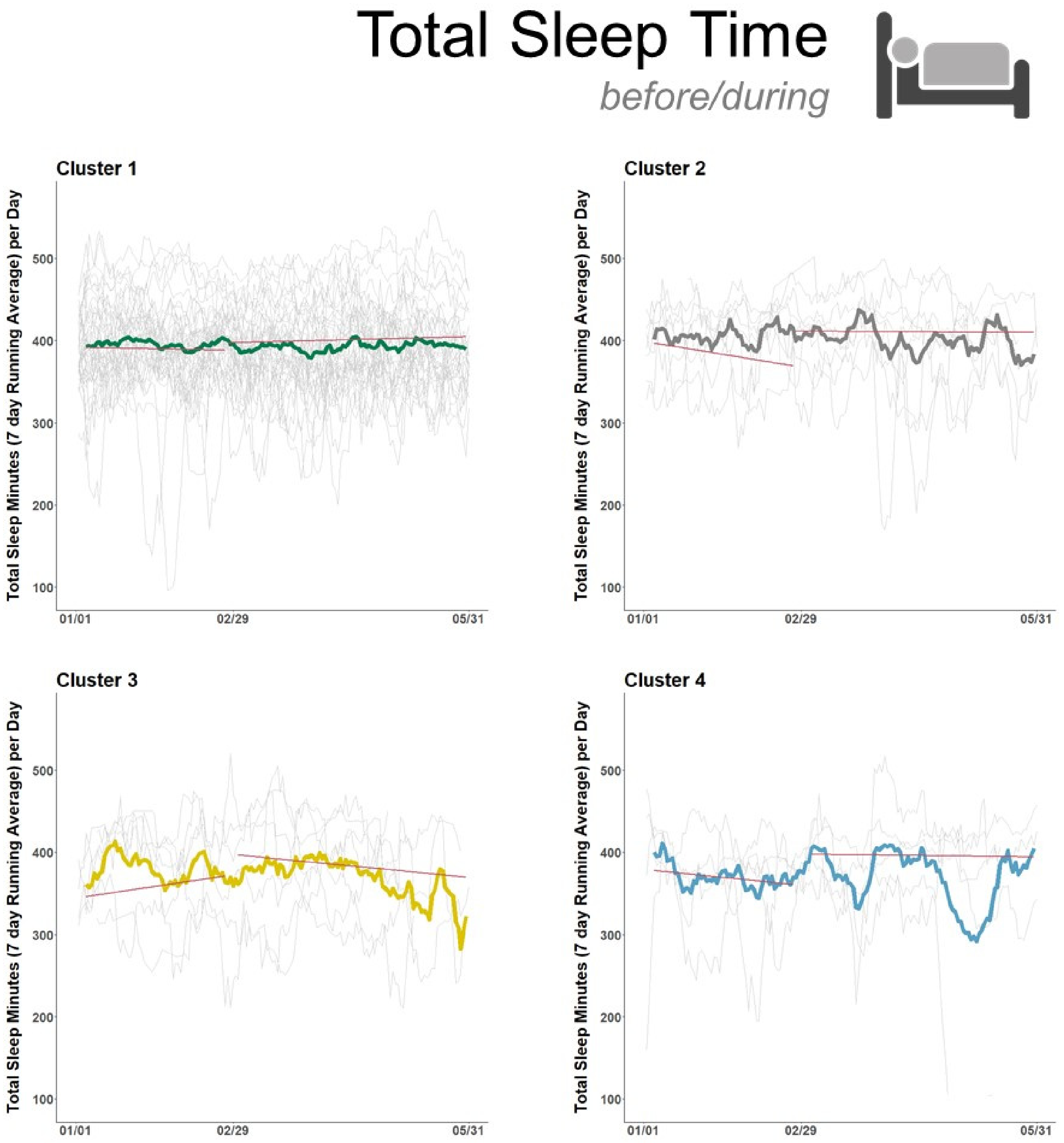


**Figure F6.** 7-day moving average of total sleep time in minutes with linear trendline. The bolded trace is the average for the cluster; the light grey tracings are for each individual in the cluster.

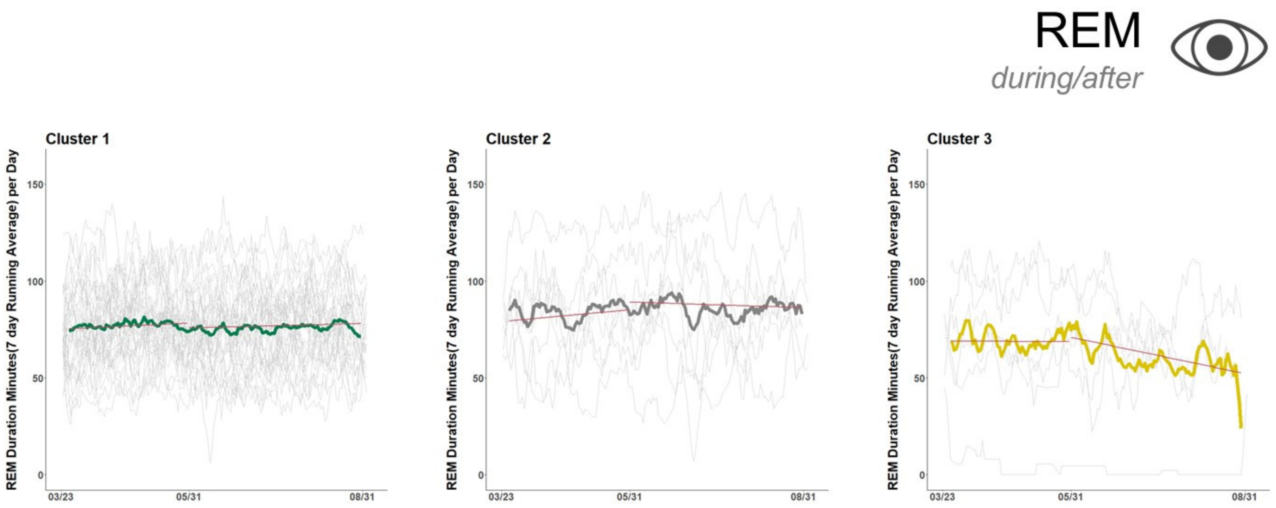


**Figure F7.** 7-day moving average of REM duration in minutes with linear trendline. The bolded trace is the average for the cluster; the light grey tracings are for each individual in the cluster.

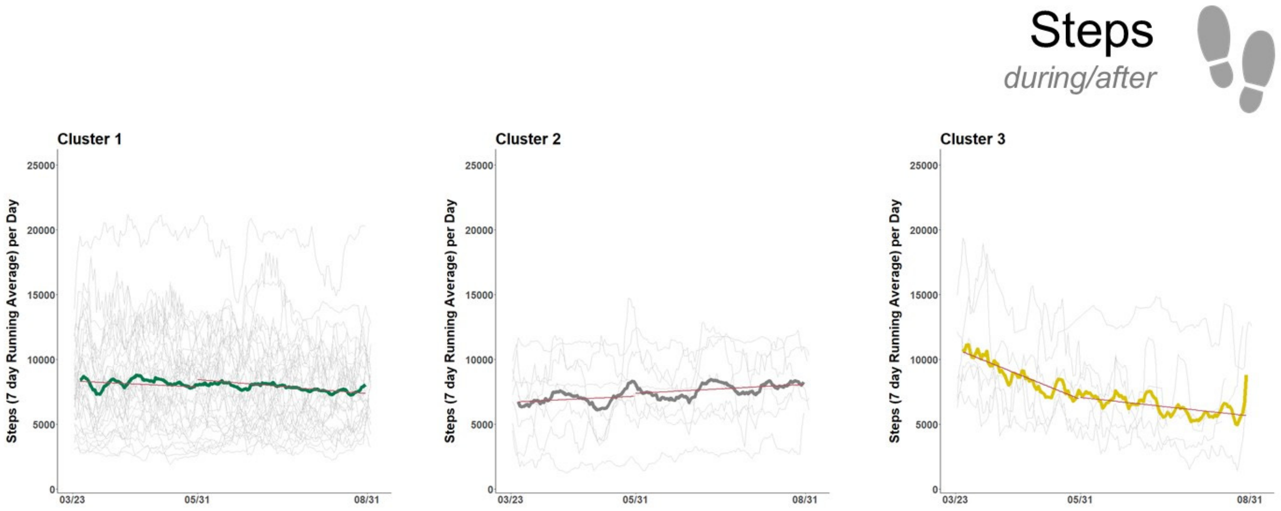


**Figure F8.** 7-day moving average of steps with linear trendline. The bolded trace is the average for the cluster; the light grey tracings are for each individual in the cluster.

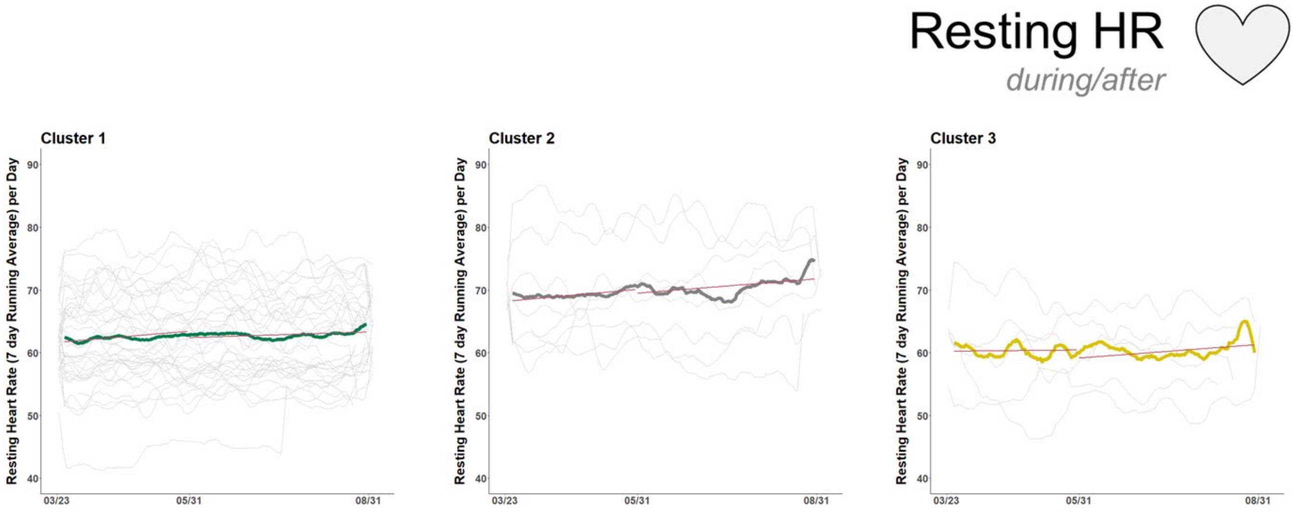


**Figure F9.** 7-day moving average of resting heart rate in beats per minute with linear trendline. The bolded trace is the average for the cluster; the light grey tracings are for each individual in the cluster.

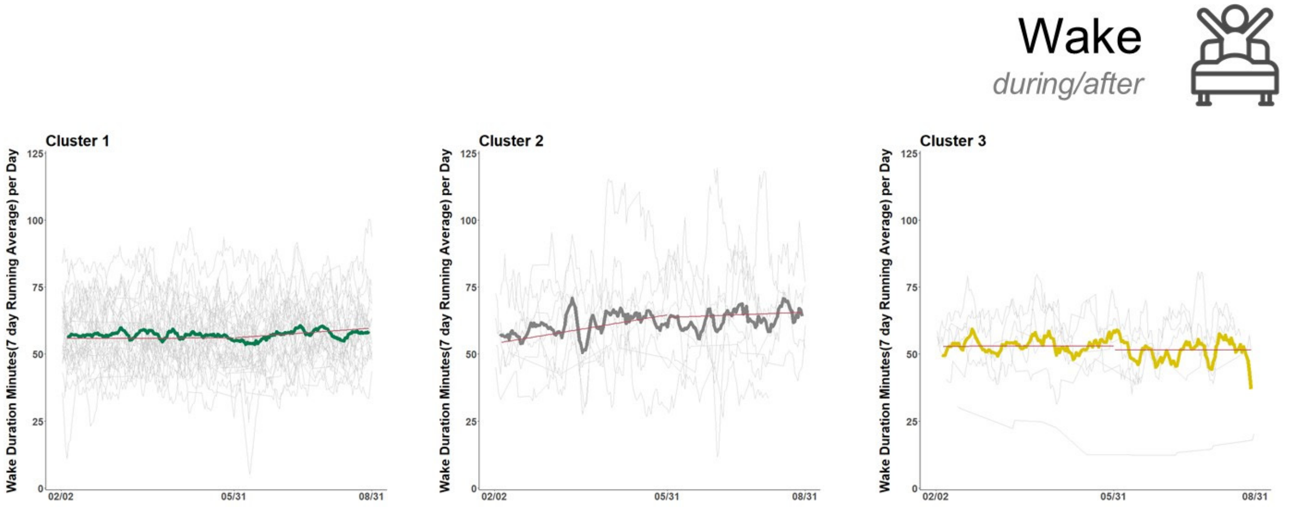


**Figure F10.** 7-day moving average of wake duration during sleep period in minutes with linear trendline. The bolded trace is the average for the cluster; the light grey tracings are for each individual in the cluster.

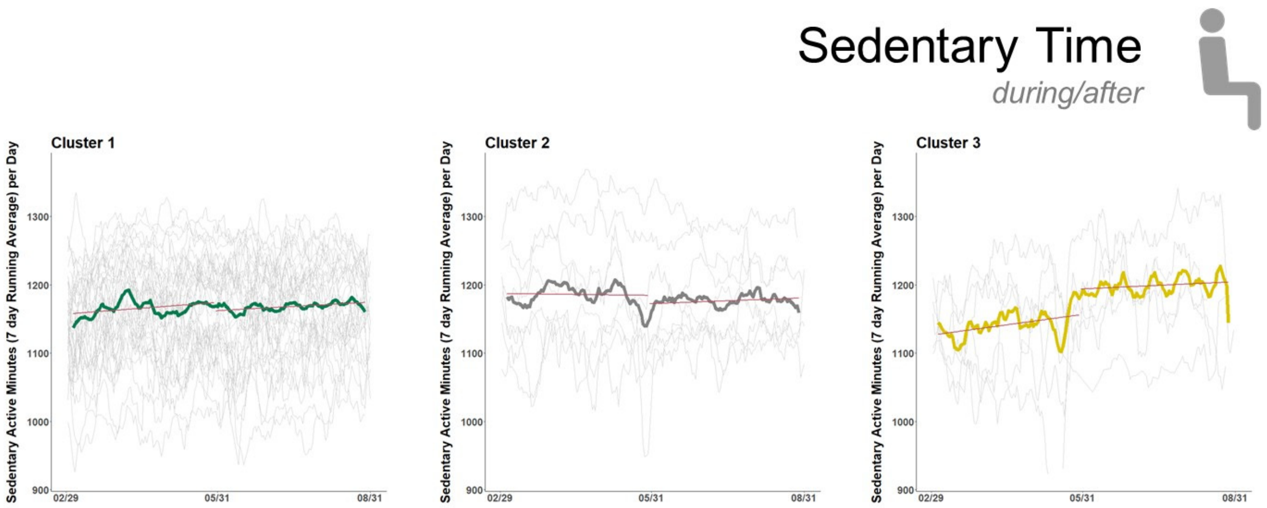


**Figure F11.** 7-day moving average of sedentary time in minutes with linear trendline. The bolded trace is the average for the cluster; the light grey tracings are for each individual in the cluster.

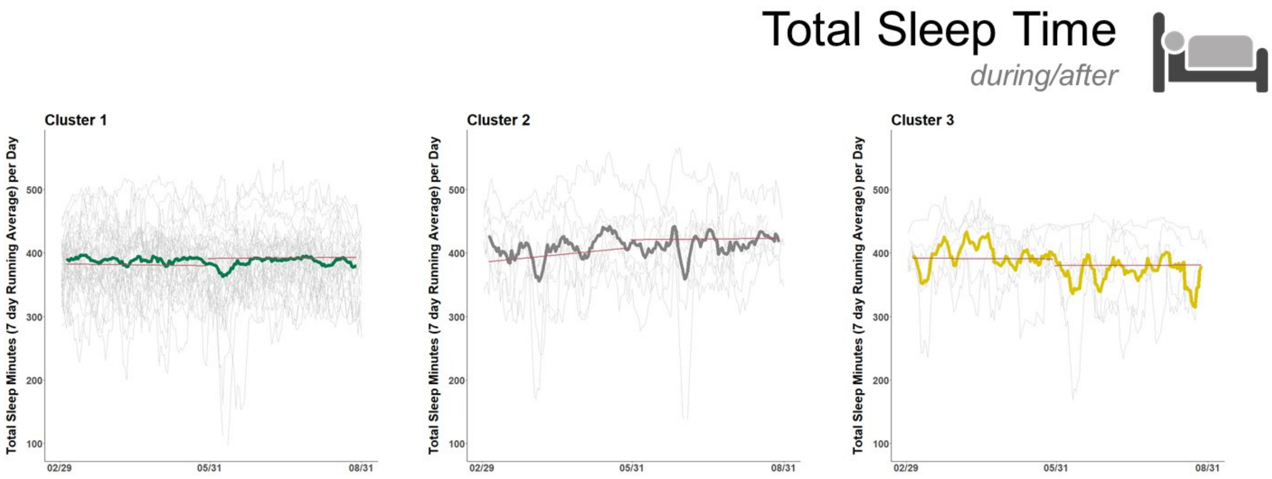


**Figure F12.** 7-day moving average of total sleep time in minutes with linear trendline. The bolded trace is the average for the cluster; the light grey tracings are for each individual in the cluster.

**Appendix G:** Self-Evaluation of Activity

Data recorded by the wearable devices for the 40 subjects in Group B differed from self-reported activity on a number of metrics including steps and sleep quality (see Figure 4, main text). For example, 55% (18/40) of subjects indicated ‘no change’ in activity while increasing their daily steps (+20.4 ± 18.6%) or while increasing sedentary time (+2.6 ± 1.7%) and decreasing steps (-15.2 ± 11.1%). 75% (30/40) of subjects indicated “no change” in the number of times they woke during the night, when the devices recorded them waking up 6-9% more or less frequently (see Tables E3 and E4 in Appendix E).

G1. Survey Questions

*Online survey accessible at*

https://quantuproject.typeform.com/report/EXJ64M/q3gvPXhVO9ZHqtcS?view_mode=print

G2. Survey Results: answers from subjects (Group B) and the public

Brain Health & the Pandemic

How has the COVID 19 pandemic altered your lifestyle and brain-healthy habits like good nutrition, exercise, social interactions & stress relief?

Notes: This survey and all questions are voluntary. We will use survey results to study how the pandemic and stay-at-home orders are affecting lifestyle factors that influence our brains' health. Data is anonymized and aggregated prior to any analysis. Our goal is to develop approaches that mitigate long-term risks to cognitive health.

1. Are you currently enrolled in the Quantu project? Yes or no

a. If yes, what is your name? (last name, first name)

---

If no… (only for survey participants NOT in Quantu study.

2. What is your age?

3. Gender: male, female, other

4. Race: African American or Black, American Indian or Alaska Native, Asian, Native Hawaiian or Pacific Islander, White, Multiracial, Other

5. Ethnicity: Hispanic or Latino, not Hispanic or Latino

6. What is the highest level of education you have achieved: less than a high school diploma, high school degree or equivalent, Associates degree, Bachelor's degree (e.g. BA or BS), Master's Degree (e.g. MA, MS, MBA), Doctorate (e.g. PhD, EdD) or Professional Degree (e.g. MD, JD)

7. Do you have children under the age of 18 in the home: yes or no

8. What is your household income per year? Below $10K, $10K - $35K, $35 - $50K, $50K - $75K, $75K - $100K, $100K or more

---

9. How many people live in your household?
10. Do you have children under the age of 18 in the home?
11. Please describe your household:
    a. I live alone.
    b. I live with immediate family. (Spouse/partner, children)
    c. I live with extended family (Grandchildren, etc.)
    d. I live with a roommate.
    e. I live with multiple roommates.
    f. I live in a facility (nursing home, hospital, assisted living)
    g. other

12. In which city or region are you located?

   a. Houston area
   b. San Antonio area
   c. Los Angeles area
   d. CA Bay area
   e. Other

13. How would you describe the area in which you live?

   a. Urban
   b. Suburban
   c. Rural (within 25 miles of the city)
   d. Rural (more than 25 miles)

14. Is your area currently under shelter-in-place orders (federal, state, or local)?

   a. Yes
   b. No
   c. Unsure

15. Have you tried to comply with shelter-in-place orders?

   a. Yes
   b. No

16. Were you working outside of the home prior to shelter-in-place orders?

   a. Full time
   b. Part time
   c. Retired
   d. Unemployed

17. How has/had your employment status changed while stay-at-home orders were issued? Check as many as apply.

   a. No change
   b. Increased hours
   c. Reduced hours
   d. Furloughed (anticipating going back to the same employer)
   e. Laid off (not anticipating going back to the same employer)
   f. Working partially/exclusively from home
   g. Other

18. Are you considered an essential worker?

   a. Yes

b. No
c. Unsure

19. What was your activity level in the months before Covid19 stay-at-home orders?

a. High to very high (>10K steps/day, running, daily workouts, etc.)
b. Moderate to high (5-10K steps/day, workouts a few times a wk)
c. Limited to moderate (2-5K steps/day, occasional workouts)
d. Sedentary (<2K steps/day, no workouts)

20. What is your activity level since Covid19 stay-at-home orders were issued?

a. High to very high (>10K steps/day, running, daily workouts, etc.)
b. Moderate to high (5-10K steps/day, workouts a few times a week)
c. Limited to moderate (2-5K steps/day, occasional workouts)
d. Sedentary (<2K steps/day, no workouts)

21. Have the stay-at-home orders affected your activity levels?

a. Yes
b. No
c. Unsure

22. What are the main reasons for your activity changes?

a. My motivation to exercise changed with the pandemic.
b. Gyms and fitness centers are closed.
c. I have been limiting time spent outside.
d. I usually exercise in a group/social circle that are limiting contact.
e. Illness
f. Other:

23. What was your level of remote/virtual interactions in the months before stay-at-home orders? This includes phone calls, video conferencing, online/text chatting, etc.

a. High to very high (daily)
b. Moderate to high (weekly)
c. Limited to moderate (occasional)
d. Low (rarely/never)

24. How have your remote/virtual social interactions changed? This includes phone calls, video conferencing, online chatting, etc.?

a. Decreased in frequency
b. Remained the same
c. Increased in frequency

25. What was your level of in-person interactions in the months before stay-at-home orders In-person interactions can include a partner, children or colleagues?

a. High to very high (daily)
b. Moderate to high (weekly)
c. Limited to moderate (occasional)
d. Low (rarely/never)

26. How have your in-person interactions changed?

a. Decreased in frequency
b. Remained the same
c. Increased in frequency

27. Do you feel isolated?

28. How has your diet changed since stay-at-home orders were in place?

a. Healthier
b. Less healthy
c. Remained the same

29. How has your sleep changed since stay-at-home orders were in place? Check all that apply

a. Less consistent sleep schedule
b. More consistent sleep schedule
c. Shorter in duration
d. Longer in duration
e. Waking earlier
f. Waking later
g. Earlier bedtime
h. Later bedtime
i. More naps
j. Fewer naps
k. Waking more frequently during the night
l. Waking less frequently during the night
m. No change

30. Have you been doing anything for stress relief?

31. If so, what have you been doing to relieve stress?

A quick note on ways Quantu Project volunteers and team members are helping fellow volunteers, friends, and our communities:

Remote Exercise (https://www.quantuproject.org/onlineexercise): Partnering with trainer Teofilo (Teo) Reyes we are offering remote exercise classes to all volunteers and friends Monday, Wednesdays and Fridays through May 15.

Recovery & Resource website (https://www.covid19recoverytexas.org/): With the help of students and faculty at UTSA, we built a Recovery & Resource website that includes pages listing resources (like options for grocery delivery or financial help), dates when institutes open, and daily living tips. The site works by crowdsourcing information from the public. Everyone can make a difference by providing updates.

Other volunteering: Quantu Project members are also volunteering with Big Brothers Big Sisters to remotely teach computer programming to children. The Recovery website also provides additional suggestions on volunteering (https://www.covid19recoverytexas.org/help).

Contact us if you're interested in joining these efforts or have suggestions.

32. Any other questions or suggestions?

Thank you – The Quantu Project Team @quantuproject.org

Add in the following questions:

What is your activity level after Covid19 stay-at-home orders were lifted?

a. High to very high (>10K steps/day, running, daily workouts, etc.)
b. Moderate to high (5-10K steps/day, workouts a few times a week)
c. Limited to moderate (2-5K steps/day, occasional workouts)
d. Sedentary (<2K steps/day, no workouts)
e. I am currently under stay-at-home orders.

What is your level of remote/virtual interactions after stay-at-home orders were lifted? This includes phone calls, video conferencing, online/text chatting, etc.

a. High to very high (daily)
b. Moderate to high (weekly)
c. Limited to moderate (occasional)
d. Low (rarely/never)
e. I am currently under stay-at-home orders.

What is your level of in-person interactions after stay-at-home orders were lifted. In-person interactions can include a partner, children or colleagues?

a. High to very high (daily)

b. Moderate to high (weekly)
c. Limited to moderate (occasional)
d. Low (rarely/never)

How has/had your employment status changed after stay-at-home orders were lifted? Check as many as apply.

a. No change
b. Increased hours
c. Reduced hours
d. Furloughed (anticipating going back to the same employer)
e. Laid off (not anticipating going back to the same employer)
f. Working partially/exclusively from home
g. Other

### Brain Health Survey

267 responses

---

Are you currently enrolled in the Quantu Project?

266 out of 267 answered

| | | | |
|---|---|---|---|
| 1 | Yes | 35% | / 93 resp. |
| 2 | No | 65% | / 173 resp. |

---

What is your gender?

172 out of 267 answered

| | | | |
|---|---|---|---|
| 1 | female | 69% | / 119 resp. |
| 2 | male | 30% | / 53 resp. |
| 3 | Other | 0% | / 0 resp. |

What is your race?

168 out of 267 answered

| | | | |
|---|---|---|---|
| 1 | White | 81% | / 136 resp. |
| 2 | Asian | 6% | / 11 resp. |
| 3 | Multiracial | 4% | / 8 resp. |
| 4 | African American or Black | 3% | / 5 resp. |
| 5 | American Indian or Alaska Native | 0% | / 0 resp. |
| 6 | Native Hawaiian or Pacific Islander | 0% | / 0 resp. |
| 7 | Other | 4% | / 8 resp. |

Are you Hispanic?

170 out of 267 answered

| | | | |
|---|---|---|---|
| 1 | Yes | 18% | / 31 resp. |
| 2 | No | 81% | / 139 resp. |

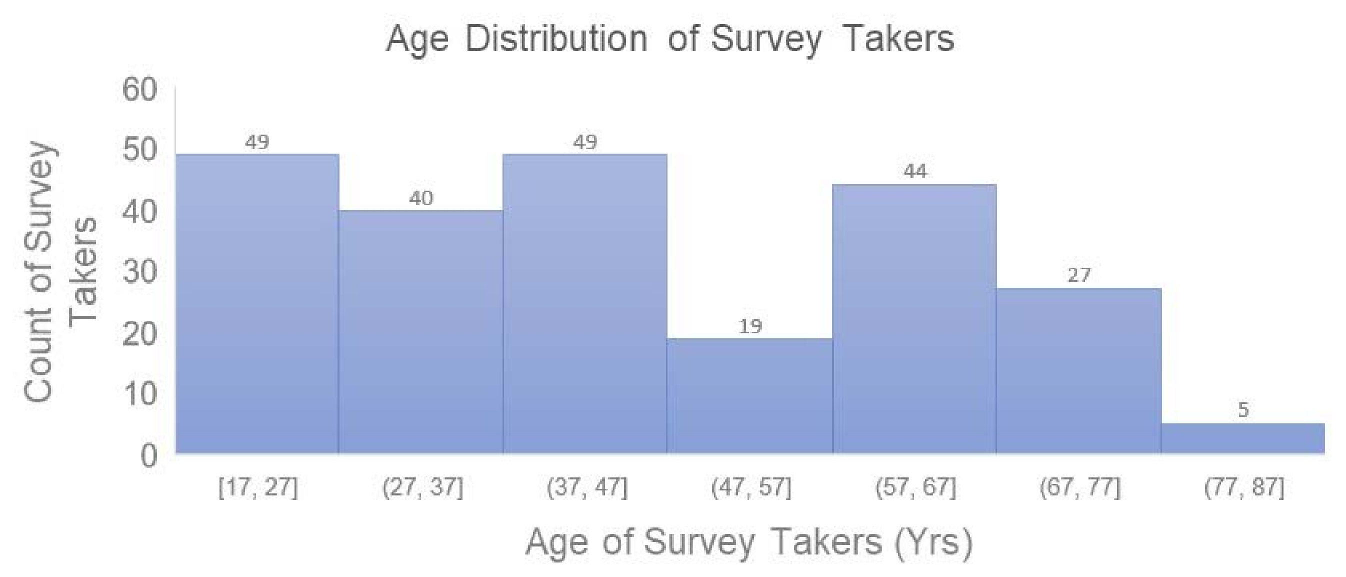
Age Distribution of Survey Takers
Count of Survey Takers
60
50
40
30
20
10
0
49
40
49
19
44
27
5
[17, 27]
(27, 37]
(37, 47]
(47, 57]
(57, 67]
(67, 77]
(77, 87]
Age of Survey Takers (Yrs)

What is the highest level of education you have achieved?

172 out of 267 answered

| | | | |
|---|---|---|---|
| 1 | Bachelor's degree (e.g. BA or BS) | 46% | / 80 resp. |
| 2 | Master's degree (e.g. MA, MS, MBA) | 22% | / 39 resp. |
| 3 | Doctorate or Professional degree (e.g. MD, JD, PhD) | 12% | / 22 resp. |
| 4 | Associates degree or Vocational Certification | 11% | / 19 resp. |
| 5 | high school degree or equivalent | 7% | / 12 resp. |
| 6 | less than a high school diploma | 0% | / 0 resp. |

What is your household income per year?

163 out of 267 answered

| | | | |
|---|---|---|---|
| 1 | $100K or more | 42% | / 70 resp. |
| 2 | $75K - $100K | 19% | / 32 resp. |
| 3 | $50K - $75K | 14% | / 24 resp. |
| 4 | $35K - $50K | 12% | / 20 resp. |
| 5 | $10K - $35K | 8% | / 13 resp. |
| 6 | Below $10K | 2% | / 4 resp. |

Do you have children under the age of 18 in the home?

264 out of 267 answered

| | | | |
|---|---|---|---|
| 1 | Yes | 23% | / 63 resp. |
| 2 | No | 76% | / 201 resp. |

Please describe your household

262 out of 267 answered

1. I live with immediate family. (Spouse/partner, children) — 67% / 177 resp.
2. I live alone. — 21% / 55 resp.
3. I live with extended family (Grandchildren, etc.) — 5% / 13 resp.
4. I live with a roommate. — 3% / 8 resp.
5. I live with multiple roommates. — 2% / 6 resp.
6. I live in a facility (nursing home, hospital, assisted living) — 0% / 0 resp.
7. Other — 1% / 3 resp.

In which city or region are you located?

258 out of 267 answered

| # | Option | Percent | Responses |
|---|---|---|---|
| 1 | San Antonio area | 51% | / 132 resp. |
| 2 | Los Angeles area | 9% | / 24 resp. |
| 3 | Houston area | 7% | / 18 resp. |
| 4 | CA Bay Area | 3% | / 10 resp. |
| 5 | Other | 28% | / 74 resp. |

How would you describe the area in which you live?

261 out of 267 answered

| # | Option | Percent | Responses |
|---|---|---|---|
| 1 | Suburban | 52% | / 138 resp. |
| 2 | Urban | 35% | / 92 resp. |
| 3 | Rural (within 25 miles of the city) | 7% | / 20 resp. |
| 4 | Rural (more than 25 miles from the city) | 4% | / 11 resp. |

Is your area currently under shelter-in-place orders (federal, state, or local) ?

264 out of 267 answered

| | | |
|---|---|---|
| 1 | yes | 66% / 175 resp. |
| 2 | no | 23% / 61 resp. |
| 3 | unsure | 10% / 28 resp. |

Have you tried to comply with shelter-in-place orders?

263 out of 267 answered

| | | |
|---|---|---|
| 1 | Yes | 96% / 253 resp. |
| 2 | No | 3% / 10 resp. |

Were you working outside of the home prior to shelter-in-place orders?

260 out of 267 answered

| | | | |
|---|---|---|---|
| 1 | Full time | 51% | / 133 resp. |
| 2 | Retired | 25% | / 66 resp. |
| 3 | Part time | 16% | / 43 resp. |
| 4 | Unemployed | 6% | / 18 resp. |

How has your employment status changed while stay-at-home orders were issued? Check all that apply.

262 out of 267 answered

| | | |
|---|---|---|
| 1 | No change | 52% / 137 resp. |
| 2 | Working partially/exclusively from home | 33% / 88 resp. |
| 3 | Reduced hours | 10% / 28 resp. |
| 4 | Increased hours | 6% / 18 resp. |
| 5 | Furloughed (anticipating going back to the same employer) | 5% / 14 resp. |
| 6 | Laid off (not anticipating going back to the same employer) | 3% / 9 resp. |
| 7 | Other | 2% / 7 resp. |

How has/had your employment status changed **AFTER** stay-at-home orders were lifted compared to before the stay-at-home order? Check as many as apply.

158 out of 267 answered

| | | | |
|---|---|---|---|
| 1 | No change | 42% | / 67 resp. |
| 2 | I am currently under a stay-at-home order. | 31% | / 49 resp. |
| 3 | Working partially/exclusively from home | 24% | / 38 resp. |
| 4 | Reduced hours | 5% | / 9 resp. |
| 5 | Increased hours | 4% | / 7 resp. |
| 6 | Laid off (not anticipating going back to the same employer) | 2% | / 4 resp. |
| 7 | Furloughed (anticipating going back to the same employer) | 1% | / 3 resp. |
| 8 | Other | 3% | / 5 resp. |

Are you considered an essential worker?

264 out of 267 answered

| | | |
|---|---|---|
| 1 | no | 62% / 166 resp. |
| 2 | yes | 29% / 79 resp. |
| 3 | unsure | 7% / 19 resp. |

What was your activity level in the months **before** Covid19 stay-at-home orders were issued?

263 out of 267 answered

| | | |
|---|---|---|
| 1 | moderate to high (5-10K steps/day, workouts a few times a wk) | 45% / 119 resp. |
| 2 | limited to moderate (2-5K steps/days, occasional workouts) | 26% / 70 resp. |
| 3 | high to very high (>10K steps/day, running, daily workouts, etc) | 21% / 57 resp. |
| 4 | sedentary (<2 K steps/day, no workouts) | 6% / 16 resp. |
| 5 | Other | 0% / 1 resp. |

What is your activity level since Covid19 stay-at-home orders were issued?

262 out of 267 answered

| | | | |
|---|---|---|---|
| 1 | limited to moderate (2-5K steps/days, occasional workouts) | 34% | / 90 resp. |
| 2 | moderate to high (5-10K steps/day, workouts a few times a wk) | 31% | / 83 resp. |
| 3 | sedentary (<2 K steps/day, no workouts) | 19% | / 50 resp. |
| 4 | high to very high (>10K steps/day, running, daily workouts, etc) | 14% | / 37 resp. |
| 5 | Other | 0% | / 2 resp. |

What is your activity level **after** COVID19 stay-at-home orders were lifted?

156 out of 267 answered

| | | | |
|---|---|---|---|
| 1 | I am currently under stay-at-home orders. | 34% | / 53 resp. |
| 2 | limited to moderate (2-5K steps/days, occasional workouts) | 24% | / 38 resp. |
| 3 | moderate to high (5-10K steps/day, workouts a few times a wk) | 17% | / 27 resp. |
| 4 | sedentary (<2 K steps/day, no workouts) | 13% | / 21 resp. |
| 5 | high to very high (>10K steps/day, running, daily workouts, etc) | 10% | / 17 resp. |

Have the stay-at-home orders or other aspects of the pandemic affected your activity levels?

263 out of 267 answered

| # | Answer | Percent | Responses |
|---|---|---|---|
| 1 | yes | 68% | 180 resp. |
| 2 | no | 28% | 75 resp. |
| 3 | unsure | 3% | 8 resp. |

What are the main reasons for your activity changes?

178 out of 267 answered

| # | Answer | Percent | Responses |
|---|---|---|---|
| 1 | My motivation to exercise changed | 46% | 83 resp. |
| 2 | Gyms and fitness centers are closed | 43% | 77 resp. |
| 3 | I have been limiting time spent outside | 34% | 61 resp. |
| 4 | I usually exercise in a group/social circle that are limiting contact. | 23% | 41 resp. |
| 5 | Illness (yourself or family) | 6% | 11 resp. |
| 6 | Other | 14% | 26 resp. |

What was your level of remote/virtual interactions in the months **before** stay-at-home orders? This includes phone calls, video conferencing, online/text chatting, etc.

264 out of 267 answered

1 limited to moderate (occasional) 42% / 111 resp.

2 moderate to high (weekly) 29% / 77 resp.

3 high to very high (daily) 15% / 40 resp.

4 low (rarely/never) 13% / 36 resp.

How have your remote / virtual social interactions changed since stay-at-home orders have been in place?

264 out of 267 answered

1 Increased in frequency 67% / 177 resp.

2 Remained the same 26% / 69 resp.

3 Decreased in frequency 6% / 18 resp.

What is your level of remote/virtual interactions **after** stay-at-home orders were lifted? This includes phone calls, video conferencing, online/text chatting, etc.

156 out of 267 answered

1. I am currently under stay-at-home orders. 34% / 53 resp.
2. high to very high (daily) 23% / 37 resp.
3. moderate to high (weekly) 21% / 34 resp.
4. limited to moderate (occasional) 17% / 27 resp.
5. low (rarely/never) 3% / 5 resp.

What was your level of in-person interactions in the months **before** stay-at-home orders? In-person interactions can include a partner, children or colleagues.

261 out of 267 answered

1. high to very high (daily) 59% / 155 resp.
2. moderate to high (weekly) 26% / 68 resp.
3. limited to moderate (occasional) 12% / 33 resp.
4. low (rarely/never) 1% / 5 resp.

How have your in-person interactions changed?

263 out of 267 answered

1. Decreased in frequency — 79% / 210 resp.
2. Remained the same — 17% / 46 resp.
3. Increased in frequency — 2% / 7 resp.

---

What is your level of in-person interactions **after** stay-at-home orders were lifted. In-person interactions can include a partner, children or colleagues?

158 out of 267 answered

1. I am currently under stay-at-home orders. — 40% / 64 resp.
2. limited to moderate (occasional) — 22% / 36 resp.
3. moderate to high (weekly) — 13% / 21 resp.
4. high to very high (daily) — 12% / 19 resp.
5. low (rarely/never) — 11% / 18 resp.

Do you feel isolated?

263 out of 267 answered

| | | |
|---|---|---|
| 1 | Yes | 38% / 101 resp. |
| 2 | No | 61% / 162 resp. |

How has your diet changed since the stay-at-home orders were in place?

263 out of 267 answered

| | | |
|---|---|---|
| 1 | Remained the same | 47% / 124 resp. |
| 2 | Become less healthy | 26% / 70 resp. |
| 3 | Become healthier | 26% / 69 resp. |

How has your sleep changed since the stay-at-home orders were in place?

265 out of 267 answered

| | | |
|---|---|---|
| 1 | Less consistent sleep schedule | 34% / 92 resp. |
| 2 | Later bedtime | 27% / 72 resp. |
| 3 | Waking more frequently during the night | 26% / 69 resp. |
| 4 | Waking later | 25% / 67 resp. |
| 5 | No change | 23% / 63 resp. |
| 6 | Longer in duration | 20% / 53 resp. |
| 7 | Shorter in duration | 18% / 50 resp. |
| 8 | More naps | 17% / 47 resp. |
| 9 | More consistent sleep schedule | 13% / 36 resp. |
| 10 | Waking earlier | 13% / 36 resp. |
| 11 | Earlier bedtime | 7% / 21 resp. |
| 12 | Waking less frequently during the night | 1% / 5 resp. |
| 13 | Fewer naps | 1% / 4 resp. |

Have you been doing anything for stress relief?

266 out of 267 answered

| | | |
|---|---|---|
| 1 | Yes | 60% / 162 resp. |
| 2 | No | 39% / 104 resp. |
